# Neighborhood, household, and individual socioeconomic position in early life and childhood cardiovascular health measures: an international cross-cohort study

**DOI:** 10.1101/2025.11.16.25340203

**Authors:** Kavindi Gamage, Susanna Pätsi, Tong Chen, Kate Mooney, Sylvain Sebert, Naomi Priest, Toby Mansell, David Burgner, Barwon Infant Study Investigator Group

**Author notes:** Joint last authors (Naomi Priest, Toby Mansell, David Burgner). Group authors/consortia. **Corresponding author:** Kavindi Gamage; The University of Melbourne, Grattan Street, Parkville, Victoria 3010, Australia.

## Abstract

**Background:** Cardiovascular disease (CVD) is socioeconomically patterned and risk accrues from early life. Evidence in younger populations is scarce. We investigated the extent to which early life socioeconomic position (SEP) affects cardiovascular health (CVH) in children and adolescents, and the extent of mediation by body mass index (BMI).

**Methods and Results:** We analysed 5 longitudinal cohorts: the Barwon Infant Study (BIS, Australia, mean age 4.1 years, n=708); Born in Bradford (United Kingdom, mean age 9.3 years, n=4576); the Longitudinal Study of Australian Children’s Child Health CheckPoint (LSAC-CP, Australia, mean 12.0 years, n=1874); the Northern Finland Birth Cohort 1986 (NFBC1986) (Finland, mean 16.0 years, n=9467); and the Avon Longitudinal Study of Parents and Children (ALSPAC, United Kingdom, mean 17.8 years, n=4875). Exposures were neighborhood disadvantage, household SEP, and maternal education (in pregnancy/at birth). Outcomes were carotid intima-media thickness, pulse wave velocity, blood pressure, and lipids (in childhood/adolescence). From 12 years, those with lower SEP had worse CVH, adjusted for age, sex, and ethnicity. For example, LSAC-CP 12-year-olds in the most disadvantaged neighborhoods had higher pulse wave velocity (β = 0.1 m/s; 95% CI, 0.03-0.2; *P*=0.003) than those in the most advantaged, and NFBC1986 16-year-olds whose mothers had the lowest education had higher triglycerides (β = 0.07 mmol/L; 95% CI, 0.0-0.1; *P*=0.04) than those whose mothers had the highest education. Hypothetical reductions in CVH differences by shifting BMI distributions varied by cohort and exposure.

**Conclusions:** The adverse effects of lower SEP on CVH are apparent from mid-childhood. Prevention should address intermediate mechanisms and upstream neighborhood, household, and maternal factors.

**Clinical Perspective:** *What Is New?:*

- This is the first and largest multicohort study to date to investigate the effects of multiple indicators SEP on CVH across multiple time points in childhood and adolescence, comprehensively assessed by vascular measures and lipids.
- Effects of low SEP on adverse CVH were consistent across several high-income contexts from 12 years of age.
- BMI was an important mediator in most relationships.

*What Are the Clinical Implications?:*

- The effects of socioeconomic conditions on CVH are likely established by mid-childhood.
- Clinicians must advocate for policy efforts to provide equitable socioeconomic resources for neighborhoods, families and children.
- Intervening on BMI may reduce socioeconomic inequalities in CVH in children and adolescents.

## Introduction

Cardiovascular disease (CVD) is the leading cause of morbidity and mortality worldwide.^1^ Despite widespread secondary prevention efforts, the global burden of CVD continues to rise.^2^ In high-income countries, there are marked population-level socioeconomic inequalities.^1, 3^ Lower socioeconomic position (SEP), including lower educational attainment, lower income, and neighborhood deprivation, is associated with a higher risk of CVD, a pattern that persists across the socioeconomic gradient.^4^ As socioeconomic inequality increases globally, previous declines in CVD rates in high-income countries are now beginning to plateau—or in some instances, reverse.^1, 5^

The pathogenesis of atherosclerotic CVD begins early in life. Lesions indicative of early atherosclerosis are present *in utero* and infancy, and are strongly associated with socioeconomically patterned maternal risk factors such as smoking and hypercholesterolemia.^6, 7^ The risk of CVD accumulates across the life course, and cardiovascular health (CVH) from childhood onwards is predictive of adult CVD.^8, 9^ Early life is therefore increasingly recognized as a critical but largely untapped opportunity to mitigate the rising CVD burden and associated inequalities. However, there is a paucity of data from children and adolescents. While some studies have shown that lower SEP is associated with adverse cardiovascular phenotypes in pediatric populations,^10, 11, 12^ most include a wide range of ages in the analytical sample and focus on a single exposure and outcome. Disaggregating age groups and considering multiple dimensions of the socioeconomic environment is needed to improve understanding of mechanistic pathways and inform timely interventions.

**Table 1.**
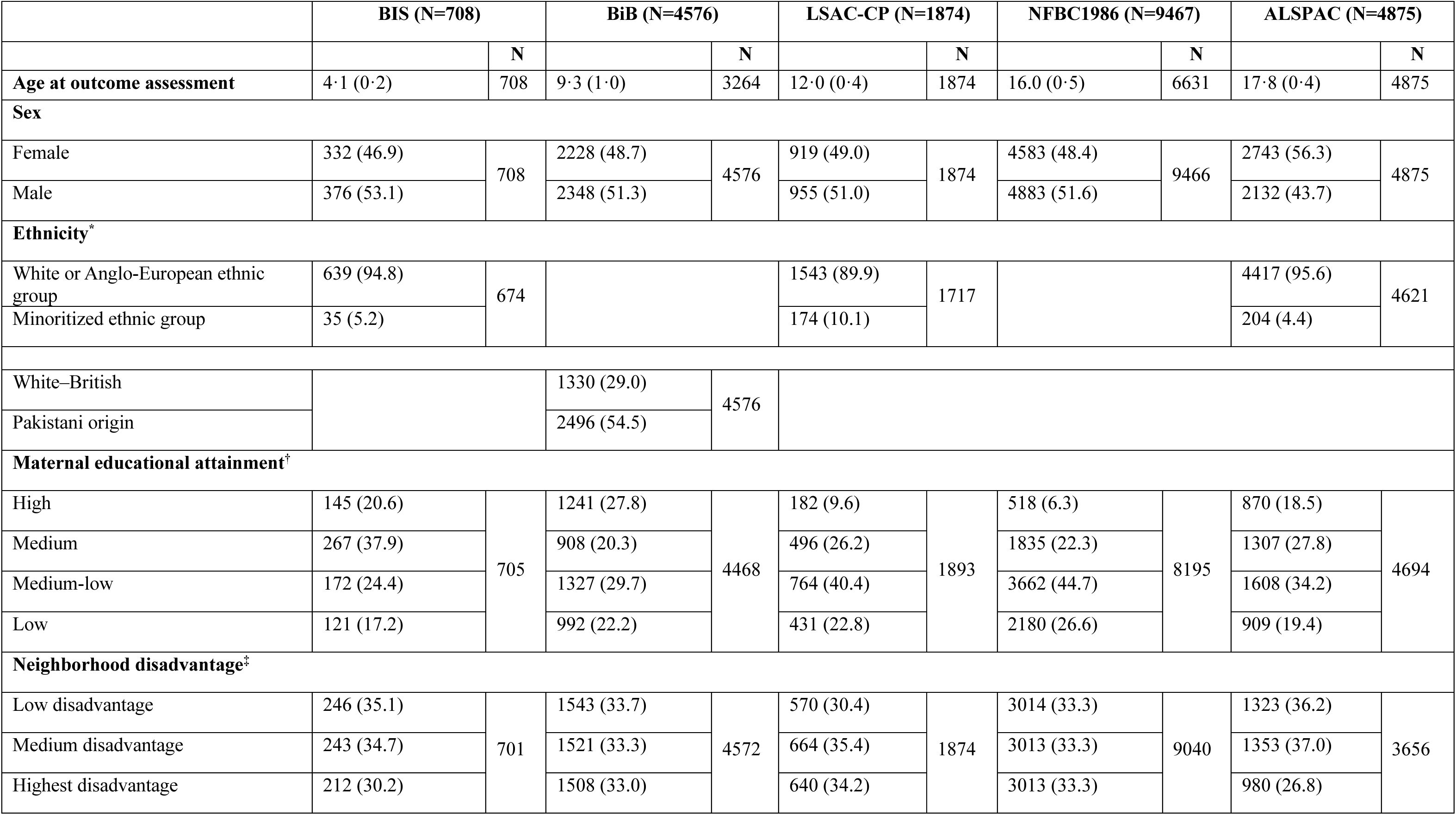
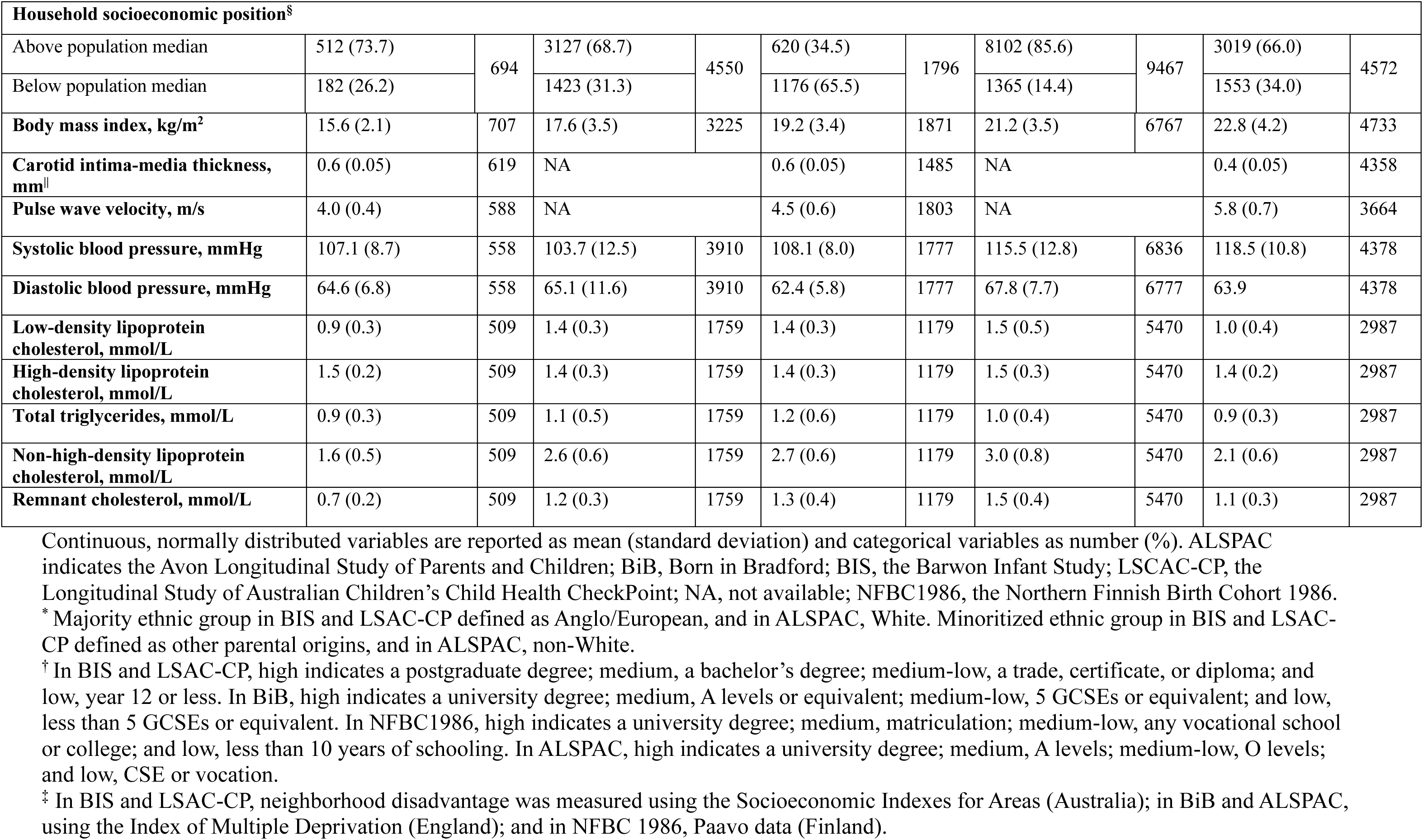
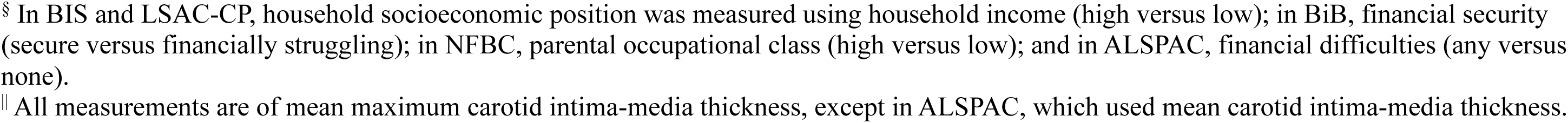
Cohort characteristics.

In this study, we investigated the extent to which measures of neighborhood disadvantage, household SEP, and maternal education in early life affect several vascular and lipid measures of CVH across childhood and adolescence. As overweight and obesity contribute to CVD risk,^13^ we also investigated the extent to which body mass index (BMI) explains these effects, and thus, its utility as a potential intervention target. We present data from five population-derived birth cohort studies from Australia, Finland, and the United Kingdom, with participants aged 4-18 years.

## Methods

### Study Cohorts

We used data from five population-derived birth cohorts: the Barwon Infant Study (BIS, Australia, mean age at outcome measurement 4.1 years, n=708); Born in Bradford (BiB, United Kingdom, mean age 9.3 years, n=4576); the Longitudinal Study of Australian Children’s Child Health CheckPoint (LSAC-CP, Australia, mean age 12.0 years, n=1874); the Northern Finland Birth Cohort 1986 (NFBC1986, Finland, mean age 16.0 years, n=9467); and the Avon Longitudinal Study of Parents and Children (ALSPAC, United Kingdom, mean age 17.8 years, n=4875). Features of study design, geographical setting, participants, and recruitment have been previously described and are presented in Supplemental Methods S1.^14, 15, 16, 17, 18^ Participants recruited at baseline and who attended a study visit at the chosen wave of data collection were included in this study.

### Measurement of Early Life Socioeconomic Position

Three indicators of SEP, all measured at baseline (pregnancy or infancy) or the earliest available time point, were included in this study: neighborhood disadvantage, household SEP, and maternal education. Measures were harmonized across cohorts such that each indicator had the same number of categories. Data sources and cut-offs were cohort-specific.

Neighborhood disadvantage was measured, where available, using pre-existing indices that rank areas based on the socioeconomic characteristics of inhabitants. In BIS and LSAC-CP, the Socio-Economic Indexes for Areas (SEIFA) Index of Relative Socio-Economic Advantage and Disadvantage (IRSAD) was used.^19^ In BiB and ALSPAC, the English Index of Multiple Deprivation (IMD) was used.^20^ In NFBC1986, an index was derived using Paavo postal code area data collected by Statistics Finland.^21^ Cohort-specific tertiles were derived for each index.

Household SEP was operationalized as a binary measure using absolute values of household income or reports of the family financial situation obtained from questionnaire data. In BIS and LSAC-CP, household income was used, with cut-offs at the value closest to the population median household income at the time of measurement. In BiB, mothers reported on how the family were doing financially. Households were dichotomized as ‘financially secure’ if mothers responded that they were “living comfortably” or “doing alright”, or ‘financially struggling’ if mothers responded that they were “just about getting by”, “finding it quite difficult”, or “finding it very difficult”. A four-category sensitivity measure was also derived according to previous work,^22^ in which the final two responses were collapsed into a single ‘financially insecure’ category. In ALSPAC, mothers reported on their ability to afford accommodation, food, and heating. Households were categorized as experiencing ‘financial difficulties’ if there was difficulty affording any of these items, and ‘no financial difficulties’ if not. In NFBC1986, parental occupational class was used as a proxy and dichotomised as ‘high’ or ‘low’.

Maternal education was measured as the highest qualification obtained. Four categories were derived for each cohort: in BIS and LSAC-CP, less than Year 12, trade or diploma, bachelor’s degree, and postgraduate degree; in BiB, less than 5 General Certificates of Secondary Education (GCSEs) or equivalent, 5 GCSEs or equivalent, A levels or other certification, or a university degree; in NFBC1986, less than 10 years of school, any vocational school or college, matriculation, or a university degree; and in ALSPAC, Certificate of Secondary Education or vocation, O levels, A levels, or university degree.

Further details on the measurement and operationalization of these indicators are presented in Supplemental Methods S2.

### Measurement of Cardiovascular Health Measures

Vascular and lipid CVH measures were taken from the time point of interest for each cohort. Vascular measures, where available, included maximum carotid intima-media thickness (cIMT, measured in mm); aortic- or carotid-femoral pulse wave velocity (PWV, m/s); systolic blood pressure (SBP, mmHg); and diastolic blood pressure (DBP, mmHg). Lipid measures included low-density lipoprotein cholesterol (LDL-C, measured in mmol/L); high-density lipoprotein cholesterol (HDL-C, mmol/L); triglycerides (mmol/L); non-HDL-C (mmol/L); and remnant cholesterol (mmol/L). Details on the collection of these measures are available Supplemental Methods S3.

### Measurement of Body Mass Index

At each time point, BMI was measured using the standard calculation of weight (in kilograms) divided by height (in meters) squared. Weight and height were measured either one time or two times (and the average taken), as previously reported.^23, 24, 25, 26, 27^

### Measurement of Covariates and Confounders

Covariates were age at outcome measurement (in years), sex assigned at birth (male or female). Ethnicity was an additional baseline confounder. As recommended, we define ethnicity as a social construct with no biological basis.^28, 29^ In BIS and LSAC-CP, participants were classified as Anglo-European or minoritized ethnic group. In BiB, participants were classified as Pakistani, White-British, or Other (if otherwise classified). In ALSPAC, participants were classified as White or non-White. In NFBC1986, information on ethnicity was not collected. Details on the measurement of intermediate confounders (included in the mediation models) are available in Supplemental Methods S4.

### Statistical Analysis

Data analysis was conducted from May 2024 to June 2025. Each cohort was analyzed separately as we were interested in the timing of effects by age group. Participant characteristics were summarised. Before analysis, multiple imputation by chained equations was conducted to address missingness in all study variables (Table 1). Further detail is presented in Supplemental Methods S5. The primary analysis used imputed data; estimates using complete-case data were obtained as a sensitivity analysis. A directed acyclic graph (DAG) was developed to hypothesize the causal pathways between exposures, the mediator, outcomes, covariates, and confounders (Supplemental Methods S4).

The effect of each SEP indicator in early life on each CVH measure in childhood/adolescence was estimated using multivariable linear regression. The most socioeconomically advantaged category for each indicator was set as the reference group, and unadjusted and adjusted (for age, sex, and ethnicity) models were included. Estimates were presented with 95% CIs. In secondary analyses, a sex-SEP interaction term was included in models to examine effect modification by sex, and sex-stratified analysis conducted to characterize any differences. We considered sex to encompass both biologically sex-linked and socially gendered mechanisms.^30^ We could not examine the effects of gender alone as it was not consistently measured in all the cohorts. Effect modification by ethnicity was investigated in BiB to estimate any differential effects of SEP in Pakistani and White-British children. In BiB, additional sensitivity analysis was conducted using a four-category financial security measure as previously used.^22^

Mediation analysis using an interventional effects approach (defined by mapping to a hypothetical randomized trial)^31^ was conducted to evaluate the mediating role of BMI and evaluate hypothetical interventions. This method is based on a Monte Carlo simulation-based g-computation approach and can also account for exposure–induced mediator–outcome (intermediate) confounding (Figure 1).^32, 33^ The total effect and the interventional indirect effect through BMI were estimated. This indirect effect could be considered the extent to which a hypothetical intervention that shifts the distribution of BMI from that in a lower (exposed) SEP group to be the same as that in the reference (unexposed) SEP group would reduce differences in the CVH measures. We present these relative differences as percentages (the proportion of the total causal effect that is the interventional indirect effect). Further detail on the mediation analysis is available in Supplemental Methods S5.

All statistical analyses were performed in R.4.1.2.^34^

## Results

This study included a total of 21,500 participants aged 4-18 years: in BIS, 708 children (mean [SD] age, 4.1 [0.2] years; 332 girls [46.9%] and 376 boys [53.1%]); in BiB, 4576 children (mean [SD] age, 9.3 [1.0] years; 2228 girls [48.7%] and 2348 boys [51.3%]); in LSAC-CP, 1874 children (mean [SD] age, 12.0 [0.4] years; 919 girls [49.0%] and 955 boys [51.0%]); in NFBC1986, 9467 adolescents (mean [SD] age, 16.0 [0.5] years; 4583 girls [48.4%] and 4883 boys [51.6%]); and in ALSPAC, 4875 adolescents (mean [SD] age, 17.8 [0.4] years; 2743 girls [56.3%] and 2132 boys [43.7%]) (Table 1).

### Neighborhood Disadvantage and Cardiovascular Health

In LSAC-CP, NFBC1986, and ALSPAC, children and adolescents in neighborhoods with greater disadvantage had worse CVH in multiple measures. In LSAC-CP, children in the most disadvantaged neighborhoods had higher cIMT (β = 0.008 mm; 95% CI, 0.002-0.013; *P*=0.01), PWV (β = 0.10 m/s; 95% CI, 0.03-0.16; *P*=0.003), SBP (β = 1.5 mmHg; 95% CI, 0.6-2.4; *P*=0.001), and triglycerides (β = 0.1 mmol/L; 95% CI, 0.1-0.2; *P=*0.004), and lower HDL-C (β = −0.06 mmol/L; 95% CI, −0.10 to −0.02; *P=*0.002), than those in the most advantaged neighborhoods (Figure 1C). Children in medium disadvantage neighborhoods also had higher PWV (β = 0.1 m/s; 95% CI, 0.0-0.1; *P=*0.03) and triglycerides (β = 0.1 mmol/L; 95% CI, 0.02-0.2; *P=*0.02) than those in the most advantaged neighborhoods. In NFBC1986, adolescents in the most disadvantaged neighborhoods had higher non-HDL-C (β = 0.07 mmol/L; 95% CI, 0.02-0.12; *P=*0.01) and remnant cholesterol (β = 0.027 mmol/L; 95% CI, 0.005-0.049; *P*=0.02) than those in the most advantaged neighborhoods (Figure 1D). Adolescents in medium disadvantaged neighborhoods also had higher non-HDL-C (β = 0.1 mmol/L; 95% CI, 0.0-0.1; *P=*0.04) than those in the most advantaged neighborhoods. In ALSPAC, adolescents in the most disadvantaged neighborhoods had higher SBP (β = 0.9 mmHg; 95% CI, 0.1-1.8; *P*=0.03) and DBP (β = 0.8 mmHg; 95% CI, 0.2-1.4, *P*=0.005]) than those in the most advantaged neighborhoods (Figure 1E).

In the younger children in BIS and BiB, results were less consistent. BIS children in medium disadvantage neighborhoods had lower SBP (β = −2.0 mmHg, 95% CI, −3.8 to −0.3, *P=*0.02) and DBP (β = −1.7 mmHg; 95% CI, −3.1 to −0.3; *P=*0.01) than those in the most advantaged neighborhoods (Figure 1A). In BiB, estimates were imprecise, and confidence intervals included both meaningful benefit and harm (Figure 1B).

Subgroup analysis showed that girls in the most disadvantaged neighborhoods in NFBC1986 may have higher triglycerides (β = 0.1 mmol/L; 95% CI, −0.0-0.1; *P=*0.07), but not boys (β = 0.02; 95% CI, −0.05-0.07; *P=*0.40) (difference: β = −0.1 mmol/L; 95% CI, −0.1-0.0; *P=*0.06) (Tables S1, S2, and S3).

Mediation analysis indicated that BMI explained some of the relationship between neighborhood disadvantage and CVH in the older cohorts. In LSAC-CP, a (hypothetical) intervention to shift the distribution of BMI in children from the most disadvantaged neighborhoods to the levels of those from the least disadvantaged neighborhoods would lead to a 20% reduction in PWV differences (interventional indirect effect [IIE] = 0.02 m/s; 95% CI, 0.0-0.04; *P=*0.008), a 53% reduction in SBP differences (IIE = 0.8 mmHg; 95% CI, 0.4-1.2; *P=*<0.001), and a 15% reduction in HDL-C differences (IIE = −0.009 mmol/L; 95% CI, - 0.02-0.0; *P=*0.03) (Tables S4 and S5). Shifting the distribution of BMI in children from medium disadvantage neighborhoods to the levels of those from the most advantaged neighborhoods would also lead to a 43% reduction in PWV differences (IIE = 0.03 m/s; 95% CI, 0.006-0.05; *P=*0.01) (Table S4). In ALSPAC, shifting the distribution of BMI in adolescents from the most disadvantaged neighborhoods to the levels of those in the least disadvantaged neighborhoods would lead to a complete reduction in SBP differences (IIE = 1.0 mmHg; 95% CI, 0.6-1.3; *P=*<0.001) and a 75% reduction in DBP differences (IIE = 0.6 mmHg; 95% CI, 0.4-0.8; *P*=0.005) (Table S6). Confidence intervals were wide in BIS, BiB, and NFBC1986 (Tables S7, S8, S9, S10, S11, and S12).

**Figure 1.**
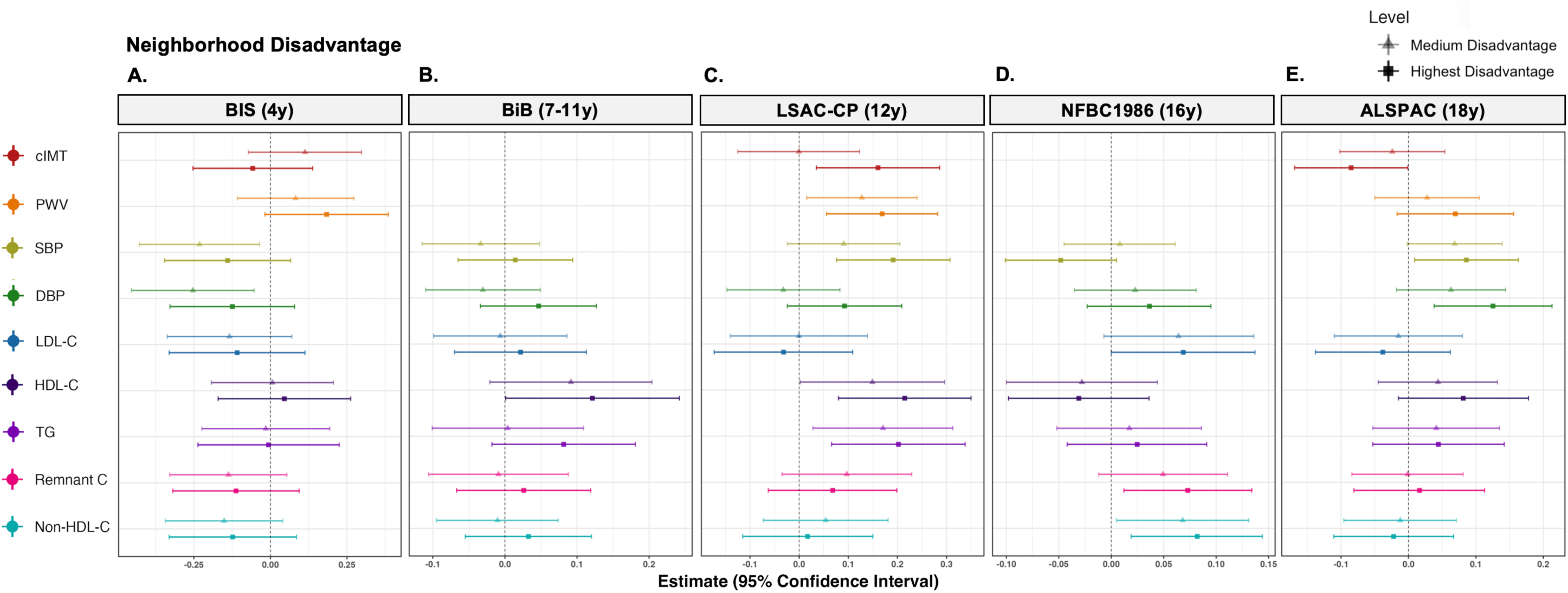
Effects of neighborhood disadvantage in early life (pregnancy or infancy) on CVH measures in childhood and adolescence. Estimates (triangles and squares) are the mean difference in the outcome measure in standard deviation units compared to the reference group (low neighborhood disadvantage, indicated by the dotted vertical line), adjusted for age, sex, and ethnicity. Horizontal bars indicate 95% confidence intervals of estimates. HDL-C has been reverse-coded. ALSPAC indicates the Avon Longitudinal Study of Parents and Children; BiB, Born in Bradford; BIS, the Barwon Infant Study; cIMT, carotid intima-media thickness; DBP, diastolic blood pressure; HDL-C, high-density lipoprotein cholesterol; LDL-C, low-density lipoprotein cholesterol; LSAC-CP, the Longitudinal Study of Australian Children’s Child Health CheckPoint; NFBC1986, the Northern Finland Birth Cohort 1986; Non-HDL-C, non-high-density lipoprotein cholesterol; PWV, pulse wave velocity; Remnant C, remnant cholesterol; SBP, systolic blood pressure; TG, total triglycerides.

### Household Socioeconomic Position and Cardiovascular Health

In LSAC-CP, NFBC1986, and ALSPAC, children and adolescents in lower-SEP households generally had worse CVH measures. LSAC-CP children in lower-income households had higher SBP (β = 0.9 mmHg; 95% CI, 0.1-1.7; *P=*0.02), lower HDL-C (β = −0.04 mmol/L; 95% CI, −0.08 to −0.01; *P*=0.01), and higher triglycerides (β = 0.1 mmol/L; 95% CI, 0.0-0.2; *P*=0.03) compared to their more advantaged counterparts (Figure 2C). Similarly, NFBC1986 adolescents with parents with lower parental occupational class had higher SBP (β = 2.0 mmHg; 95% CI, 1.2-2.8; *P=*<0.001), DBP (β = 0.8 mmHg; 95% CI, 0.2-1.3; *P*=0.005), and non-HDL-C (β = 0.1 mmol/L; 95% CI, 0.0-0.1; *P*=0.03) (Figure 2D). ALSPAC adolescents in households experiencing financial difficulties also had higher SBP (β = 0.7; mmHg; 95% CI, 0.1-1.3; *P=*0.02) (Figure 2E).

No clear evidence of effects and wide confidence intervals were observed in BIS, but in BiB, children in families struggling financially had lower DBP (β = −0.8 mm; 95% CI, −1.6 to −0.1; *P*=0.03) (Figure 2B).

In sex-stratified models, BIS, boys in lower income households had higher cIMT (β = 0.014 mm; 95% CI, 0.002-0.027; *P*=0.03]), but not girls (β = −0.01 mm; 95% CI, −0.02-0.01; *P*=0.31), difference (β = 0.019 mm; 95% CI, 0.003-0.036; *P*=0.02) (Tables S13, S14, and S15). Sensitivity analysis in BiB showed that specifically children in families ‘just about getting by’ had lower DBP than those ‘living comfortably’ (β = −1.0 mmHg; 95% CI, −2.0 to 0.0; *P*=0.05) (Table S16).

There was some evidence of mediation by BMI. A (hypothetical) intervention in LSAC-CP to shift the distribution of BMI in lower-income household children to be the same as those in higher-income households would reduce 78% of SBP differences (interventional indirect effect [IIE] = 0.7 mmHg; 95% CI, 0.3-1.0; *P*=<0.001), 23% of HDL-C differences (IIE = - 0.009 mmol/L; 95% CI, −0.020 to −0.002; *P*=0.02), and 22% of triglyceride differences (IIE = 0.02 mmol/L; 95% CI, 0.002-0.04; *P=*0.03) (Tables S4 and S5). Shifting the distribution of BMI in ALSPAC adolescents in families experiencing financial difficulties to be the same as those with none would reduce 57% of SBP differences (IIE = 0.4 mmHg; 95% CI, 0.2-0.7; *P*=<0.001) (Table S6). There was no evidence of mediation in BIS, BiB, and NFBC1986, as confidence intervals were wide (Supplementary Tables S7, S8, S9, S10, S11, and S12).

**Figure 2.**
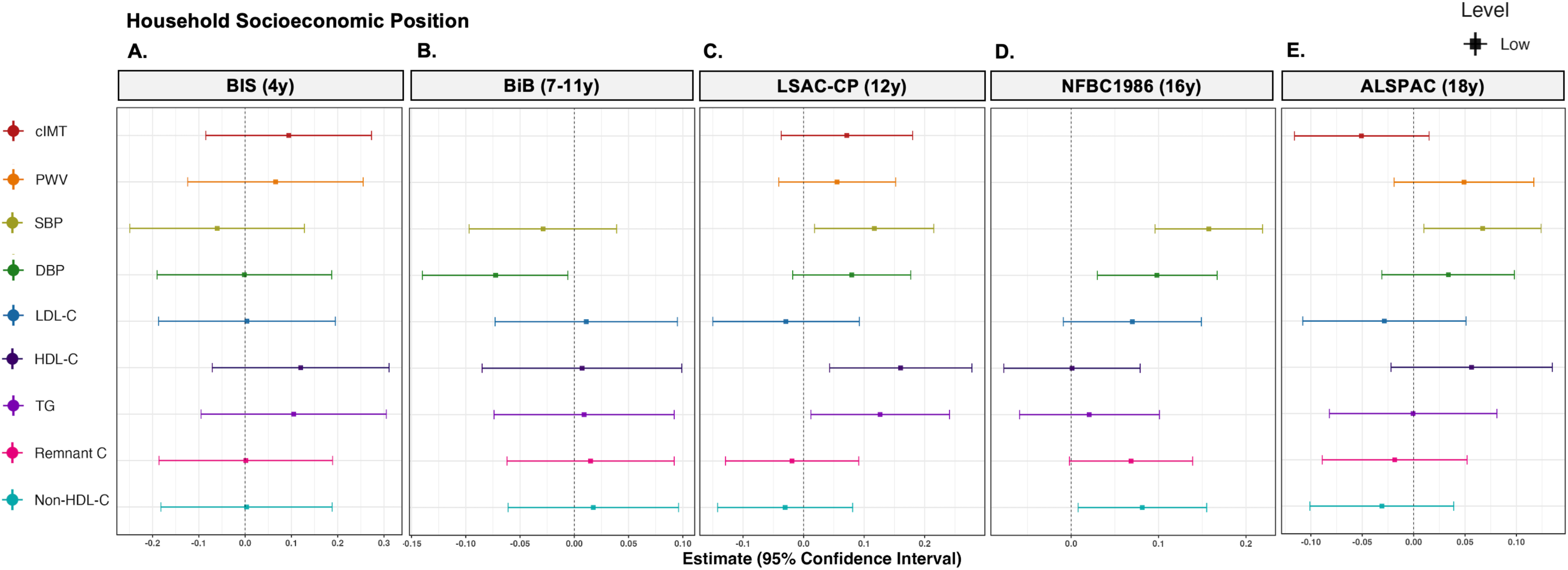
Longitudinal effects of household socioeconomic position (SEP) in early life (pregnancy or infancy) on CVH measures in childhood and adolescence. Squares are the point estimates of the mean difference in the outcome measure in standard deviation units in low household SEP groups compared to the reference group (high household SEP), indicated by the dotted vertical line, adjusted for age, sex, and ethnicity. Household SEP was measured in each cohort as follows: BIS, household income (reference group: higher household income); BiB, financial security (reference group: financially secure); LSAC-CP, household income (reference group: higher household income); NFBC1986, parental occupational class (reference group: high parental occupational class); and ALSPAC, financial difficulties (reference group: no financial difficulties). Horizontal bars indicate 95% confidence intervals of estimates. HDL-C has been reverse-coded. ALSPAC indicates the Avon Longitudinal Study of Parents and Children; BiB, Born in Bradford; BIS, the Barwon Infant Study; cIMT, carotid intima-media thickness; DBP, diastolic blood pressure; HDL-C, high-density lipoprotein cholesterol; LDL-C, low-density lipoprotein cholesterol; LSAC-CP, the Longitudinal Study of Australian Children’s Child Health CheckPoint; NFBC1986, the Northern Finland Birth Cohort 1986; Non-HDL-C, non-high-density lipoprotein cholesterol; PWV, pulse wave velocity; Remnant C, remnant cholesterol; SBP, systolic blood pressure; TG, total triglycerides.

### Maternal Education and Cardiovascular Health

In LSAC-CP, NFBC1986, and ALSPAC, children and adolescents whose mothers had lower education levels had several worse CVH measures. LSAC-CP children whose mothers had less than a postgraduate degree had higher SBP (less than Year 12: β = 2.0 mmHg; 95% CI, 0.6-3.4; *P=*<0.001; trade/diploma/certificate: β = 2.4 mmHg; 95% CI, 1.1-3.7; *P=*0.004; bachelor’s degree: β = 1.4 mmHg; 95% CI, 0.0-2.8; *P=*0.05) and DBP (less than Year 12: β = 1.4 mmHg; 95% CI, 0.3-2.4; *P=*0.009; trade/diploma/certificate: β = 1.6 mmHg; 95% CI, 0.7-2.6; *P*=0.005; bachelor’s degree: β = 1.4 mmHg; 95% CI, 0.4-2.4; *P=*0.005) (Figure 3C). In NFBC1986, adolescents with mothers with less than 10 years of schooling had higher triglycerides than those whose mothers had a university degree (β = 0.07 mmol/L; 95% CI, 0.0-0.1; *P*=0.04) (Figure 3D). Finally, in ALSPAC, adolescents with mothers with a vocational qualification/CSE had higher PWV (β = 0.08 m/s; 95% CI, 0.01-0.2; *P*=0.02), SBP (β = 2.3 mmHg; 95% CI, 1.4-3.3; *P=<*0.001]), and DBP (β = 1.7 mmHg; 95% CI, 1.1-2.3; *P=*<0.001) than those whose mothers had a university degree. Adolescents whose mothers had completed O levels also had higher SBP (β = 1.2 mmHg; 95% CI, 0.4-2.1; *P=*0.004) and DBP (β = 1.0 mmHg; 95% CI, 0.4-1.5; *P=*0.001). The results showed that ALSPAC adolescents whose mothers had a vocational qualification/CSE had lower cIMT (β = −0.008 mm; 95% CI, −0.013 to −0.002; *P=*0.004) (Figure 3E). Wide confidence intervals were present in BiB.

In BIS, subgroup analysis indicated that girls whose mothers had completed year 12 or less had lower remnant cholesterol (β = 2.0 mmol/L; 95% CI, 0.6-3.4; *P=*0.01) and lower non-HDL-C (β = 2.0 mmHg; 95% CI, 0.6-3.4; *P=*0.01); difference with boys 0.1 mmol/L (95% CI, 0.0-0.2; *P=*0.10) and 0.3 mmol/L (95% CI, 0.0-0.6; *P=*0.04) respectively (Tables S13, S14, and S15). In the ALSPAC adolescents, the above effect of maternal education on cIMT was specific to girls (β = −0.011 mm; 95% CI, −0.018 to −0.005; *P*=0.003; difference with boys: β = 0.009 mm; 95% CI, −0.002-0.019; *P*=0.09) (Tables S17, S18, and S19). The effect of having a mother with a Vocational/CSE-level education on SBP and DBP was also stronger for girls (e.g., DBP in girls: β = 2.3 mmHg; 95% CI, 1.4-3.2; *P*=<0.001; DBP in boys: β = 0.9 mmHg; 95% CI, −0.1-1.8; *P*=0.07; difference: β = −1.4 m/s; 95% CI, −2.6 to - 0.1; *P*=0.03) (Tables S17, S18, and S19).

There was evidence for mediation in part by BMI for some of these effects. In LSAC-CP, a (hypothetical) intervention to shift the distribution of BMI in children with mothers who had completed Year 12 or less to be the same as those whose mothers had completed a postgraduate degree would reduce 42% of SBP differences (interventional indirect effect [IIE] = 0.8 mmHg; 95% CI, 0.2-1.4; *P*=0.01) and 21% of DBP differences (IIE = 0.3 mmHg; 95% CI, 0.0-0.6; *P*=0.04). Similarly, shifting the distribution of BMI in children whose mothers had a trade/diploma/certificate to be the same as in those whose mothers had a postgraduate degree would reduce 42% of SBP differences (IIE = 1.0 mmHg; 95% CI, 0.4-1.5; *P*=0.001) and 12% of DBP (IIE = 0.2 mmHg; 95% CI, 0.0-0.4; *P*=0.02) (Table S4).

Shifting the distribution of BMI in ALSPAC adolescents with mothers with a Vocation/CSE to be the same as those with mothers with a university degree would reduce SBP differences by 48% (IIE = 1.1 mmHg; 95% CI, 0.8-1.5; *P*=<0.001) and DBP differences by 41% (IIE = 0.7 mmHg; 95% CI, 0.5-1.0; *P*=<0.001). There was a similar if not more marked effect of shifting the distribution of BMI in adolescents with mothers with O levels to be the same as those with mothers with a university degree: SBP differences would be reduced by 100% (IIE = 0.8 mmHg; 95% CI, 0.5-1.1; *P*=<0.001) and DBP differences by 50% (IIE = 0.5 mmHg; 95% CI, 0.3-0.7; *P*=<0.001) (Table S6). Confidence intervals were wide in BIS, BiB, and NFBC1986 (Tables S7, S8, S9, S10, S11, and S12).

Estimates using complete case data were comparable to those from the primary analyses (Tables S20, S21, S22, S23, and S24). In ethnicity-stratified analyses in BiB, confidence intervals were wide and there was little evidence of a difference in the effects of SEP indicators on cardiovascular measures in Pakistani versus White-British children (Tables S25, S26, and S27).

**Figure 3.**
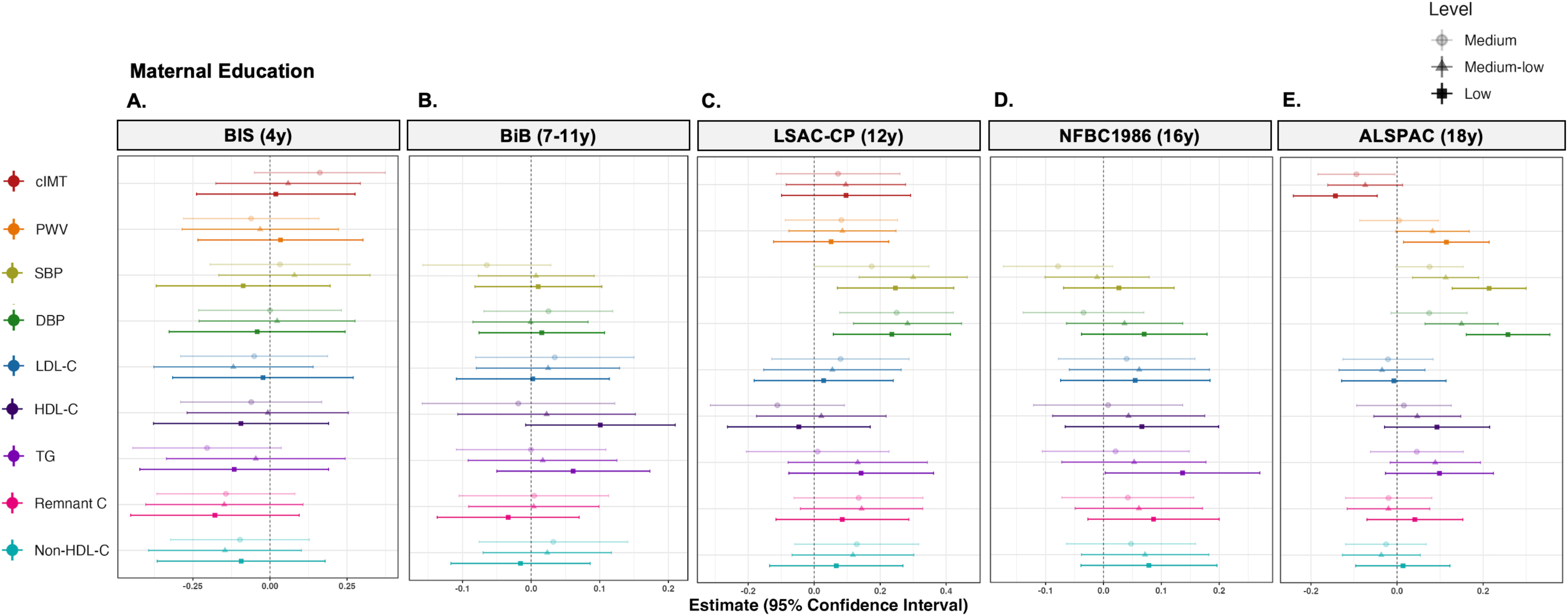
Longitudinal effects of maternal education in early life (pregnancy or infancy) on CVH measures in childhood and adolescence. Estimates (circles, triangles, and squares) are the mean difference in the outcome measure in standard deviation units in lower maternal education groups compared to the reference group, indicated by the dotted vertical line, adjusted for age, sex, and ethnicity. In BIS, the reference group was a postgraduate degree; medium, a bachelor’s degree; medium-low, a trade, certificate, or diploma; and low, year 12 or less. In BiB, the reference group was a university degree; medium, A levels or equivalent; medium-low, 5 GCSEs or equivalent; and low, less than 5 GCSEs or equivalent. In LSAC-CP, the reference group was a postgraduate degree; medium, a bachelor’s degree; medium-low, a trade, certificate, or diploma; and low, year 12 or less. In NFBC1986, the reference group was a university degree; medium, matriculation; medium-low, any vocational school or college; and low, less than 10 years of schooling. In ALSPAC, the reference group was a university degree; medium, A levels; medium-low, O levels; and low, CSE or vocation. Horizontal bars indicate 95% confidence intervals of estimates. HDL-C has been reverse-coded. ALSPAC indicates the Avon Longitudinal Study of Parents and Children; BiB, Born in Bradford; BIS, the Barwon Infant Study; cIMT, carotid intima-media thickness; DBP, diastolic blood pressure; HDL-C, high-density lipoprotein cholesterol; LDL-C, low-density lipoprotein cholesterol; LSAC-CP, the Longitudinal Study of Australian Children’s Child Health CheckPoint; NFBC1986, the Northern Finland Birth Cohort 1986; Non-HDL-C, non-high-density lipoprotein cholesterol; PWV, pulse wave velocity; Remnant C, remnant cholesterol; SBP, systolic blood pressure; TG, total triglycerides.

## Discussion

This multicohort study of 21,500 participants from five longitudinal birth cohorts aged 4-18 years suggests that the adverse effects of lower early-life SEP on later CVH are established from mid-childhood. These effects were consistent across different population contexts in Australia, Finland, and the UK, and for a range of vascular and lipid measures. Effects were observed for SEP at the neighborhood, household, and individual level. Mediation analysis indicated that between 15% and 100% of relative differences in CVH measures could be reduced by a hypothetical intervention on BMI.

We report the largest multicohort study to date, in which we disaggregated age groups to investigate the effects of individual, household and neighborhood socioeconomic indicators on multiple measures of CVH across several large, population-based cohorts. Our findings align with previous studies, which have shown that lower SEP is associated with higher cIMT,^35^ BP,^11, 36, 37^ and an adverse lipid profile^38, 39^ in children and adolescents. We provide further evidence that socioeconomic inequalities in the risk and pathogenesis of CVD may originate early in life, well before the presentation of clinical disease.

In this study, we did not use clinical cut-offs for the outcome measures, which currently only exist for BP and lipid levels in pediatric populations.^40, 41^ Rather than identify which children were at risk, we aimed to explore the social production of inequalities in early CVH and quantify differences between socially stratified groups. This is particularly important given recent evidence indicating that higher CVD risk in childhood is associated with future CVD events, independent of adult CVD risk, per unit increase in mean risk score.^8^ When clinically informed risk factor categories were used, there was a higher risk of adult CVD for not only the high-risk but high-normal and high-acceptable groups. The differences observed in our study may therefore contribute to inequalities in clinical CVD later in life, even if socioeconomic conditions improve into adulthood.^42^ This is supported by the large body of evidence on the importance of maintaining optimal CVH throughout the life course.^43, 44^

The effects observed in this study were inconsistent at younger ages (4-11 years), with estimates in both positive and negative directions. It is difficult to ascertain whether these inconsistencies are attributable to cohort-specific differences or to age. For example, BIS is a relatively affluent cohort, with a majority of English-speaking, Anglo/European families.^14^ The mechanisms that explain effects in BIS may be different to those that operate in more heterogeneous contexts. In contrast, BiB is located in a particularly deprived region of the UK.^15^ Previous research at similarly young ages is sparse. Lower SEP has been associated with higher BP in two studies of children aged 5-6 years,^11, 45^ but not in another multicohort study of those aged 4-5 years.^46^ As BP tracks from early childhood across the life course,^47^ further work using repeated measures in the same individuals may identify how early these effects emerge, and whether they are context-specific. Some cohorts have begun to collect these data;^14, 48^ future longitudinal studies should consider more frequent biological measurements during this critical developmental period.

Variation in the effects of SEP were also observed for cIMT across the cohorts in which it was measured, from early childhood to late adolescence. Boys in lower-income households in BIS and children in more disadvantaged neighborhoods in LSAC-CP had higher maximum cIMT compared to their more advantaged counterparts. In ALSPAC, adolescents with mothers with the lowest educational qualifications had lower mean cIMT. Maximum cIMT in BIS and LSAC-CP refers to the mean of the thickest measurement across several frames of the chosen segment of the carotid artery,^35^ whereas mean cIMT in ALSPAC refers to the mean of measurements across the entire segment.^49^ Some evidence suggests that maximum cIMT is a better predictor of future CVD events than mean cIMT.^50^ Indeed, previous work in LSAC-CP showed that lower composite SEP was associated with higher maximum but not mean cIMT.^35^ These differences in measurement preclude the effects across the cohorts from being directly comparable, particularly at younger ages when inter-individual variations may be too small to compare without standardized methodology. In adults, maximum cIMT is a stronger predictor of myocardial infarction than mean cIMT.^50^ The extent to which cIMT in adults reflects subclinical atherosclerosis has also been questioned, with the phenotype likely reflecting adaptive processes more than atherosclerotic burden, especially in childhood.^51, 52^ Previous work in ALSPAC has shown that cIMT at 17-18 years is associated with fat-free mass and BP, but not metabolic or lifestyle CVD risk factors.^53^ More consistent measurement of cIMT with standardized protocols is needed to identify generalizable effects.^50^

Our study indicates that early-life socioeconomic exposures at the neighborhood, household, and individual level are important for CVH in later childhood and adolescence. Each exposure was associated with both vascular and lipid measures from mid-childhood. There are likely myriad explanatory mechanisms by which these socioeconomic conditions are reflected in cardiovascular phenotypes. These may include direct pathways, where an exposure results in a direct physiological process, or indirect pathways, where exposures are internalized via neuroendocrine responses.^54^ For example, children in lower-SEP families may have limited access to a healthy diet or opportunities for physical activity (direct);^55, 56^ be exposed to greater air and water pollution (direct);^57^ or experience a stressful family emotional environment due to compromised parental capacity or a reduced sense of belonging due to poor neighborhood cohesion (indirect).^58^ We also observed effect modification by sex, particularly for girls. This indicates the potential relevance of sex-linked and gendered mechanisms that may exacerbate the biological effects of socioeconomic disadvantage in girls.^30^ Confidence intervals were wide for models investigating effect modification by ethnicity in BiB. It is plausible that the effects of social cohesion and collectivism in the Pakistani community may protect against the adverse effects of low SEP.^59, 60^ Our findings highlight the importance of precise social and structural interventions, such as those that improve neighborhood conditions; provide families with the material and social resources needed for their children to thrive; and support mothers with paid parental leave, free child-care, and opportunities for further education.

Mediation analysis indicated that a hypothetical intervention shifting the distribution of BMI in children and adolescents with lower SEP to be the same as their more advantaged counterparts would reduce differences in CVH in LSAC-CP and ALSPAC by between 15% to almost completely. Obesity is a major CVD risk factor,^13^ and BMI tends to track from childhood to adulthood, with approximately 50% of children with obesity also having obesity as adolescents, of which around 80% have obesity as adults.^61, 62^ Recent systematic reviews have shown that people with persistent overweight from childhood to adulthood have a higher risk of CVD risk factors and clinical outcomes, but not those with overweight-to-normal trajectories.^63, 64^ These findings underscore the importance of early intervention efforts to alter trajectories of BMI, although BMI-defined thresholds are not optimal indicators of overweight and obesity.^65^ Body mass and body composition reflect both genetic and environmental factors,^66^ with many of the latter being socially structured. Addressing both upstream mechanisms closely related to SEP and downstream factors such as diet and physical activity are likely to be more effective than individual-level interventions alone.^67^

The main strength of this study is its multicohort design, in which we used prospective data to build evidence for the effects of SEP on childhood CVH across various population-contexts. We aimed to consider multiple dimensions of SEP across individual, household and neighborhood levels; similarly, we assessed CVH holistically by multiple vascular and lipid measures. Novel interventional effects methodology^32^ allowed us to investigate the potential efficacy of an intervention target. However, several limitations must be considered. In using cohorts that differ in setting, age-group, methodology, and sources of bias, it is difficult to disentangle the effects of age and population context, and estimates cannot be directly compared. Analyses should be repeated at older ages in our younger cohorts and in other population-derived samples. Likewise, we note that differential effects were observed for the cardiovascular measures in each cohort, where some were patterned by SEP in one cohort but not another. Without repeated measures, it is difficult to ascertain whether null effects were due to age, cohort, or mechanistic reasons. We also note difficulties harmonizing household SEP due to data restrictions, and highlight that the given measures may not wholly or comparably capture household resources. While we show evidence that low SEP in early life has implications for CVH in later childhood and adolescence, we also cannot make inferences about the windows of exposure with most biological consequence. In addition, some intermediate confounders (such as diet and physical activity) were either not available or not collected in sufficient participants to be included in mediation analysis, which may affect the validity of some estimates. Finally, our findings are only generalizable to similar high-income contexts. The burden of CVD falls disproportionately on low- and middle-income countries, where it occurs approximately a decade younger on average than in high-income countries,^1^ but life course mechanisms of risk likely differ considerably.^68^ Granular social and biological data across diverse population contexts is essential to provide general and context-specific evidence that informs precise prevention targets.

## Conclusions

Our findings suggest that low early-life SEP may contribute to adverse CVH measures from mid-childhood onwards, across cohorts in Australia, Finland, and the UK. Intervening on BMI and the obesogenic environment may partly reduce socioeconomic differences in vascular and lipid profiles. Future work should consider further disaggregating the pathways by which neighbourhood disadvantage, household SEP, and maternal education influence CVH, to inform intervention targets and alter adverse risk trajectories.

## Data Availability

Barwon Infant Study (BIS) data are available under restricted access on scientific and ethical grounds by the BIS Steering Committee and provided through a collaborative research agreement. A description of the available data is available on the cohort website: https://barwoninfantstudy.org.au. Born in Bradford (BiB) data are available under restricted access through application on the study website, where information on the available data is also available: https://borninbradford.nhs.uk/research/how-to-access-data/. Data from the Longitudinal Study of Australian Children's Child Health CheckPoint (LSAC-CP) are available under restricted access and may be obtained by request through the Australian Data Archive (https://ada.edu.au/) for researchers in Australia affiliated with an approved research body. Northern Finland Birth Cohort 1986 (NFBC1986) data are available under restricted access through application on the study website (https://www.oulu.fi/en/university/facultiesand-units/faculty-medicine/northern-finland-birth-cohorts-and-arctic-biobank/nfbcaineistopyynto) and provided through a collaborative research agreement with the University of Oulu. The informed consent obtained from ALSPAC (Avon Longitudinal Study of Parents and Children) participants does not allow the data to be made available through any third party maintained public repository. Supporting data are available from ALSPAC on request under the approved proposal number, B4806. Full instructions for applying for data access can be found here: http://www.bristol.ac.uk/alspac/researchers/access/. The ALSPAC study website contains details of all available data, including a data dictionary and variable search tool (http://www.bristol.ac.uk/alspac/researchers/our-data/).

https://www.oulu.fi/en/university/faculties-and-units/faculty-medicine/northern-finland-birth-cohorts-and-arctic-biobank/nfbc-aineistopyynto

https://ada.edu.au/

https://borninbradford.nhs.uk/research/how-to-access-data/

https://barwoninfantstudy.org.au

## Acknowledgements

KG performed the data analysis and visualisation and wrote the original draft. NP, TM, and DB supervised the study and are joint senior authors. SP conducted data analysis for the NFBC1986 cohort. KG and TM accessed and verified the data. TC guided data analysis and methodology development. DB, TM, NP, and KG conceptualized the study. All authors reviewed and edited the manuscript and approved content for submission. The authors are grateful to the participating families and staff of the BIS, BiB, LSAC-CP, NFBC1986, and ALSPAC cohorts.

## Ethical approval

The Human Research and Ethics Committee (reference: 10/24) of Barwon Health gave ethical approval for the Barwon Infant Study. The Health Research Authority Yorkshire and the Humber (Bradford Leeds) Research Ethics Committee (reference: 16/YH/0320) of the National Health Service United Kingdom gave ethical approval for Born in Bradford. The Human Research Ethics Committee of the Royal Children’s Hospital Melbourne (reference: 33225D) and the Ethics Committee of the Australian Institute of Family Studies (reference: 14-26) gave ethical approval for the Longitudinal Study of Australian Children’s Child Health CheckPoint. The Ethical Committee of the Northern Ostrobothnia Hospital District (reference: 108/2017) gave ethical approval for the Northern Finland Birth Cohort 1986. The Ethics and Law Committee of the Avon Longitudinal Study of Parents and Children and the Research Ethics Committees of the Bristol and Weston Health Authority (reference: E1808), Southmead Health Authority (reference: 49/89), Frenchay Health Authority (reference: 90/8), and the North Somerset & South Bristol Authority (reference: 08/H0106/9), gave ethical approval for the Avon Longitudinal Study of Parents and Children.

## Data sharing

Barwon Infant Study (BIS) data are available under restricted access on scientific and ethical grounds by the BIS Steering Committee and provided through a collaborative research agreement. A description of the available data is available on the cohort website: https://barwoninfantstudy.org.au. Born in Bradford (BiB) data are available under restricted access through application on the study website, where information on the available data is also available: https://borninbradford.nhs.uk/research/how-to-access-data/. Data from the Longitudinal Study of Australian Children’s Child Health CheckPoint (LSAC-CP) are available under restricted access and may be obtained by request through the Australian Data Archive (https://ada.edu.au/) for researchers in Australia affiliated with an approved research body. Northern Finland Birth Cohort 1986 (NFBC1986) data are available under restricted access through application on the study website (https://www.oulu.fi/en/university/faculties-and-units/faculty-medicine/northern-finland-birth-cohorts-and-arctic-biobank/nfbc-aineistopyynto) and provided through a collaborative research agreement with the University of Oulu. The informed consent obtained from ALSPAC (Avon Longitudinal Study of Parents and Children) participants does not allow the data to be made available through any third party maintained public repository. Supporting data are available from ALSPAC on request under the approved proposal number, B4806. Full instructions for applying for data access can be found here: http://www.bristol.ac.uk/alspac/researchers/access/. The ALSPAC study website contains details of all available data, including a data dictionary and variable search tool (http://www.bristol.ac.uk/alspac/researchers/our-data/).

## Supplemental Material

Supplemental Methods

Tables S1-S35

References #68-#99

