## Supplemental Material for "Neighborhood, household, and individual socioeconomic position in early life and childhood cardiovascular health measures: an international cross-cohort study"

**Supplemental Methods S1. Cohort descriptions**

*The Barwon Infant Study*

The Barwon Infant Study (BIS) is a longitudinal, population-derived birth cohort study conducted in the Barwon region of Victoria, in south-eastern Australia. Women attending an antenatal appointment at 15 weeks of pregnancy at Geelong Hospital (government-funded) and St John of God Hospital (private) in the Barwon region were invited to participate in BIS. Recruitment began in 2010 and concluded in 2013. Women were eligible to participate if they were residents of the Barwon Statistical Division, no more than 28 weeks pregnant at enrolment, planning to give birth at either of the two hospitals, and expecting to be available for the study period. Women were ineligible if they were not an Australian permanent resident, were not able to complete the questionnaires without an interpreter, did not or were not able to give informed consent, were under 18 years old, already had a child in BIS, planned to privately store their child's cord blood, or had moved out of the Barwon Statistical Division by the time of birth. Infants were ineligible if they were born prior to 32 weeks of gestation, had a serious illness, or had a congenital malformation or disease. The resulting inception cohort comprised 1158 women (33% recruitment rate) and 1074 infants. The cohort was comparable to the Australian population, but participants were more likely to be socioeconomically advantaged and of non-English-speaking background. Data collection included questionnaires and physical, physiological, and clinical measurements. Participants were followed up frequently during their first year of life, and since, there have been reviews at 1.5 years, 2 years, 4 years, and 8-11 years. Ethical approval for the study was obtained from the Barwon Health Human Research and Ethics Committee (reference: 10/24). Informed consent was obtained from the attending parent or guardian at pre-natal recruitment and at the pre-school review. BIS is funded by the National Health and Medical Research Council of Australia, The Shane O'Brien Memorial Asthma Foundation, Our Women's Our Children's fundraising committee, The Cotton On Foundation, and The Jack Brockhoff Foundation.[69]

*Born in Bradford*

Born in Bradford (BiB) is a longitudinal cohort study based in the city of Bradford, in the United Kingdom (UK). All hospital births in Bradford occur at Bradford Royal Infirmary. From 2007 to 2010, women attending an oral glucose tolerance test at 26–28 weeks gestation were invited to participate in BiB. The recruitment rate was over 80%, and 12 453 women with 13 776 pregnancies were in the inception cohort. All infants born to these women were eligible for the study, of which there were 13,818. Around half of the babies born in Bradford have parents of Pakistani origin. This was reflected in the BiB cohort, which had 6900 (50.1%) infants of South Asian ethnicity. Compared to non-recruited participants, the cohort had a lower proportion of younger mothers and a higher proportion of first-time mothers.[15] Data collection included questionnaires to extract detailed demographic and lifestyle information as well as various physical and biological measures. This study uses the main BiB cohort. Participant follow-up has occurred at multiple time points throughout childhood and adolescence. Ethical approval was obtained from the National Health Service Health Research Authority Yorkshire and the Humber (Bradford Leeds) Research Ethics Committee for the community-based family assessments and school-based measures (reference: 16/YH/0320). Informed consent was obtained from parents for themselves and their child and included consent for future linkage to health and education records. BiB is funded by the UK's National Institute for Health Research Collaboration for Applied Health Research and Care and the Programme Grants for Applied Research funding scheme.[70]

*The Longitudinal Study of Australian Children's Child Health CheckPoint*
The Child Health CheckPoint was a cross-sectional wave of Growing Up in Australia: The Longitudinal Study of Australian Children (LSAC). LSAC CheckPoint (LSAC-CP) was focused on physiological, biological, and clinical measures. At inception, LSAC comprised two cohorts: the K cohort (children aged 4-5 years, N=4984 families) and the B cohort (children aged 0-1 years, N=5107 families). Children were recruited from 2004 to 2005 using a national random sampling design, where postcodes were stratified by state and statistical division. Children in these areas were randomly selected using the Medicare (Australia's universal public health insurance) database and only excluded if from remote postcodes, had a sibling already selected or were deceased.[71] Uptake for each cohort was around 50%. CheckPoint occurred between waves 6 and 7 of LSAC using the B cohort, when children were aged 11-12 years, from 2015-2016. Any LSAC B cohort family that had completed a wave 6 home interview was eligible. Ethical approval was obtained from the Royal Children's Hospital Melbourne Human Research Ethics Committee (33225D) and the Australian Institute of Family Studies Ethics Committee (14-26). Informed consent was obtained from the parent or guardian for their own and their child's participation in the study. LSAC-CP is funded by the National Health and Medical Research Council of Australia and several other Australian foundations and research institutions.[16]

*The Northern Finland Birth Cohort 1986*
The Northern Finland Birth Cohort of 1986 (NFBC1986) is a longitudinal cohort study conducted in northern Finland's Oulu and Lapland regions. NFBC1986 covered the populations of Oulu and Lapland using an unselected sample. Pregnant women whose expected due date fell between July 1985 and June 1986 were enrolled at 25 weeks of gestation during a visit to any maternity health centre in Oulu and Lapland. Those who miscarried prior to 24 weeks of gestation, had multiple births, or whose place of birth was not known or was outside the study area were excluded. The inception cohort comprised 9362 mothers and 9479 infants (99% of all infants in the target area). Data collection included demographic information extracted using questionnaires and physical, clinical, biological, and developmental measures. Participants have been followed up at 1 year, 7 years, 8 years, 15-16 years, and 33-35 years. Ethical approval was obtained from the Ethical Committee of the Northern Ostrobothnia Hospital District (reference: 108/2017). Informed consent was obtained from all participants and their parents. Funding was provided by the University of Oulu, the Oulu University and national and European research funding bodies.[72, 73]

*The Avon Longitudinal Study of Parents and Children*
The Avon Longitudinal Study of Parents and Children (ALSPAC) is a transgenerational longitudinal cohort study conducted in the former county of Avon in the UK. All pregnant women with an expected delivery date between April 1991 and December 1992 in the catchment area were eligible to participate. Also eligible were pregnant women migrating into the area. Ineligible were women migrating out of the area who had not completed their third-trimester questionnaire. All infants delivered from eligible pregnancies were themselves eligible. 20,248 pregnancies have been identified as being eligible and the initial number of pregnancies enrolled was 14,541. Of the initial pregnancies, there was a total of 14,676 fetuses, resulting in 14,062 live births and 13,988 children who were alive at 1 year of age. When the oldest children were approximately 7 years of age, an attempt was made to bolster the initial sample with eligible cases who had failed to join the study originally. As a result, when considering variables collected from the age of seven onwards (and potentially abstracted from obstetric notes) there are data available for more than the 14,541 pregnancies mentioned above: The number of new pregnancies not in the initial sample (known as Phase I

enrolment) that are currently represented in the released data and reflecting enrolment status at the age of 24 is 906, resulting in an additional 913 children being enrolled (456, 262 and 195 recruited during Phases II, III and IV respectively). The phases of enrolment are described in more detail in the cohort profile paper and its update (see footnote 5 below). The total sample size for analyses using any data collected after the age of seven is therefore 15,447 pregnancies, resulting in 15,658 fetuses. Of these 14,901 children were alive at 1 year of age.[74] Compared to mothers in Britain, ALSPAC were more socioeconomically advantaged, more likely to be married and more likely to be White.[18] Data collection included demographic information extracted using questionnaires and physical, biological and developmental measures extracted during clinical assessments conducted at various points throughout childhood and adolescence. There have been 68 follow-up time points between birth and 18 years, categorised into 6 phases of data collection: infancy (2 weeks to 2 years), early childhood (2 to 7 years), childhood (7 years), late childhood (7 to 13 years), adolescence (13 to 16 years) and transition to adulthood (16 to 18 years).[74] Ethical approval for the study was obtained from the ALSPAC Ethics and Law Committee and the Local Research Ethics Committees. Informed consent for the use of all data collected was obtained from participants following the recommendations of the ALSPAC Ethics and Law Committee at the time. Participants can contact the study team at any time to retrospectively withdraw consent for their data to be used. Study participation is voluntary and during all data collection sweeps, information was provided on the intended use of data. The completion of a questionnaire, either on paper or online, was considered to be written consent from participants to use their data for research purpose. For the majority of tests undertaken during face to face visits, verbal consent was obtained from participants (both parents and children as appropriate) prior to the start of any data collection. However, some tests required the completion of a written consent form. Biological samples are collected in accordance with the Human Tissue Act (2004). Specific Research Ethics Committee approval is sought for the consenting process at each collection sweep. Written consent, including permission for future use, is obtained from adult participants or from the parents of children as appropriate. Ethical approval for future use is covered by ALSPAC's Research Tissue Bank approval. All historical consents to hold biological samples have been reviewed as part of the Tissue Bank approval process. Participants can contact the study team at any time to retrospectively withdraw consent for use of their samples. The UK Medical Research Council and Wellcome Trust (Grant ref: MR/Z505924/1) and the University of Bristol provide core support for ALSPAC. This publication is the work of the authors and KG will serve as guarantor for the contents of this paper. A comprehensive list of grants funding is available on the ALSPAC website (http://www.bristol.ac.uk/alspac/external/documents/grant acknowledgements.pdf); This research was specifically funded by Wellcome Trust grants 086684, and 076467/Z/05/Z and UK Medical Research Council grants G0800612/86812 and MC_UU_12013/1. For the purpose of Open Access, the author has applied a CC BY public copyright license to any Author Accepted Manuscript version arising from this submission.

**Supplemental Methods S2. Measurement of socioeconomic position**

*Neighbourhood disadvantage*

Neighborhood disadvantage was measured using pre-existing nationally-derived indices of area-level socioeconomic disadvantage and/or advantage in BIS, BiB, LSAC-CP, and ALSPAC. In BIS and LSAC-CP, the Socio-Economic Indexes for Areas (SEIFA) Index of Relative Socio-Economic Advantage and Disadvantage (IRSAD) was used. This is a measure of the relative level of advantage and disadvantage of an area—in this case Statistical Areas Level 1 (SA1s), which have a population of around 200 to 800 inhabitants.[19] In BiB and ALSPAC, the English Index of Multiple Deprivation (IMD) was used. This is a measure of the relative level of deprivation of Lower-layer Super Output Areas, which have an average population of 1500.[75] In NFBC1986, an index was derived using Paavo postal code area statistics collected by Statistics Finland.[76] Paavo contains area-level data (including data on neighbourhood socioeconomic disadvantage) based on the postal areas of the newest system (year 2024). The study index was derived from the following Paavo variables, as previously:[77] aged 18 or over; adults with a basic education; employed; unemployed; households, total; and households living in rental dwellings. The ratio variables were rescaled to a standard distribution of mean 0 and standard deviation 1. The higher the score for each variable, the greater the disadvantage. The SD-scores were weighted according to the population density of the corresponding postal area. These densities were calculated as a ratio of the number of households to postal area's area (km2). The neighbourhood disadvantage index was then derived by obtaining the mean value of the three SD-scores and division into tertiles: the lowest tertile corresponded to the most advantaged areas; the middle tertile corresponded to medium disadvantage; and the highest tertile corresponded to the least advantage/most disadvantage. The tertile cut-offs were as follows: low (-1·6, 0·2); middle (0·2, 0·7); and high (0·7, 6·6]. NFBC 1986 participants were linked to the index by mapping their original residence coordinates to the new postal areas (2024), using Statistics Finland's mapping tool. Participants living abroad were excluded. Data on neighbourhood disadvantage were collected

*Household socioeconomic position*

In BIS and LSAC-CP, household socioeconomic position (SEP) was measured using responses by study mothers to survey questions on the gross level of income the household receives annually. Responses were categorical and provided within a range, so original variables were dichotomised closest to the population median household income at the time of data collection. In BIS, this was AUD 75,000 (the population median being AUD 64,000 in 2011),[78] and in LSAC-CP this was AUD 78,000 (the population median being AUD 67,808 in 2003).[79] In BiB and ALSPAC, a measure of financial difficulty or financial security was used (as described in the main material) according to mother responses to questionnaires. In NFBC1986, parental occupational class was used, which largely corresponded to income at the time of data collection.[80] The highest occupational class of the mother or father was used as the overall highest parental occupational class. Data on household SEP were collected in pregnancy in BIS, BiB, and ALSPAC, and in infancy in LSAC-CP and NFBC1986.

*Maternal education*

Maternal education was obtained using mother responses to survey questions in all cohorts as described in the main material. These data were collected in pregnancy in BIS, BiB, and ALSPAC, and in infancy in LSAC-CP and NFBC1986.

**Supplemental Methods S3. Measurement of cardiovascular health measures**

*Carotid intima-media thickness*

In BIS, maximum carotid intima-media thickness (cIMT) was measured in the right common carotid artery at end-diastole. Three cineloops of five or more cardiac cycles were captured using a GE Vivid I ultrasound machine with 10 MHz linear transducer (GE Healthcare, New South Wales, Australia). Measurements were analyzed using edge-detection software (Coast) programmed in Matlab (R2022b, The Mathworks Inc, Natick, MA). Maximum cIMT measurements from three to five images of the selected segment were averaged for the final measurement.[81, 82] In LSAC-CP, maximum cIMT was also measured in the right common carotid artery at end-diastole. The best cineloop of five to six cardiac cycles captured using a GE Vivid I ultrasound machine with 10 MHz linear transducer was taken. This was analyzed using a Carotid Analyzer (Medical Imaging Applications, Coralville, Iowa, USA). The mean maximum cIMT of three to five images was calculated.[83, 84] In ALSPAC, mean cIMT was measured in the right and left common carotid artery at end-diastole. Ten-second cineloops across three cardiac cycles were captured using a 12-MHz linear transducer (Vivid7, GE Medical, Chicago, Illinois). Measurements were analyzed using Carotid Analyzer (Medical Imaging Applications, Coralville, Iowa, USA). Three images from both the right and left carotid artery were taken and averaged for the final measurement. [85, 86]

*Pulse wave velocity*

In BIS, carotid-femoral pulse wave velocity (PWV) was measured using the SphygmoCor XCEL device (AtCor Medical Pty, New South Wales, Australia). Three to five measurements were taken after a five-minute rest period, which were averaged for the final measurement.[81] In LSAC-CP, carotid-femoral PWV was also measured using the SphygmoCor XCEL device after a few minutes of rest. The mean of two valid measurements was used for the final measure.[87] In ALSPAC, carotid-femoral PWV was measured using a Vicorder device (Skidmore Medical). Three measurements were taken the average used for the final measurement.[88]

*Blood pressure*

In BIS, systolic and diastolic blood pressure (BP) were measured using the SphygmoCor XCEL device at the brachial artery. Three to five measurements were taken after a five-minute rest period, which were averaged for the final values.[81] In Born in Bradford, BP was measured at the brachial artery using an Omron electronic blood pressure monitor 705-CPII (OMRON Matsusaka Co. Ltd, Japan). The average of two measurements was used as the final measure.[89] In LSAC-CP, BP was measured using the SphygmoCor XCEL device. After a few minutes of rest, several measures were taken at the brachial artery, and the mean of two valid measurements was used for the final measure.[87] In NFBC1986, BP was measured using an Omron 705CP monitor after 15 minutes of rest. The average of two measurements was used as the final measure.[90] In ALSPAC, BP was measured using an Omron 705-IT monitor. The final two of three measurements was taken and averaged for the final measure.[88]

*Metabolic measures*

Low-density lipoprotein cholesterol (LDL-C), high-density lipoprotein cholesterol (HDL-C), serum total cholesterol, and serum total triglycerides were measured using a nuclear magnetic resonance-based metabolomics platform (Nightingale Health, Helsinki, Finland) in all cohorts using processed blood samples.[91, 92, 93, 94, 95] Non-HDL-C and remnant cholesterol were calculated for each cohort using the following formulae: non-HDL-C = total cholesterol – HDL-C; remnant cholesterol = total cholesterol – HDL-C – LDL-C.

**Supplemental Methods S4. Mediation analysis**

Mediation analysis to investigate the extent to which BMI explained the effects of SEP on cardiovascular health was conducted using an interventional effects approach. This method is useful when attempting to evaluate hypothetical mediator interventions and is appropriate in populations and settings where no well-defined mediator interventions—in this case, weight-loss or lifestyle programs—have been implemented, thereby helping to inform potential intervention targets.[32] Here, estimation of interventional effects was conducted using the Monte Carlo simulation-based g-computation method.[96]

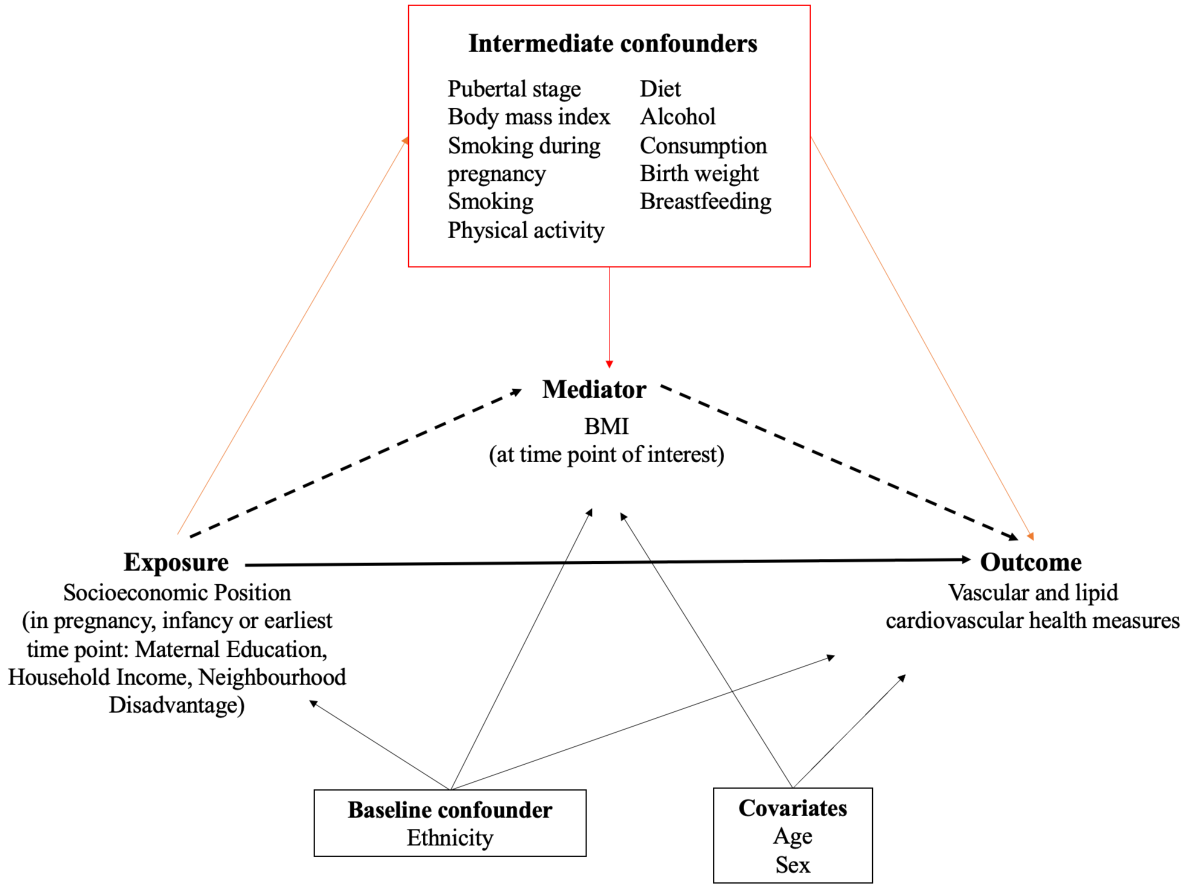


**Figure 1. Directed acyclic graph depicting the hypothesised causal relationships between socioeconomic position (SEP), body mass index (BMI), and cardiovascular health in childhood and adolescence.** Solid lines indicate direct causal pathways; the dotted line indicates the indirect effect of SEP on cardiovascular health through BMI. Covariates and baseline confounders are shown in the black boxes, while intermediate confounders (those caused by the exposure but confound the relationship between the mediator and outcome) are shown in the red box.

<u>Mathematical notation and causal estimand:</u>

Let $Y$ denote the outcome, $A$ denote a binary exposure, where $a = 1$ indicates exposed and $a = 0$ indicates unexposed. Let $M$ be the mediator, $Z$ the intermediate confounder and $C$ the confounders. Let $Y_a$ be the potential outcome when $A$ is set to $a$ and $Y_{a,m}$ the potential outcome when $A$ is set to $a$ and $M$ is set to $m$, where $a \in (0,1)$. Similarly, $M_a$ and $Z_a$ denote the potential value for $M$ and $Z$ respectively when setting the exposure $A$.

We considered a hypothetical mediator intervention that shifts the distribution of the mediator $M$. Our target estimand is the interventional indirect effect (IIE), $IIE = E(Y_1) - \theta$, where

$$\theta = \int E(Y_{1,m}|C = c)P(M_0 = m|C = c)P(Z_1 = z|C = c)P(C = c)dmdzdc$$

Under the identification assumptions described in Moreno-Betancur et al. (2021), $\theta$ can be identified as:

$$\theta = \int E(Y|A = 1, Z = Z, M = m, C = c)P(M|A = 0, C = c)P(Z|A = 1, C = c)P(C = c)dmdzdc$$

<u>Measurement of intermediate confounders</u>

*Maternal smoking during pregnancy*

Maternal smoking was dichotomised across all cohorts as 'any' versus 'none'. In the Barwon Infant Study (BIS) and Born in Bradford (BiB), pre-derived measures by cohort investigators were used. In the Longitudinal Study of Australian Children's Child Health CheckPoint (LSAC-CP), mothers were asked the number of cigarettes they smoked on average during pregnancy. Any value above zero was categorised as 'any' smoking. In NFBC 1986, mothers were asked in an antenatal survey whether they were previously a smoker, and whether their habits had changed during pregnancy. No previous smoking and having given up smoking were categorised as 'none' and sustained smoking was categorised as 'any'. In the Avon Longitudinal Study of Parents and Children, mothers were asked at 18 weeks gestation and 8 weeks post-partum whether they had smoked in the first three months or last two months (or two weeks) of pregnancy. Any reported smoking of cigarettes or cigars was categorised as 'any', while no reports of smoking were categorised as 'none'.

*Physical activity*

In BIS, physical activity was measured using an ActiGraph GT3X+ accelerometer (The ActiGraph, Pensacola, FL, USA), which measured activity for seven days outside of sleep and water. Mean hours of moderate- to vigorous-intensity physical activity (MVPA) per day was used in this study. In BiB, a seven-day recall questionnaire based on the Physical Activity Questionnaire for Older Children (PAQ-C) was delivered, but poor response rates (82% missing) prevented use in the mediation models. In LSAC-CP, physical activity was measured using a tri-axial GENEActiv accelerometer (Activinsights, Cambridge, United Kingdom) for eight days. Mean hours of MVPA was used in this study. In NFBC 1986, adolescents were asked how many hours per week they spent doing strenuous physical activity. Responses were coded as a continuous variable and included in mediation models. In ALSPAC, no data were available on physical activity at the time point of interest.

*Diet*

In BIS, a food frequency questionnaire was administered for mothers, who responded how frequently the study child consumed a range of fruits, vegetables, grains, sweets and other foods. Principal components analysis (PCA) was conducted using these data to derive two patterns of diet. A 'healthier' diet corresponded to a pattern to which fruit, vegetable, and whole grain consumption loaded more positively. A 'western' diet corresponded to a pattern to which sweets, meat, and snacks loaded more positively. The participants' scores for these patterns indicated how well they conformed to each type of diet, and both variables were included in mediation models. In BiB, a poor response rate to dietary questions prevented the derivation of a meaningful and informative diet variable. In LSAC-CP and NFBC 1986, an identical methodology to that in BIS was used with the dietary variables available in the cohort and time point. In ALSPAC, diet was not measured at the time point of interest.

*Birth weight*
Birth weight was available from all cohorts and included in mediation models as a continuous measure in grams.

*Breastfeeding duration*
Breastfeeding duration was dichotomised as ‘six months or more’ or ‘less than six months’. In BIS, mothers were asked how long they had breastfed in weeks, which was used to derive a dichotomous variable. In BiB, breastfeeding was only measured in a subset of participants, and a high degree of missingness (>90%) precluded inclusion in mediation models. In LSAC-CP, a pre-derived measure of breastfeeding (yes or no) at six months was used. In NFBC 1986, breastfeeding duration in months was used to derive a dichotomous variable. In ALSPAC, mothers who reported having breastfed for less than 3 months, 3-5 months or never were categorised as ‘less than six months’, and mothers who reported having breastfed for more than 6 months were categorised as ‘six months or more’.

*Pubertal stage*
Pubertal stage was included in BiB, LSAC-CP and NFBC 1986 models (not considered relevant in BIS and ALSPAC). In BiB, parents were asked if their children were showing signs of puberty (yes/no). In LSAC-CP, pubertal stage was assessed using a pubertal development scale and a sexual maturity scale to categorise participants as prepubertal, early pubertal, mid pubertal, late pubertal and post pubertal. Before inclusion in mediation models, the variable was dichotomised into ‘early pubertal’ (prepubertal, early pubertal, mid pubertal) or ‘late pubertal’ (late pubertal, post pubertal). In NFBC 1986, pubertal status was measured using the Tanner scale. If any of pubic hair, breast and genital development were at stages one, two or three, participants were categorised as ‘early pubertal’. The presence of any attribute at stages four or five resulted in categorisation as ‘late pubertal’.

*Current smoking*
Current smoking was included in NFBC 1986 and ASLPAC as participants were considered old enough to begin smoking. In NFBC 1986, participants were asked whether they currently smoke (yes/no). This variable was included in mediation models. In ALSPAC, a high degree of missingness (61%) in response to being asked whether they, the young person, had smoked in the last 30 days, precluded inclusion in the mediation models.

*Current alcohol consumption*
Alcohol consumption was included in NFBC 1986 and ALSPAC as participants were considered old enough to begin consuming alcohol. In NFBC 1986, alcohol consumption was derived by cohort investigators into a continuous measure of grams per day, based on questionnaire data. This variable was included in mediation models. In ALSPAC, participants were asked how frequently they had a drink containing alcohol. Those who responded “monthly or less” or “never” were categorised as ‘infrequent’ alcohol consumption and those who responded “two or four times a month”, “two or three times a week”, or “four or more times a week” were categorised as ‘frequent’ alcohol consumption.

**Supplemental Methods S5. Multiple imputation by chained equations**

Multiple imputation by chained equations was conducted to address missingness in all study cohorts.[97] Missing data on all exposure, mediator, outcome, and covariate data were imputed, and all analysis variables were included in the imputation models. Auxiliary variables, where available, were also included to improve prediction of missingness in the analysis variables.[98] In BIS, auxiliary variables were LDL-C, HDL-C, and triglycerides measured at birth and twelve months, body fat percentage, and mean triceps skinfold thickness. In BiB, auxiliary variables were abdominal circumference and body fat percentage. In LSAC-CP and NFBC 1986, the auxiliary variable was waist circumference. In ALSPAC, the auxiliary variable was Neighbourhood Quality Index (measured in pregnancy). Ordered categorical variables were imputed using proportional odds logistic regression, binary variables were imputed using logistic regression, and continuous variables were imputed using predictive mean matching. Sixty imputed datasets were generated across thirty iterations for each cohort. Estimates from multiple imputed datasets were combined using Rubin's rules.[99]

### Table S1. Subgroup results in girls, Northern Finland Birth Cohort 1986

| | Systolic Blood Pressure | | Diastolic Blood Pressure | | LDL-C | | HDL-C | | Total Triglycerides | | Non-HDL-C | | Remnant C | |
|---|---|---|---|---|---|---|---|---|---|---|---|---|---|---|
| | Regression Coefficient (95% CI) | P Value | Regression Coefficient (95% CI) | P Value | Regression Coefficient (95% CI) | P Value | Regression Coefficient (95% CI) | P Value | Regression Coefficient (95% CI) | P Value | Regression Coefficient (95% CI) | P Value | Regression Coefficient (95% CI) | P Value |
| | | | | | | | ***Unadjusted*** | | | | | | | |
| **Neighbourhood Disadvantage** | | | | | | | | | | | | | | |
| Low Disadvantage | Reference category | | | | | | | | | | | | | |
| Medium Disadvantage | 0.21 (-0.71, 1.12) | 0.66 | 0.27 (-0.34, 0.88) | 0.38 | 0.05 (-0.01, 0.1) | 0.13 | 0.01 (-0.03, 0.05) | 0.67 | 0.02 (-0.03, 0.06) | 0.53 | 0.07 (0, 0.15) | 0.07 | 0.03 (-0.01, 0.06) | 0.13 |
| High Disadvantage | -0.55 (-1.47, 0.36) | 0.23 | 0.36 (-0.26, 0.97) | 0.25 | 0.05 (-0.01, 0.1) | 0.11 | 0.00 (-0.04, 0.04) | 0.98 | 0.05 (0, 0.1) | 0.08 | 0.09 (0.01, 0.16) | 0.02 | 0.04 (0.01, 0.07) | 0.02 |
| **Parental Occupational Class** | | | | | | | | | | | | | | |
| High | Reference category | | | | | | | | | | | | | |
| Low | 2.40 (1.35, 3.46) | 0.00 | 0.97 (0.26, 1.68) | 0.01 | 0.03 (-0.04, 0.1) | 0.36 | -0.01 (-0.06, 0.04) | 0.68 | 0.01 (-0.05, 0.07) | 0.67 | 0.05 (-0.04, 0.14) | 0.24 | 0.02 (-0.02, 0.06) | 0.31 |
| **Maternal Education** | | | | | | | | | | | | | | |
| University Degree | Reference category | | | | | | | | | | | | | |
| Matriculation | -0.35 (-1.98, 1.27) | 0.67 | 0.23 (-0.86, 1.32) | 0.68 | 0.03 (-0.07, 0.13) | 0.54 | -0.01 (-0.08, 0.07) | 0.88 | 0.04 (-0.05, 0.13) | 0.42 | 0.06 (-0.07, 0.19) | 0.37 | 0.03 (-0.03, 0.09) | 0.34 |
| Any Vocational School/College | 0.36 (-1.2, 1.92) | 0.65 | 0.90 (-0.16, 1.95) | 0.10 | 0.04 (-0.06, 0.14) | 0.40 | -0.02 (-0.1, 0.05) | 0.50 | 0.06 (-0.03, 0.15) | 0.17 | 0.08 (-0.05, 0.21) | 0.23 | 0.04 (-0.02, 0.1) | 0.21 |
| Less than 10 Years of Schooling | 0.97 (-0.64, 2.59) | 0.24 | 1.11 (0, 2.22) | 0.05 | 0.04 (-0.07, 0.14) | 0.49 | -0.04 (-0.11, 0.03) | 0.28 | 0.11 (0.02, 0.2) | 0.02 | 0.08 (-0.05, 0.22) | 0.23 | 0.05 (-0.01, 0.11) | 0.11 |
| | | | | | | | ***Adjusted*** | | | | | | | |
| **Neighbourhood Disadvantage** | | | | | | | | | | | | | | |
| Low Disadvantage | Reference category | | | | | | | | | | | | | |
| Medium Disadvantage | 0.17 (-0.74, 1.09) | 0.71 | 0.25 (-0.36, 0.86) | 0.42 | 0.04 (-0.01, 0.1) | 0.14 | 0.01 (-0.03, 0.05) | 0.67 | 0.01 (-0.03, 0.06) | 0.55 | 0.07 (-0.01, 0.14) | 0.07 | 0.02 (-0.01, 0.06) | 0.15 |
| High Disadvantage | -0.48 (-1.39, 0.42) | 0.30 | 0.40 (-0.21, 1.01) | 0.20 | 0.05 (-0.01, 0.1) | 0.10 | 0.00 (-0.04, 0.04) | 0.98 | 0.05 (0, 0.1) | 0.07 | 0.09 (0.02, 0.16) | 0.02 | 0.04 (0.01, 0.08) | 0.01 |
| **Parental Occupational Class** | | | | | | | | | | | | | | |
| High | Reference category | | | | | | | | | | | | | |
| Low | 2.43 (1.38, 3.48) | 0.00 | 0.99 (0.28, 1.69) | 0.01 | 0.03 (-0.04, 0.1) | 0.35 | -0.01 (-0.06, 0.04) | 0.68 | 0.01 (-0.05, 0.07) | 0.66 | 0.05 (-0.03, 0.14) | 0.23 | 0.02 (-0.02, 0.06) | 0.28 |
| **Maternal Education** | | | | | | | | | | | | | | |
| University Degree | Reference category | | | | | | | | | | | | | |
| Matriculation | -0.41 (-2.03, 1.21) | 0.62 | 0.20 (-0.89, 1.29) | 0.72 | 0.03 (-0.07, 0.13) | 0.55 | -0.01 (-0.08, 0.07) | 0.88 | 0.04 (-0.06, 0.13) | 0.44 | 0.06 (-0.07, 0.19) | 0.40 | 0.03 (-0.03, 0.09) | 0.38 |
| Any Vocational School/College | 0.34 (-1.21, 1.89) | 0.67 | 0.88 (-0.17, 1.93) | 0.10 | 0.04 (-0.06, 0.14) | 0.41 | -0.02 (-0.1, 0.05) | 0.50 | 0.06 (-0.03, 0.15) | 0.18 | 0.08 (-0.05, 0.21) | 0.23 | 0.04 (-0.02, 0.1) | 0.22 |
| Less than 10 Years of Schooling | 0.92 (-0.69, 2.54) | 0.26 | 1.08 (-0.03, 2.18) | 0.06 | 0.04 (-0.07, 0.14) | 0.50 | -0.04 (-0.11, 0.03) | 0.28 | 0.11 (0.02, 0.2) | 0.02 | 0.08 (-0.06, 0.22) | 0.25 | 0.05 (-0.01, 0.1) | 0.13 |

### Table S2. Subgroup results in boys, Northern Finland Birth Cohort 1986

| | Systolic Blood Pressure | | Diastolic Blood Pressure | | LDL-C | | HDL-C | | Total Triglycerides | | Non-HDL-C | | Remnant C | |
|---|---|---|---|---|---|---|---|---|---|---|---|---|---|---|
| | Regression Coefficient (95% CI) | P Value | Regression Coefficient (95% CI) | P Value | Regression Coefficient (95% CI) | P Value | Regression Coefficient (95% CI) | P Value | Regression Coefficient (95% CI) | P Value | Regression Coefficient (95% CI) | P Value | Regression Coefficient (95% CI) | P Value |
| ***Unadjusted*** | | | | | | | | | | | | | | |
| **Neighbourhood Disadvantage** | | | | | | | | | | | | | | |
| Low Disadvantage | Reference category | | | | | | | | | | | | | |
| Medium Disadvantage | 0.05 (-0.96, 1.06) | 0.92 | 0.11 (-0.56, 0.77) | 0.75 | 0.03 (-0.03, 0.08) | 0.29 | 0.01 (-0.02, 0.05) | 0.47 | 0.00 (-0.05, 0.05) | 0.91 | 0.04 (-0.03, 0.11) | 0.24 | 0.01 (-0.02, 0.04) | 0.47 |
| High Disadvantage | -0.81 (-1.82, 0.2) | 0.12 | 0.11 (-0.54, 0.77) | 0.73 | 0.03 (-0.02, 0.08) | 0.24 | 0.02 (-0.01, 0.06) | 0.16 | -0.02 (-0.07, 0.03) | 0.40 | 0.04 (-0.03, 0.11) | 0.24 | 0.01 (-0.02, 0.04) | 0.54 |
| **Parental Occupational Class** | | | | | | | | | | | | | | |
| High | Reference category | | | | | | | | | | | | | |
| Low | 1.54 (0.36, 2.73) | 0.01 | 0.49 (-0.29, 1.27) | 0.22 | 0.05 (-0.02, 0.11) | 0.14 | 0.01 (-0.03, 0.05) | 0.63 | 0.01 (-0.05, 0.07) | 0.80 | 0.08 (0, 0.16) | 0.06 | 0.03 (-0.01, 0.06) | 0.12 |
| **Maternal Education** | | | | | | | | | | | | | | |
| University Degree | Reference category | | | | | | | | | | | | | |
| Matriculation | -1.47 (-3.3, 0.36) | 0.12 | -0.64 (-1.85, 0.57) | 0.30 | 0.02 (-0.09, 0.12) | 0.76 | 0.00 (-0.07, 0.07) | 0.94 | -0.01 (-0.1, 0.08) | 0.78 | 0.02 (-0.1, 0.15) | 0.72 | 0.01 (-0.05, 0.06) | 0.81 |
| Any Vocational School/College | -0.59 (-2.32, 1.15) | 0.51 | -0.31 (-1.45, 0.84) | 0.60 | 0.03 (-0.07, 0.13) | 0.56 | -0.01 (-0.08, 0.06) | 0.80 | -0.01 (-0.1, 0.08) | 0.88 | 0.04 (-0.08, 0.16) | 0.53 | 0.01 (-0.05, 0.06) | 0.74 |
| Less than 10 Years of Schooling | -0.19 (-2.04, 1.67) | 0.84 | 0.03 (-1.19, 1.25) | 0.96 | 0.03 (-0.08, 0.13) | 0.60 | -0.01 (-0.08, 0.06) | 0.73 | 0.04 (-0.06, 0.14) | 0.46 | 0.05 (-0.08, 0.17) | 0.47 | 0.02 (-0.04, 0.08) | 0.53 |
| ***Adjusted*** | | | | | | | | | | | | | | |
| **Neighbourhood Disadvantage** | | | | | | | | | | | | | | |
| Low Disadvantage | Reference category | | | | | | | | | | | | | |
| Medium Disadvantage | 0.04 (-0.96, 1.04) | 0.93 | 0.10 (-0.55, 0.76) | 0.76 | 0.03 (-0.03, 0.08) | 0.29 | 0.01 (-0.02, 0.05) | 0.47 | 0.00 (-0.05, 0.05) | 0.91 | 0.04 (-0.03, 0.11) | 0.25 | 0.01 (-0.02, 0.04) | 0.47 |
| High Disadvantage | -0.76 (-1.77, 0.24) | 0.14 | 0.15 (-0.51, 0.8) | 0.66 | 0.03 (-0.02, 0.08) | 0.24 | 0.02 (-0.01, 0.06) | 0.17 | -0.02 (-0.07, 0.03) | 0.40 | 0.04 (-0.03, 0.11) | 0.23 | 0.01 (-0.02, 0.04) | 0.50 |
| **Parental Occupational Class** | | | | | | | | | | | | | | |
| High | Reference category | | | | | | | | | | | | | |
| Low | **1.57 (0.4, 2.75)** | **0.01** | 0.51 (-0.26, 1.28) | 0.20 | 0.05 (-0.02, 0.11) | 0.14 | 0.01 (-0.03, 0.05) | 0.64 | 0.01 (-0.05, 0.07) | 0.80 | 0.08 (0, 0.16) | 0.06 | 0.03 (-0.01, 0.07) | 0.11 |
| **Maternal Education** | | | | | | | | | | | | | | |
| University Degree | Reference category | | | | | | | | | | | | | |
| Matriculation | -1.63 (-3.45, 0.18) | 0.08 | -0.75 (-1.95, 0.46) | 0.22 | 0.02 (-0.09, 0.12) | 0.76 | 0.00 (-0.07, 0.07) | 0.98 | -0.01 (-0.11, 0.08) | 0.76 | 0.02 (-0.1, 0.14) | 0.76 | 0.00 (-0.05, 0.06) | 0.90 |
| Any Vocational School/College | -0.64 (-2.36, 1.07) | 0.46 | -0.34 (-1.48, 0.79) | 0.55 | 0.03 (-0.07, 0.13) | 0.56 | -0.01 (-0.08, 0.06) | 0.81 | -0.01 (-0.1, 0.08) | 0.88 | 0.04 (-0.09, 0.16) | 0.55 | 0.01 (-0.05, 0.06) | 0.77 |
| Less than 10 Years of Schooling | -0.26 (-2.1, 1.59) | 0.79 | -0.01 (-1.23, 1.2) | 0.98 | 0.03 (-0.08, 0.13) | 0.60 | -0.01 (-0.08, 0.06) | 0.74 | 0.04 (-0.06, 0.13) | 0.46 | 0.05 (-0.08, 0.17) | 0.49 | 0.02 (-0.04, 0.08) | 0.56 |

### Table S3. Effect modification (interaction test) by sex, Northern Finland Birth Cohort 1986

| | Systolic Blood Pressure | | Diastolic Blood Pressure | | LDL-C | | HDL-C | | Total Triglycerides | | Non-HDL-C | | Remnant C | |
|---|---|---|---|---|---|---|---|---|---|---|---|---|---|---|
| | Regression Coefficient (95% CI) | P Value | Regression Coefficient (95% CI) | P Value | Regression Coefficient (95% CI) | P Value | Regression Coefficient (95% CI) | P Value | Regression Coefficient (95% CI) | P Value | Regression Coefficient (95% CI) | P Value | Regression Coefficient (95% CI) | P Value |
| ***Unadjusted*** | | | | | | | | | | | | | | |
| **Neighbourhood Disadvantage** | | | | | | | | | | | | | | |
| Low Disadvantage | Reference category | | | | | | | | | | | | | |
| Medium Disadvantage | -0.16 (-1.51, 1.2) | 0.82 | -0.17 (-1.07, 0.74) | 0.72 | -0.02 (-0.09, 0.06) | 0.67 | 0.00 (-0.04, 0.05) | 0.85 | -0.01 (-0.08, 0.06) | 0.72 | -0.03 (-0.13, 0.07) | 0.56 | -0.01 (-0.06, 0.03) | 0.57 |
| High Disadvantage | -0.26 (-1.61, 1.1) | 0.71 | -0.24 (-1.14, 0.65) | 0.59 | -0.02 (-0.09, 0.06) | 0.70 | 0.02 (-0.03, 0.07) | 0.36 | -0.07 (-0.14, 0) | 0.07 | -0.05 (-0.14, 0.05) | 0.36 | -0.03 (-0.08, 0.01) | 0.19 |
| **Parental Occupational Class** | | | | | | | | | | | | | | |
| High | Reference category | | | | | | | | | | | | | |
| Low | -0.86 (-2.45, 0.73) | 0.29 | -0.48 (-1.53, 0.56) | 0.36 | 0.02 (-0.08, 0.11) | 0.76 | 0.02 (-0.04, 0.09) | 0.54 | -0.01 (-0.09, 0.08) | 0.90 | 0.02 (-0.1, 0.14) | 0.69 | 0.01 (-0.04, 0.06) | 0.75 |
| **Maternal Education** | | | | | | | | | | | | | | |
| University Degree | Reference category | | | | | | | | | | | | | |
| Matriculation | -1.11 (-3.56, 1.34) | 0.37 | -0.87 (-2.52, 0.78) | 0.30 | -0.02 (-0.16, 0.13) | 0.84 | 0.00 (-0.1, 0.1) | 0.96 | -0.05 (-0.18, 0.08) | 0.43 | -0.04 (-0.22, 0.15) | 0.69 | -0.02 (-0.1, 0.06) | 0.60 |
| Any Vocational School/College | -0.95 (-3.29, 1.39) | 0.43 | -1.20 (-2.77, 0.36) | 0.13 | -0.01 (-0.15, 0.13) | 0.87 | 0.02 (-0.08, 0.11) | 0.76 | -0.07 (-0.19, 0.05) | 0.27 | -0.04 (-0.22, 0.14) | 0.66 | -0.03 (-0.11, 0.05) | 0.49 |
| Less than 10 Years of Schooling | -1.16 (-3.63, 1.31) | 0.36 | -1.08 (-2.71, 0.55) | 0.20 | -0.01 (-0.15, 0.14) | 0.91 | 0.03 (-0.07, 0.13) | 0.60 | -0.07 (-0.2, 0.06) | 0.27 | -0.04 (-0.22, 0.15) | 0.69 | -0.03 (-0.11, 0.05) | 0.49 |
| ***Adjusted*** | | | | | | | | | | | | | | |
| **Neighbourhood Disadvantage** | | | | | | | | | | | | | | |
| Low Disadvantage | Reference category | | | | | | | | | | | | | |
| Medium Disadvantage | -0.12 (-1.47, 1.23) | 0.86 | -0.14 (-1.04, 0.76) | 0.76 | -0.02 (-0.09, 0.06) | 0.68 | 0.00 (-0.04, 0.05) | 0.86 | -0.01 (-0.08, 0.06) | 0.73 | -0.03 (-0.13, 0.07) | 0.58 | -0.01 (-0.06, 0.04) | 0.60 |
| High Disadvantage | -0.31 (-1.66, 1.03) | 0.65 | -0.28 (-1.16, 0.61) | 0.54 | -0.02 (-0.09, 0.06) | 0.69 | 0.02 (-0.03, 0.08) | 0.35 | -0.07 (-0.14, 0) | 0.06 | -0.05 (-0.15, 0.05) | 0.34 | -0.03 (-0.08, 0.01) | 0.16 |
| **Parental Occupational Class** | | | | | | | | | | | | | | |
| High | Reference category | | | | | | | | | | | | | |
| Low | -0.87 (-2.45, 0.71) | 0.28 | -0.49 (-1.53, 0.55) | 0.36 | 0.02 (-0.08, 0.11) | 0.76 | 0.02 (-0.04, 0.09) | 0.54 | -0.01 (-0.09, 0.08) | 0.90 | 0.02 (-0.1, 0.14) | 0.70 | 0.01 (-0.05, 0.06) | 0.76 |
| **Maternal Education** | | | | | | | | | | | | | | |
| University Degree | Reference category | | | | | | | | | | | | | |
| Matriculation | -1.17 (-3.61, 1.26) | 0.35 | -0.91 (-2.55, 0.73) | 0.28 | -0.02 (-0.16, 0.13) | 0.84 | 0.00 (-0.1, 0.1) | 0.96 | -0.05 (-0.18, 0.07) | 0.43 | -0.04 (-0.22, 0.14) | 0.67 | -0.02 (-0.11, 0.06) | 0.57 |
| Any Vocational School/College | -0.96 (-3.29, 1.36) | 0.42 | -1.22 (-2.77, 0.34) | 0.12 | -0.01 (-0.15, 0.13) | 0.87 | 0.02 (-0.08, 0.11) | 0.75 | -0.07 (-0.19, 0.05) | 0.27 | -0.04 (-0.22, 0.14) | 0.65 | -0.03 (-0.11, 0.05) | 0.48 |
| Less than 10 Years of Schooling | -1.15 (-3.6, 1.3) | 0.36 | -1.07 (-2.69, 0.55) | 0.20 | -0.01 (-0.15, 0.14) | 0.91 | 0.03 (-0.07, 0.13) | 0.60 | -0.07 (-0.2, 0.06) | 0.28 | -0.04 (-0.22, 0.15) | 0.70 | -0.03 (-0.11, 0.05) | 0.50 |

#### Table S4. Mediation analysis, vascular measures, Longitudinal Study of Parents and Children's Child Health CheckPoint

| | Max. cIMT (mm) | | | | PWV (m/s) | | | | SBP (mmHg) | | | | DBP (mmHg) | | | |
|---|---|---|---|---|---|---|---|---|---|---|---|---|---|---|---|---|
| | **TCE (95% CI)** | **P Val** | **IIE (95% CI)** | **P Val** | **TCE (95% CI)** | **P Val** | **IIE (95% CI)** | **P Val** | **TCE (95% CI)** | **P Val** | **IIE (95% CI)** | **P Val** | **TCE (95% CI)** | **P Val** | **IIE (95% CI)** | **P Val** |
| **Neighbourhood Disadvantage** | | | | | | | | | | | | | | | | |
| High SEIFA | Reference group | | | | | | | | | | | | | | | |
| Medium SEIFA | -0.0002 (-0.006, 0.006) | 0.94 | -0.0001 (-0.002, 0.001) | 0.83 | 0.07 (0.01, 0.1) | 0.02 | 0.03 (0.006, 0.05) | 0.01 | 0.8 (-0.1, 1.6) | 0.10 | 0.6 (0.2, 1.0) | 0.003 | -0.1 (-0.8, 0.5) | 0.66 | 0.2 (-0.007, 0.3) | 0.06 |
| Low SEIFA | 0.007 (0.001, 0.01) | 0.02 | 0.001 (-0.0002, 0.002) | 0.11 | 0.1 (0.03, 0.2) | 0.003 | 0.02 (0.006, 0.04) | 0.008 | 1.5 (0.6, 2.5) | 0.001 | 0.8 (0.4, 1.2) | 0.00 | 0.5 (-0.1, 1.2) | 0.12 | 0.2 (0.04, 0.4) | 0.01 |
| **Household Income** | | | | | | | | | | | | | | | | |
| Above $75,000 | Reference group | | | | | | | | | | | | | | | |
| Below $75,000 | 0.003 (-0.002, 0.008) | 0.23 | 0.0004 (-0.0006, 0.001) | 0.42 | 0.03 (-0.03, 0.08) | 0.30 | 0.03 (0.01, 0.04) | 0.002 | 0.9 (0.2, 1.7) | 0.02 | 0.7 (0.3, 1.0) | 0.00 | 0.5 (-0.1, 1.0) | 0.11 | 0.2 (0.05, 0.3) | 0.007 |
| **Maternal Education** | | | | | | | | | | | | | | | | |
| Postgraduate Degree | Reference group | | | | | | | | | | | | | | | |
| Bachelor's Degree | 0.004 (-0.004, 0.01) | 0.34 | -0.0005 (-0.002, 0.0006) | 0.36 | 0.04 (-0.05, 0.1) | 0.36 | 0.0005 (-0.01, 0.02) | 0.94 | 1.3 (-0.05, 2.7) | 0.06 | -0.03 (-0.4, 0.4) | 0.90 | 1.4 (0.5, 2.4) | 0.004 | -0.005 (-0.1, 0.1) | 0.94 |
| Trade/Diploma/Cert/Other | 0.005 (-0.003, 0.01) | 0.25 | 0.002 (-0.00006, 0.003) | 0.06 | 0.05 (-0.04, 0.1) | 0.29 | 0.04 (0.01, 0.06) | 0.002 | 2.4 (1.0, 3.7) | 0.00 | 1.0 (0.4, 1.5) | 0.001 | 1.7 (0.7, 2.6) | 0.00 | 0.2 (0.03, 0.4) | 0.02 |
| Year 12 or Less | 0.005 (-0.004, 0.01) | 0.25 | -0.0006 (-0.002, 0.001) | 0.51 | 0.03 (-0.07, 0.1) | 0.62 | 0.04 (0.005, 0.07) | 0.02 | 1.9 (0.4, 3.3) | 0.01 | 0.8 (0.2, 1.4) | 0.02 | 1.4 (0.4, 2.4) | 0.008 | 0.3 (0.01, 0.6) | 0.04 |

### Table S5. Mediation analysis, metabolic measures, Longitudinal Study of Parents and Children’s Child Health CheckPoint

| | LDL-C (mmol/L) | | | | HDL-C (mmol/L) | | | | Total Triglycerides (mmol/L) | | | | Remnant-C (mmol/L) | | | | Non-HDL-C (mmol/L) | | | |
|---|---|---|---|---|---|---|---|---|---|---|---|---|---|---|---|---|---|---|---|---|
| | **TCE (95% CI)** | **P Val** | **IIE (95% CI)** | **P Val** | **TCE (95% CI)** | **P Val** | **IIE (95% CI)** | **P Val** | **TCE (95% CI)** | **P Val** | **IIE (95% CI)** | **P Val** | **TCE (95% CI)** | **P Val** | **IIE (95% CI)** | **P Val** | **TCE (95% CI)** | **P Val** | **IIE (95% CI)** | **P Val** |
| **Neighbourhood Disadvantage** | | | | | | | | | | | | | | | | | | | | |
| High SEIFA | Reference group | | | | | | | | | | | | | | | | | | | |
| Medium SEIFA | 0.0006 (-0.06, 0.06) | 0.98 | 0.003 (-0.01, 0.02) | 0.68 | -0.04 (-0.08, 0.003) | 0.07 | -0.006 (-0.02, 0.003) | 0.17 | 0.1 (0.02, 0.2) | 0.02 | 0.02 (-0.005, 0.05) | 0.12 | 0.03 (-0.01, 0.08) | 0.15 | 0.009 (-0.003, 0.020 | 0.13 | 0.03 (-0.04, 0.1) | 0.39 | 0.01 (-0.008, 0.03) | 0.25 |
| Low SEIFA | -0.01 (-0.07, 0.04) | 0.65 | 0.003 (-0.008, 0.01) | 0.58 | -0.06 (-0.09, -0.01) | 0.003 | -0.009 (-0.02, -0.0008) | 0.03 | 0.1 (0.05, 0.2) | 0.004 | 0.02 (-0.005, 0.04) | 0.12 | 0.02 (-0.02, 0.07) | 0.32 | 0.004 (-0.004, 0.01) | 0.32 | 0.01 (-0.07, 0.09) | 0.82 | 0.007 (-0.008, 0.02) | 0.34 |
| **Household Income** | | | | | | | | | | | | | | | | | | | | |
| Above $75,000 | Reference group | | | | | | | | | | | | | | | | | | | |
| Below $75,000 | -0.01 (-0.06, 0.04) | 0.62 | 0.003 (-0.006, 0.01) | 0.55 | -0.04 (-0.08, -0.01) | 0.01 | -0.009 (-0.02, -0.002) | 0.02 | 0.09 (0.009, 0.2) | 0.03 | 0.02 (0.002, 0.04) | 0.03 | -0.007 (-0.05, 0.03) | 0.71 | 0.007 (-0.0007, 0.010 | 0.07 | -0.02 (-0.09, 0.05) | 0.57 | 0.01 (-0.003, 0.02) | 0.14 |
| **Maternal Education** | | | | | | | | | | | | | | | | | | | | |
| Postgraduate Degree | Reference group | | | | | | | | | | | | | | | | | | | |
| Bachelor’s Degree | 0.02 (-0.06, 0.1) | 0.54 | -0.0006 (-0.009, 0.008) | 0.90 | 0.04 (-0.02, 0.09) | 0.18 | 0.002 (-0.007, 0.01) | 0.72 | -0.005 (-0.2, 0.2) | 0.95 | -0.001 (-0.02, 0.02) | 0.89 | 0.04 (-0.03, 0.1) | 0.22 | -0.0006 (-0.009, 0.007) | 0.89 | 0.07 (-0.05, 0.2) | 0.25 | -0.001 (-0.02, 0.01) | 0.88 |
| Trade/Diploma/Cert/Other | 0.02 (-0.07, 0.1) | 0.66 | 0.007 (-0.005, 0.02) | 0.30 | 0.002 (-0.05, 0.06) | 0.95 | -0.01 (-0.03, -0.003) | 0.01 | 0.09 (-0.07, 0.2) | 0.29 | 0.03 (0.001, 0.06) | 0.04 | 0.05 (-0.02, 0.1) | 0.14 | 0.01 (-0.001, 0.02) | 0.05 | 0.07 (-0.05, 0.2) | 0.24 | 0.02 (-0.0005, 0.04) | 0.06 |
| Year 12 or Less | 0.006 (-0.08, 0.1) | 0.89 | 0.0006 (-0.02, 0.02) | 0.95 | 0.02 (-0.04, 0.08) | 0.50 | -0.008 (-0.02, 0.004) | 0.19 | 0.09 (-0.07, 0.3) | 0.28 | 0.02 (-0.01, 0.05) | 0.26 | 0.03 (-0.04, 0.09) | 0.47 | 0.008 (-0.006, 0.02) | 0.25 | 0.03 (-0.09, 0.2) | 0.62 | 0.009 (-0.01, 0.03) | 0.46 |

#### Table S6. Mediation analysis, vascular measures, Avon Longitudinal Study of Parents and Children

| | Max. cIMT (mm) | | | | PWV (m/s) | | | | SBP (mmHg) | | | | DBP (mmHg) | | | |
|---|---|---|---|---|---|---|---|---|---|---|---|---|---|---|---|---|
| | TCE (95% CI) | P Val | IIE (95% CI) | P Val | TCE (95% CI) | P Val | IIE (95% CI) | P Val | TCE (95% CI) | P Val | IIE (95% CI) | P Val | TCE (95% CI) | P Val | IIE (95% CI) | P Val |
| **Neighbourhood Disadvantage** | | | | | | | | | | | | | | | | |
| Low Deprivation | Reference group | | | | | | | | | | | | | | | |
| Mediation Deprivation | -0.001 (-0.005, 0.003) | 0.60 | -0.0004 (-0.0009, 0.0001) | 0.13 | 0.02 (-0.04, 0.07) | 0.52 | 0.002 (-0.004, 0.009) | 0.45 | 0.8 (-0.02, 1.5) | 0.06 | 0.5 (0.2, 0.8) | 0.00 | 0.4 (-0.1, 0.9) | 0.13 | 0.3 (0.1, 0.5) | 0.00 |
| High Deprivation | -0.004 (-0.009, 0.00004) | 0.05 | -0.0003 (-0.001, 0005) | 0.46 | 0.05 (-0.01, 0.1) | 0.12 | 0.007 (-0.005, 0.02) | 0.27 | 0.9 (0.09, 1.8) | 0.03 | 1.0 (0.6, 1.3) | 0.00 | 0.8 (0.3, 1.4) | 0.005 | 0.6 (0.4, 0.8) | 0.00 |
| **Financial Difficulties** | | | | | | | | | | | | | | | | |
| No Difficulties | Reference group | | | | | | | | | | | | | | | |
| Difficulties | -0.003 (-0.006, 0.0007) | 0.13 | -0.0002 (-0.0006, 0.0003) | 0.47 | 0.03 (-0.01, 0.08) | 0.17 | 0.003 (-0.004, 0.009) | 0.40 | 0.7 (0.1, 1.3) | 0.02 | 0.4 (0.2, 0.7) | 0.00 | 0.2 (-0.2, 0.6) | 0.32 | 0.3 (0.1, 0.4) | 0.00 |
| **Maternal Education** | | | | | | | | | | | | | | | | |
| University Degree | Reference group | | | | | | | | | | | | | | | |
| A Levels | -0.005 (-0.01, -0.0002) | 0.04 | -0.000006 (-0.0005, 0.0004) | 0.79 | 0.002 (-0.06, 0.07) | 0.96 | 0.003 (-0.004, 0.01) | 0.45 | 0.8 (-0.09, 1.6) | 0.08 | 0.5 (0.2, 0.8) | 0.002 | 0.5 (-0.09, 1.1) | 0.10 | 0.3 (0.1, 0.5) | 0.002 |
| O Levels | -0.004 (-0.01, 0.0007) | 0.10 | -0.0006 (-0.001, 0.00007) | 0.08 | 0.06 (-0.003, 0.1) | 0.06 | 0.005 (-0.005, 0.01) | 0.32 | 0.8 (0.3, 2.0) | 0.006 | 0.8 (0.5, 1.1) | 0.00 | 1.0 (0.4, 1.6) | 0.001 | 0.5 (0.3, 0.7) | 0.00 |
| Vocational/CSE | -0.007 (-0.01, -0.002) | 0.005 | -0.0001 (-0.001, 0.001) | 0.81 | 0.08 (0.009, 0.2) | 0.03 | 0.003 (-0.02, 0.02) | 0.78 | 2.3 (1.4, 3.3) | 0.00 | 1.1 (0.8, 1.5) | 0.00 | 1.7 (1.1, 2.4) | 0.00 | 0.7 (0.5, 1.0) | 0.00 |

#### Table S7. Mediation analysis, vascular measures, Barwon Infant Study

| | Max. cIMT (mm) | | | | PWV (m/s) | | | | SBP (mmHg) | | | | DBP (mmHg) | | | |
|---|---|---|---|---|---|---|---|---|---|---|---|---|---|---|---|---|
| | **TCE (95% CI)** | **P Val** | **IIE (95% CI)** | **P Val** | **TCE (95% CI)** | **P Val** | **IIE (95% CI)** | **P Val** | **TCE (95% CI)** | **P Val** | **IIE (95% CI)** | **P Val** | **TCE (95% CI)** | **P Val** | **IIE (95% CI)** | **P Val** |
| **Neighbourhood Disadvantage** | | | | | | | | | | | | | | | | |
| High SEIFA | Reference group | | | | | | | | | | | | | | | |
| Medium SEIFA | 0.005 (-0.01, 0.02) | 0.53 | 0.001 (-0.003, 0.008) | 0.60 | -0.003 (-0.1, 0.1) | 0.97 | 0.03 (-0.02, 0.07) | 0.29 | -1.3 (-4.7, 2.1) | 0.47 | -0.5 (-2.3, 1.6) | 0.59 | -1.5 (-4.1, 0.7) | 0.22 | 0.07 (-1.1, 1.4) | 0.90 |
| Low SEIFA | 0.01 (0.002, 0.03) | 0.02 | 0.001 (-0.004 0.006) | 0.70 | -0.05 (-0.2, 0.08) | 0.42 | 0.006 (-0.04, 0.06) | 0.81 | -2.4 (-5.0, 0.6) | 0.10 | 0.03 (-1.5, 1.7) | 0.97 | -2.5 (-5.1, 0.08) | 0.05 | 0.2 (-1.2, 1.7) | 0.78 |
| **Household Income** | | | | | | | | | | | | | | | | |
| Above $75,000 | Reference group | | | | | | | | | | | | | | | |
| Below $75,000 | 0.007 (-0.006, 0.02) | 0.29 | -0.0002 (-0.006, 0.006) | 0.94 | -0.06 (-0.1, 0.08) | 0.40 | 0.03 (-0.05, 0.1) | 0.40 | 0.55 (-2.5, 3.6) | 0.73 | 0.60 (-1.2, 2.5) | 0.52 | 0.7 (-1.5, 2.9) | 0.50 | 0.6 (-0.8, 2.0) | 0.40 |
| **Maternal Education** | | | | | | | | | | | | | | | | |
| Postgraduate Degree | Reference group | | | | | | | | | | | | | | | |
| Bachelor’s Degree | -0.002 (-0.03, 0.02) | 0.87 | 0.002 (-0.004, 0.009) | 0.45 | 0.01 (-0.4, 0.4) | 0.97 | 0.0009 (-0.1, 0.2) | 0.99 | -1.4 (-5.0, 1.9) | 0.44 | 0.05 (-1.2, 1.5) | 0.95 | -1.2 (-4.0, 1.9) | 0.43 | 0.1 (-1.0, 1.5) | 0.83 |
| Trade/Diploma/Cert/Other | -0.01 (-0.03, 0.002) | 0.12 | -0.0004 (-0.008, 0.9) | 0.91 | 0.03 (-0.2, 0.2) | 0.77 | -0.002 (-0.1, 0.1) | 0.97 | 1.4 (-2.0, 4.9) | 0.41 | 0.8 (-0.8, 3.0) | 0.41 | 0.4 (-2.3, 2.5) | 0.77 | 0.8 (-0.5, 2.5) | 0.26 |
| Year 12 or Less | 0.004 (-0.009, 0.02) | 0.53 | 0.002 (-0.006, 0.6) | 0.64 | 0.05 (-0.2, 0.3) | 0.74 | 0.01 (-0.4, 0.4) | 0.96 | 0.04 (-4.6, 4.0) | 0.98 | 0.1 (-3.7, 4.1) | 0.94 | 0.07 (-3.4, 2.6) | 0.96 | 0.4 (-2.0, 2.7) | 0.74 |

#### Table S8. Mediation analysis, metabolic measures, Barwon Infant Study

| | LDL-C (mmol/L) | | | | HDL-C (mmol/L) | | | | Total Triglycerides (mmol/L) | | | | Remnant-C (mmol/L) | | | | Non-HDL-C (mmol/L) | | | |
|---|---|---|---|---|---|---|---|---|---|---|---|---|---|---|---|---|---|---|---|---|
| | **TCE (95% CI)** | **P Val** | **IIE (95% CI)** | **P Val** | **TCE (95% CI)** | **P Val** | **IIE (95% CI)** | **P Val** | **TCE (95% CI)** | **P Val** | **IIE (95% CI)** | **P Val** | **TCE (95% CI)** | **P Val** | **IIE (95% CI)** | **P Val** | **TCE (95% CI)** | **P Val** | **IIE (95% CI)** | **P Val** |
| **Neighbourhood Disadvantage** | | | | | | | | | | | | | | | | | | | | |
| High SEIFA | Reference group | | | | | | | | | | | | | | | | | | | |
| Medium SEIFA | -0.06 (-0.2, 0.06) | 0.33 | 0.009 (-0.03, 0.06) | 0.72 | -0.01 (-0.09, 0.07) | 0.76 | -0.008 (-0.03, 0.02) | 0.59 | 0.02 (-0.08, 0.1) | 0.71 | -0.02 (-0.06, 0.04) | 0.46 | -0.03 (-0.04, 0.4) | 0.40 | -0.001 (-0.03, 0.05) | 0.94 | -0.09 (-0.3, 0.06) | 0.29 | 0.008 (-0.06, 0.1) | 0.86 |
| Low SEIFA | -0.02 (-0.2, 0.1) | 0.76 | -0.01 (-0.09, 0.04) | 0.68 | -0.03 (-0.1, 0.06) | 0.46 | -0.002 (-0.04, 0.02) | 0.90 | 0.07 (-0.03, 0.2) | 0.15 | -0.002 (-0.04, 0.04) | 0.91 | 0.006 (-0.1, 0.1) | 0.91 | -0.01 (-0.05, 0.03) | 0.64 | -0.02 (-0.2, 0.2) | 0.88 | -0.02 (-0.1, 0.06) | 0.66 |
| **Household Income** | | | | | | | | | | | | | | | | | | | | |
| Above $75,000 | Reference group | | | | | | | | | | | | | | | | | | | |
| Below $75,000 | 0.02 (-0.1, 0.2) | 0.77 | 0.03 (-0.02, 0.08) | 0.30 | -0.007 (-0.09, 0.07) | 0.86 | 0.003 (-0.04, 0.04) | 0.87 | 0.04 (-0.06, 0.1) | 0.39 | -0.0001 (-0.04, 0.06) | 0.96 | 0.01 (-0.07, 0.09) | 0.77 | 0.01 (-0.02, 0.06) | 0.52 | 0.03 (-0.2, 0.2) | 0.75 | 0.04 (-0.04, 0.1) | 0.40 |
| **Maternal Education** | | | | | | | | | | | | | | | | | | | | |
| Postgraduate Degree | Reference group | | | | | | | | | | | | | | | | | | | |
| Bachelor's Degree | 0.009 (-0.1, 0.1) | 0.90 | 0.02 (-0.04, 0.09) | 0.57 | -0.04 (-0.2, 0.08) | 0.53 | 0.007 (-0.08, 0.1) | 0.88 | 0.1 (0.004, 0.2) | 0.04 | -0.01 (-0.07, 0.04) | 0.62 | 0.05 (-0.06, 0.1) | 0.35 | 0.003 (-0.04, 0.05) | 0.90 | 0.06 (-0.2, 0.3) | 0.65 | 0.02 (-0.1, 0.2) | 0.81 |
| Trade/Diploma/Cert/Other | -0.08 (-0.2, 0.06) | 0.32 | 0.02 (-0.05, 0.1) | 0.54 | -0.02 (-0.1, 0.07) | 0.70 | -0.004 (-0.1, 0.1) | 0.95 | 0.04 (-0.06, 0.1) | 0.40 | -0.02 (-0.09, 0.05) | 0.56 | -0.03 (-0.1, 0.07) | 0.58 | 0.003 (-0.06, 0.08) | 0.92 | -0.1 (-0.3, 0.1) | 0.38 | 0.03 (-0.1, 0.2) | 0.72 |
| Year 12 or Less | -0.007 (-0.2, 0.2) | 0.93 | 0.009 (-0.08, 0.1) | 0.85 | -0.03 (-0.1, 0.07) | 0.62 | 0.02 (-0.08, 0.09) | 0.70 | 0.1 (-0.04, 0.3) | 0.17 | -0.01 (-0.2, 0.1) | 0.85 | 0.008 (-0.1, 0.1) | 0.90 | 0.0007 (-0.09, 0.1) | 0.99 | -0.0006 (-0.3, 0.3) | 0.99 | 0.01 (-0.2, 0.3) | 0.65 |

### Table S9. Mediation analysis (Systolic blood pressure, diastolic blood pressure, low-density lipoprotein cholesterol), Born in Bradford

| | SBP (mmHg) | | | | DBP (mmHg) | | | | LDL-C (mmol/L) | | | |
|---|---|---|---|---|---|---|---|---|---|---|---|---|
| | **TCE (95% CI)** | **P Val** | **IIE (95% CI)** | **P Val** | **TCE (95% CI)** | **P Val** | **IIE (95% CI)** | **P Val** | **TCE (95% CI)** | **P Val** | **IIE (95% CI)** | **P Val** |
| **Neighbourhood Deprivation** | | | | | | | | | | | | |
| Low deprivation | Reference group | | | | | | | | | | | |
| Medium deprivation | -0.5 (-1.5, 0.6) | 0.36 | 0.04 (-0.2, 0.3) | 0.80 | -0.3 (-1.2, 0.7) | 0.58 | 0.05 (-0.1, 0.2) | 0.60 | -0.002 (-0.05, 0.04) | 0.93 | -0.0004 (-0.006, 0.005) | 0.89 |
| High deprivation | 0.1 (-0.9, 1.2) | 0.83 | 0.3 (-0.05, 0.6) | 0.02 | 0.6 (-0.4, 1.6) | 0.22 | 0.2 (0.03, 0.40 | 0.03 | 0.01 (-0.03, 0.06) | 0.63 | 0.003 (-0.004, 0.01) | 0.43 |
| **Financial Security** | | | | | | | | | | | | |
| Financially secure | Reference group | | | | | | | | | | | |
| Financially struggling | -0.4 (-1.2, 0.5) | 0.39 | 0.3 (0.02, 0.5) | 0.03 | -0.8 (-1.6, -0.04) | 0.04 | 0.2 (0.03, 0.4) | 0.03 | 0.006 (-0.03, 0.05) | 0.78 | 0.002 (-0.004, 0.009) | 0.51 |
| **Maternal Education** | | | | | | | | | | | | |
| Tertiary Degree (UK or Foreign) | Reference group | | | | | | | | | | | |
| A Level/A Level Equivalent/Other (e.g. City and Guilds, RSA/OCR, BTEC, Foreign Other) | -0.7 (-2.0, 0.6) | 0.28 | 0.2 (-0.1, 0.5) | 0.20 | 0.4 (-0.8, 1.6) | 0.52 | 0.2 (-0.06, 0.4) | 0.16 | 0.02 (-0.04, 0.07) | 0.56 | 0.002 (-0.005, 0.008) | 0.63 |
| 5 GCSEs or Equivalent | 0.2 (-0.9, 1.2) | 0.78 | 0.2 (-0.05, 0.5) | 0.12 | 0.06 (-0.9, 1.0) | 0.90 | 0.2 (-0.03, 0.4) | 0.10 | 0.01 (-0.04, 0.06) | 0.61 | 0.001 (-0.006, 0.008) | 0.71 |
| Less than 5 GCSEs | 0.04 (-1.1, 1.2) | 0.95 | 0.4 (0.1, 0.8) | 0.007 | 0.02 (-1.0, 1.1 | 0.97 | 0.3 (0.06, 0.5) | 0.02 | 0.003 (-0.05, 0.06) | 0.93 | 0.005 (-0.006, 0.01) | 0.31 |

#### Table S10. Mediation analysis (high-density lipoprotein cholesterol, total triglycerides, remnant cholesterol, non-high-density lipoprotein cholesterol), Born in Bradford

| | **HDL-C (mmol/L)** | | | | **Total Triglycerides (mmol/L)** | | | | **Remnant-C (mmol/L)** | | | | **Non-HDL-C (mmol/L)** | | | |
|---|---|---|---|---|---|---|---|---|---|---|---|---|---|---|---|---|
| | **TCE (95% CI)** | **P Val** | **IIE (95% CI)** | **P Val** | **TCE (95% CI)** | **P Val** | **IIE (95% CI)** | **P Val** | **TCE (95% CI)** | **P Val** | **IIE (95% CI)** | **P Val** | **TCE (95% CI)** | **P Val** | **IIE (95% CI)** | **P Val** |
| **Neighbourhood Deprivation** | | | | | | | | | | | | | | | | |
| Low deprivation | Reference group | | | | | | | | | | | | | | | |
| Medium deprivation | -0.02 (-0.06, 0.01) | 0.19 | -0.000008 (-0.007, 0.007) | 0.99 | 0.002 (-0.08, 0.08) | 0.97 | 0.001 9-0.01, 0.02) | 0.84 | -0.002 (-0.03, 0.03) | 0.91 | -0.0003 (-0.004, 0.003) | 0.86 | -0.004 (-0.05, 0.04) | 0.88 | -0.0008 (-0.008, 0.008) | 0.84 |
| High deprivation | -0.03 (-0.06, 0.002) | 0.06 | -0.009 (-0.02, -0.001) | 0.03 | 0.07 (-0.009, 0.1) | 0.08 | 0.02 (-0.009, 0.04) | 0.06 | 0.008 (-0.02, 0.04) | 0.56 | 0.002 (-0.003, 0.006) | 0.42 | 0.02 (-0.03, 0.07) | 0.43 | 0.005 (-0.003, 0.01) | 0.26 |
| **Financial Security** | | | | | | | | | | | | | | | | |
| Financially secure | Reference group | | | | | | | | | | | | | | | |
| Financially struggling | 0.0007 (-0.02, 0.03) | 0.95 | -0.007 (-0.01, -0.0003) | 0.04 | 0.009 (-0.06, 0.07) | 0.80 | 0.01 (-0.001, 0.03) | 0.07 | 0.004 (-0.02, 0.03) | 0.69 | 0.002 (-0.002, 0.006) | 0.38 | 0.01 (-0.03, 0.05) | 0.64 | 0.004 (-0.004, 0.01) | 0.30 |
| **Maternal Education** | | | | | | | | | | | | | | | | |
| Tertiary Degree (UK or Foreign) | Reference group | | | | | | | | | | | | | | | |
| A Level/A Level Equivalent/Other (e.g. City and Guilds, RSA/OCR, BTEC, Foreign Other) | 0.004 (-0.03, 0.04) | 0.84 | -0.005 (-0.01, 0.003) | 0.26 | 0.001 (-0.09, 0.09) | 0.98 | 0.009 (-0.007, 0.03) | 0.27 | 0.004 (-0.03, 0.03) | 0.82 | 0.001 (-0.003, 0.006) | 0.53 | 0.02 (-0.04, 0.08) | 0.51 | 0.003 (-0.005, 0.01) | 0.49 |
| 5 GCSEs or Equivalent | -0.006 (-0.04, 0.03) | 0.71 | -0.006 (-0.01, 0.002) | 0.15 | 0.02 (-0.06, 0.1) | 0.62 | 0.01 (-0.005, 0.03) | 0.18 | 0.003 (-0.02, 0.03) | 0.81 | 0.0009 (-0.001, 0.01) | 0.14 | 0.02 (-0.04, 0.07) | 0.54 | 0.002 (-0.005, 0.01) | 0.56 |
| Less than 5 GCSEs | -0.03 (-0.07, 0.007) | 0.11 | -0.01 (-0.02, -0.003) | 0.007 | 0.06 (-0.04, 0.2) | 0.25 | 0.02 (0.002, 0.05) | 0.03 | -0.008 (-0.04, 0.2) | 0.63 | 0.004 (-0.001, 0.01) | 0.14 | -0.005 (-0.06, 0.05) | 0.86 | 0.01 (-0.002, 0.02) | 0.10 |

**Table S11. Mediation analysis (Systolic blood pressure, diastolic blood pressure, low-density lipoprotein cholesterol), Northern Finland Birth Cohort 1986**

| | SBP (mmHg) | | | | DBP (mmHg) | | | | LDL-C (mmol/L) | | | |
|---|---|---|---|---|---|---|---|---|---|---|---|---|
| | **TCE (95% CI)** | **TCE (95% CI)** | **P Val** | **IIE (95% CI)** | **TCE (95% CI)** | **P Val** | **IIE (95% CI)** | **TCE (95% CI)** | **TCE (95% CI)** | **P Val** | **IIE (95% CI)** | **TCE (95% CI)** |
| **Neighbourhood Disadvantage** | | | | | | | | | | | | |
| Low Disadvantage | Reference group | | | | | | | | | | | |
| Medium Disadvantage | 0.1 (-0.5, 0.8) | 0.70 | -0.06 (-0.3, 0.2) | 0.61 | 0.2 (-0.3, 0.6) | 0.41 | -0.03 (-0.2, 0.1) | 0.68 | 0.04 (-0.003, 0.08) | 0.07 | 0.0005 (-0.006, 0.007) | 0.88 |
| High Disadvantage | -0.6 (-1.3, 0.06) | 0.07 | 0.1 (-0.1, 0.4) | 0.27 | 0.3 (-0.2, 0.7) | 0.24 | 0.09 (-0.05, 0.2) | 0.22 | 0.04 (0.0009, 0.08) | 0.05 | 0.003 (-0.002, 0.009) | 0.25 |
| **Parental Occupational Class** | | | | | | | | | | | | |
| High | Reference group | | | | | | | | | | | |
| Low | 2.0 (1.2, 2.8) | 0.00 | 0.3 (-0.03, 0.5) | 0.08 | 0.7 (0.2, 1.3) | 0.006 | 0.2 (-0.01, 0.4) | 0.07 | 0.04 (-0.006, 0.09) | 0.09 | 0.005 (-0.002, 0.01) | 0.17 |
| **Maternal Education** | | | | | | | | | | | | |
| University Degree | Reference group | | | | | | | | | | | |
| Matriculation | -0.9 (-2.1, 0.4) | 0.18 | -0.03 (-0.4, 0.3) | 0.88 | -0.4 (-1.3, 0.5) | 0.37 | -0.02 (-0.2, 0.2) | 0.82 | 0.02 (-0.05, 0.1) | 0.57 | 0.001 (-0.005, 0.007) | 0.76 |
| Any Vocational School/College | 0.01 (-1.2, 1.2) | 0.99 | 0.3 (-0.03, 0.7) | 0.07 | 0.1 (-0.7, 1.0) | 0.74 | 0.2 (-0.01, 0.4) | 0.07 | 0.03 (-0.04, 0.1) | 0.37 | 0.006 (-0.001, 0.01) | 0.10 |
| Less than 10 years of schooling | 0.5 (-0.8, 1.8) | 0.46 | 0.4 (0.07, 0.8) | 0.02 | 0.4 (-0.5, 1.3) | 0.40 | 0.3 (0.05, 0.5) | 0.02 | 0.03 (-0.05, 0.1) | 0.45 | 0.007 (-0.001, 0.01) | 0.11 |

**Table S12. Mediation analysis (high-density lipoprotein cholesterol, total triglycerides, remnant cholesterol, non-high-density lipoprotein cholesterol), Northern Finland Birth Cohort 1986**

| | HDL-C (mmol/L) | | | | Total Triglycerides (mmol/L) | | | | Remnant-C (mmol/L) | | | | Non-HDL-C (mmol/L) | | | |
|---|---|---|---|---|---|---|---|---|---|---|---|---|---|---|---|---|
| | TCE (95% CI) | P Val | IIE (95% CI) | P Val | TCE (95% CI) | P Val | IIE (95% CI) | P Val | TCE (95% CI) | P Val | IIE (95% CI) | P Val | TCE (95% CI) | P Val | IIE (95% CI) | P Val |
| **Neighbourhood Disadvantage** | | | | | | | | | | | | | | | | |
| Low Disadvantage | Reference group | | | | | | | | | | | | | | | |
| Medium Disadvantage | 0.01 (-0.01, 0.04) | 0.36 | 0.001 (-0.004, 0.006) | 0.64 | 0.009 (-0.03, 0.05) | 0.64 | -0.001 (-0.01, 0.008) | 0.79 | 0.02 (-0.005, 0.04) | 0.12 | -0.001 (-0.007, 0.005) | 0.74 | 0.06 (0.004, 0.1) | 0.04 | -0.005 (-0.01, 0.01) | 0.93 |
| High Disadvantage | 0.01 (-0.01, 0.04) | 0.33 | -0.002 (-0.007, 0.003) | 0.38 | 0.01 (-0.02, 0.05) | 0.46 | 0.005 (-0.004, 0.01) | 0.25 | 0.03 (0.005, 0.05) | 0.02 | 0.003 (-0.002, 0.009) | 0.23 | 0.07 (0.02, 0.1) | 0.009 | 0.007 (-0.003, 0.02) | 0.18 |
| **Parental Occupational Class** | | | | | | | | | | | | | | | | |
| High | Reference group | | | | | | | | | | | | | | | |
| Low | 0.001 (-0.03, 0.03) | 0.94 | -0.004 (-0.009, 0.002) | 0.21 | 0.01 (-0.03, 0.05) | 0.65 | 0.008 (-0.003, 0.02) | 0.16 | 0.02 (-0.002, 0.05) | 0.07 | 0.005 (-0.001, 0.01) | 0.12 | 0.06 (0.005, 0.1) | 0.04 | 0.01 (-0.002, 0.02) | 0.10 |
| **Maternal Education** | | | | | | | | | | | | | | | | |
| University Degree | Reference group | | | | | | | | | | | | | | | |
| Matriculation | 0.003 (-0.05, 0.06) | 0.93 | 0.0009 (-0.006, 0.008) | 0.81 | 0.004 (-0.07, 0.08) | 0.93 | -0.0006 (-0.01, 0.01) | 0.92 | 0.009 (-0.03, 0.06) | 0.52 | 0.00002 (-0.007, 0.007) | 0.99 | 0.04 (-0.06, 0.1) | 0.47 | 0.001 (-0.01, 0.01) | 0.88 |
| Any Vocational School/College | -0.01 (-0.07, 0.04) | 0.68 | -0.006 (-0.01, 0.001) | 0.11 | 0.02 (-0.05, 0.1) | 0.57 | 0.01 (-0.002, 0.02) | 0.09 | 0.02 (-0.02, 0.07) | 0.31 | 0.007 (-0.0007, 0.01) | 0.07 | 0.06 (-0.04, 0.2) | 0.25 | 0.01 (-0.0006, 0.03) | 0.06 |
| Less than 10 years of schooling | -0.02 (-0.08, 0.04) | 0.47 | -0.006 (-0.01, 0.0004) | 0.07 | 0.06 (-0.02, 0.1) | 0.12 | 0.02 (0.001, 0.03) | 0.04 | 0.03 (-0.01, 0.08) | 0.17 | 0.009 (0.0009, 0.02) | 0.03 | 0.06 (-0.04, 0.2) | 0.23 | 0.02 (0.001, 0.03) | 0.03 |

### Table S13. Subgroup results in girls, Barwon Infant Study

| | Max. cIMT (mm) | | PWV (m/s) | | Systolic Blood Pressure (mmHg) | | Diastolic Blood Pressure (mmHg) | | LDL-C (mmol/L) | | HDL-C (mmol/L) | | Total Triglycerides (mmol/L) | | Remnant Cholesterol (mmol/L) | | Non-HDL-C (mmol/L) | |
|---|---|---|---|---|---|---|---|---|---|---|---|---|---|---|---|---|---|---|
| | Regression Coefficient (95% CI) | P Value | Regression Coefficient (95% CI) | P Value | Regression Coefficient (95% CI) | P Value | Regression Coefficient (95% CI) | P Value | Regression Coefficient (95% CI) | P Value | Regression Coefficient (95% CI) | P Value | Regression Coefficient (95% CI) | P Value | Regression Coefficient (95% CI) | P Value | Regression Coefficient (95% CI) | P Value |
| ***Unadjusted*** | | | | | | | | | | | | | | | | | | |
| **Neighbourhood Disadvantage** | | | | | | | | | | | | | | | | | | |
| High SEIFA | Reference category | | | | | | | | | | | | | | | | | |
| Medium SEIFA | 0.00 (-0.01, 0.01) | 0.52 | 0.00 (-0.12, 0.12) | 0.99 | -0.57 (-3.01, 1.87) | 0.65 | -1.09 (-2.87, 0.69) | 0.23 | -0.07 (-0.17, 0.02) | 0.13 | 0.00 (-0.07, 0.06) | 0.91 | -0.02 (-0.13, 0.09) | 0.67 | -0.06 (-0.12, 0) | 0.05 | -0.13 (-0.26, 0) | 0.05 |
| Low SEIFA | -0.01 (-0.02, 0.01) | 0.31 | -0.01 (-0.14, 0.12) | 0.92 | -0.86 (-3.48, 1.76) | 0.52 | -0.51 (-2.41, 1.39) | 0.60 | -0.05 (-0.15, 0.06) | 0.39 | 0.01 (-0.07, 0.08) | 0.87 | -0.03 (-0.14, 0.09) | 0.63 | -0.04 (-0.11, 0.03) | 0.23 | -0.09 (-0.23, 0.06) | 0.24 |
| **Household Income** | | | | | | | | | | | | | | | | | | |
| Above $75,000; population median | Reference category | | | | | | | | | | | | | | | | | |
| Below $75,000; population median | -0.01 (-0.02, 0) | 0.28 | 0.02 (-0.1, 0.14) | 0.74 | -1.20 (-3.53, 1.13) | 0.31 | -0.25 (-1.96, 1.46) | 0.78 | -0.06 (-0.15, 0.04) | 0.23 | -0.04 (-0.1, 0.03) | 0.28 | 0.04 (-0.07, 0.14) | 0.49 | -0.04 (-0.1, 0.02) | 0.19 | -0.09 (-0.22, 0.03) | 0.15 |
| **Maternal Education** | | | | | | | | | | | | | | | | | | |
| Postgraduate Degree | Reference category | | | | | | | | | | | | | | | | | |
| Bachelor's Degree | 0.01 (-0.01, 0.02) | 0.26 | -0.03 (-0.18, 0.11) | 0.65 | 0.24 (-2.63, 3.12) | 0.87 | 0.96 (-1.15, 3.07) | 0.37 | -0.08 (-0.2, 0.04) | 0.19 | 0.03 (-0.06, 0.11) | 0.54 | -0.07 (-0.2, 0.06) | 0.28 | -0.07 (-0.14, 0) | 0.06 | -0.15 (-0.31, 0.01) | 0.07 |
| Trade/Diploma/Certificate/Other | 0.01 (-0.01, 0.02) | 0.50 | -0.04 (-0.2, 0.12) | 0.60 | -0.05 (-3.16, 3.06) | 0.98 | 0.71 (-1.6, 3.02) | 0.54 | -0.07 (-0.2, 0.06) | 0.28 | 0.01 (-0.08, 0.1) | 0.78 | -0.07 (-0.21, 0.08) | 0.36 | -0.06 (-0.14, 0.01) | 0.10 | -0.14 (-0.31, 0.04) | 0.12 |
| Less than Year 12 of high school | 0.00 (-0.01, 0.02) | 0.75 | -0.04 (-0.22, 0.15) | 0.70 | -0.06 (-3.65, 3.54) | 0.97 | 0.45 (-2.18, 3.07) | 0.74 | -0.11 (-0.27, 0.04) | 0.14 | 0.04 (-0.06, 0.14) | 0.42 | -0.04 (-0.22, 0.13) | 0.61 | -0.10 (-0.18, -0.01) | 0.04 | -0.21 (-0.41, -0.01) | 0.04 |
| ***Adjusted*** | | | | | | | | | | | | | | | | | | |
| **Neighbourhood Disadvantage** | | | | | | | | | | | | | | | | | | |
| High SEIFA | Reference category | | | | | | | | | | | | | | | | | |
| Medium SEIFA | 0.00 (-0.01, 0.01) | 0.49 | 0.00 (-0.12, 0.12) | 0.95 | -0.47 (-2.92, 1.98) | 0.71 | -1.08 (-2.88, 0.71) | 0.24 | -0.07 (-0.17, 0.02) | 0.12 | 0.00 (-0.07, 0.06) | 0.90 | -0.02 (-0.13, 0.09) | 0.66 | -0.06 (-0.12, 0) | 0.05 | -0.13 (-0.27, 0) | 0.04 |
| Low SEIFA | -0.01 (-0.02, 0.01) | 0.32 | -0.01 (-0.14, 0.12) | 0.88 | -0.87 (-3.49, 1.75) | 0.52 | -0.54 (-2.44, 1.36) | 0.57 | -0.05 (-0.15, 0.06) | 0.39 | 0.00 (-0.07, 0.08) | 0.90 | -0.03 (-0.14, 0.09) | 0.64 | -0.04 (-0.11, 0.03) | 0.23 | -0.09 (-0.23, 0.06) | 0.24 |
| **Household Income** | | | | | | | | | | | | | | | | | | |
| Above $75,000; population median | Reference category | | | | | | | | | | | | | | | | | |
| Below $75,000; population median | -0.01 (-0.02, 0) | 0.31 | 0.03 (-0.09, 0.15) | 0.64 | -1.03 (-3.36, 1.3) | 0.39 | -0.20 (-1.91, 1.52) | 0.82 | -0.06 (-0.15, 0.04) | 0.23 | -0.04 (-0.1, 0.03) | 0.28 | 0.03 (-0.07, 0.14) | 0.52 | -0.04 (-0.1, 0.02) | 0.17 | -0.10 (-0.23, 0.03) | 0.14 |
| **Maternal Education** | | | | | | | | | | | | | | | | | | |
| Postgraduate Degree | Reference category | | | | | | | | | | | | | | | | | |
| Bachelor's Degree | 0.01 (-0.01, 0.02) | 0.26 | -0.04 (-0.18, 0.1) | 0.60 | 0.21 (-2.67, 3.1) | 0.88 | 0.91 (-1.21, 3.03) | 0.40 | -0.08 (-0.2, 0.04) | 0.18 | 0.02 (-0.06, 0.11) | 0.57 | -0.07 (-0.2, 0.06) | 0.30 | -0.07 (-0.14, 0) | 0.06 | -0.15 (-0.31, 0.01) | 0.07 |
| Trade/Diploma/Certificate/Other | 0.01 (-0.01, 0.02) | 0.48 | -0.03 (-0.19, 0.13) | 0.74 | 0.15 (-2.97, 3.27) | 0.92 | 0.84 (-1.49, 3.16) | 0.48 | -0.07 (-0.2, 0.06) | 0.29 | 0.02 (-0.08, 0.11) | 0.74 | -0.07 (-0.22, 0.07) | 0.33 | -0.07 (-0.14, 0.01) | 0.09 | -0.14 (-0.31, 0.04) | 0.13 |
| Less than Year 12 of high school | 0.00 (-0.01, 0.02) | 0.76 | -0.03 (-0.21, 0.15) | 0.73 | -0.02 (-3.62, 3.58) | 0.99 | 0.49 (-2.14, 3.12) | 0.72 | -0.11 (-0.27, 0.04) | 0.15 | 0.04 (-0.06, 0.14) | 0.41 | -0.05 (-0.22, 0.13) | 0.60 | -0.10 (-0.18, -0.01) | 0.04 | -0.21 (-0.41, -0.01) | 0.04 |

### Table S14. Subgroup results in boys, Barwon Infant Study

| | Max. cIMT (mm) | | PWV (m/s) | | Systolic Blood Pressure (mmHg) | | Diastolic Blood Pressure (mmHg) | | LDL-C (mmol/L) | | HDL-C (mmol/L) | | Total Triglycerides (mmol/L) | | Remnant Cholesterol (mmol/L) | | Non-HDL-C (mmol/L) | |
|---|---|---|---|---|---|---|---|---|---|---|---|---|---|---|---|---|---|---|
| | Regression Coefficient (95% CI) | P Value | Regression Coefficient (95% CI) | P Value | Regression Coefficient (95% CI) | P Value | Regression Coefficient (95% CI) | P Value | Regression Coefficient (95% CI) | P Value | Regression Coefficient (95% CI) | P Value | Regression Coefficient (95% CI) | P Value | Regression Coefficient (95% CI) | P Value | Regression Coefficient (95% CI) | P Value |
| ***Unadjusted*** | | | | | | | | | | | | | | | | | | |
| **Neighbourhood Disadvantage** | | | | | | | | | | | | | | | | | | |
| High SEIFA | Reference category | | | | | | | | | | | | | | | | | |
| Medium SEIFA | 0.01 (-0.01, 0.02) | 0.29 | 0.08 (-0.05, 0.2) | 0.22 | -3.40 (-5.95, -0.86) | 0.01 | -2.35 (-4.39, -0.3) | 0.02 | -0.02 (-0.12, 0.08) | 0.73 | 0.00 (-0.07, 0.07) | 0.97 | 0.01 (-0.09, 0.12) | 0.79 | 0.00 (-0.06, 0.06) | 0.97 | -0.02 (-0.16, 0.12) | 0.79 |
| Low SEIFA | 0.00 (-0.01, 0.01) | 0.95 | 0.16 (0.04, 0.29) | 0.01 | -1.66 (-4.18, 0.86) | 0.20 | -1.22 (-3.21, 0.77) | 0.23 | -0.02 (-0.13, 0.08) | 0.66 | -0.03 (-0.1, 0.04) | 0.44 | 0.02 (-0.09, 0.14) | 0.67 | -0.01 (-0.07, 0.06) | 0.84 | -0.03 (-0.18, 0.12) | 0.68 |
| **Household Income** | | | | | | | | | | | | | | | | | | |
| Above $75,000; population median | Reference category | | | | | | | | | | | | | | | | | |
| Below $75,000; population median | 0.01 (0, 0.03) | 0.04 | 0.03 (-0.09, 0.14) | 0.66 | 0.07 (-2.26, 2.41) | 0.95 | 0.20 (-1.63, 2.03) | 0.83 | 0.06 (-0.03, 0.15) | 0.22 | -0.02 (-0.09, 0.04) | 0.52 | 0.04 (-0.06, 0.13) | 0.48 | 0.04 (-0.02, 0.1) | 0.18 | 0.10 (-0.03, 0.22) | 0.14 |
| **Maternal Education** | | | | | | | | | | | | | | | | | | |
| Postgraduate Degree | Reference category | | | | | | | | | | | | | | | | | |
| Bachelor's Degree | 0.01 (-0.01, 0.02) | 0.31 | -0.03 (-0.17, 0.1) | 0.66 | 0.23 (-2.53, 2.98) | 0.87 | -0.78 (-3, 1.45) | 0.49 | 0.03 (-0.09, 0.15) | 0.60 | 0.01 (-0.07, 0.09) | 0.80 | -0.08 (-0.2, 0.04) | 0.21 | 0.00 (-0.07, 0.06) | 0.93 | 0.03 (-0.13, 0.19) | 0.73 |
| Trade/Diploma/Certificate/Other | 0.00 (-0.02, 0.02) | 1.00 | 0.00 (-0.15, 0.16) | 0.99 | 1.46 (-1.52, 4.45) | 0.34 | -0.27 (-2.73, 2.19) | 0.83 | -0.02 (-0.14, 0.11) | 0.78 | -0.01 (-0.1, 0.08) | 0.84 | 0.03 (-0.11, 0.17) | 0.66 | 0.00 (-0.08, 0.07) | 0.90 | -0.02 (-0.2, 0.15) | 0.80 |
| Less than Year 12 of high school | 0.00 (-0.02, 0.02) | 0.91 | 0.05 (-0.11, 0.21) | 0.57 | -1.21 (-4.4, 1.98) | 0.46 | -0.88 (-3.54, 1.78) | 0.52 | 0.08 (-0.06, 0.21) | 0.25 | 0.00 (-0.09, 0.1) | 0.93 | -0.03 (-0.17, 0.1) | 0.62 | 0.01 (-0.07, 0.09) | 0.85 | 0.09 (-0.1, 0.27) | 0.36 |
| ***Adjusted*** | | | | | | | | | | | | | | | | | | |
| **Neighbourhood Disadvantage** | | | | | | | | | | | | | | | | | | |
| High SEIFA | Reference category | | | | | | | | | | | | | | | | | |
| Medium SEIFA | 0.01 (0, 0.02) | 0.26 | 0.07 (-0.05, 0.2) | 0.23 | -3.45 (-6, -0.9) | 0.01 | -2.36 (-4.41, -0.31) | 0.02 | -0.02 (-0.12, 0.08) | 0.67 | 0.00 (-0.07, 0.07) | 0.97 | 0.01 (-0.09, 0.11) | 0.83 | 0.00 (-0.06, 0.06) | 0.91 | -0.03 (-0.17, 0.11) | 0.72 |
| Low SEIFA | 0.00 (-0.01, 0.01) | 0.82 | 0.16 (0.03, 0.29) | 0.01 | -1.77 (-4.31, 0.77) | 0.17 | -1.26 (-3.28, 0.76) | 0.22 | -0.03 (-0.14, 0.08) | 0.56 | -0.02 (-0.1, 0.05) | 0.51 | 0.02 (-0.09, 0.13) | 0.72 | -0.01 (-0.07, 0.05) | 0.75 | -0.04 (-0.19, 0.1) | 0.57 |
| **Household Income** | | | | | | | | | | | | | | | | | | |
| Above $75,000; population median | Reference category | | | | | | | | | | | | | | | | | |
| Below $75,000; population median | 0.01 (0, 0.03) | 0.03 | 0.03 (-0.09, 0.15) | 0.63 | -0.03 (-2.38, 2.32) | 0.98 | 0.15 (-1.7, 2) | 0.87 | 0.05 (-0.04, 0.14) | 0.26 | -0.02 (-0.09, 0.04) | 0.50 | 0.04 (-0.06, 0.14) | 0.45 | 0.04 (-0.02, 0.09) | 0.19 | 0.09 (-0.04, 0.22) | 0.16 |
| **Maternal Education** | | | | | | | | | | | | | | | | | | |
| Postgraduate Degree | Reference category | | | | | | | | | | | | | | | | | |
| Bachelor's Degree | 0.01 (-0.01, 0.02) | 0.31 | -0.02 (-0.16, 0.11) | 0.72 | 0.16 (-2.6, 2.92) | 0.91 | -0.83 (-3.06, 1.4) | 0.47 | 0.03 (-0.08, 0.15) | 0.58 | 0.01 (-0.07, 0.08) | 0.90 | -0.07 (-0.19, 0.05) | 0.25 | 0.00 (-0.07, 0.07) | 0.97 | 0.03 (-0.13, 0.19) | 0.70 |
| Trade/Diploma/Certificate/Other | 0.00 (-0.01, 0.02) | 0.96 | 0.00 (-0.16, 0.16) | 0.99 | 1.43 (-1.56, 4.43) | 0.35 | -0.27 (-2.73, 2.19) | 0.83 | -0.02 (-0.14, 0.1) | 0.74 | -0.01 (-0.09, 0.08) | 0.88 | 0.03 (-0.11, 0.17) | 0.68 | -0.01 (-0.08, 0.07) | 0.86 | -0.03 (-0.2, 0.15) | 0.76 |
| Less than Year 12 of high school | 0.00 (-0.02, 0.02) | 0.97 | 0.05 (-0.12, 0.21) | 0.57 | -1.39 (-4.6, 1.81) | 0.40 | -0.94 (-3.61, 1.74) | 0.49 | 0.07 (-0.06, 0.2) | 0.31 | 0.01 (-0.09, 0.1) | 0.88 | -0.04 (-0.17, 0.1) | 0.62 | 0.00 (-0.08, 0.08) | 0.93 | 0.07 (-0.11, 0.26) | 0.44 |

### Table S15. Effect modification (interaction test) by sex, Barwon Infant Study

| | **Max. cIMT (mm)** | | **PWV (m/s)** | | **Systolic Blood Pressure (mmHg)** | | **Diastolic Blood Pressure (mmHg)** | | **LDL-C (mmol/L)** | | **HDL-C (mmol/L)** | | **Total Triglycerides (mmol/L)** | | **Remnant Cholesterol (mmol/L)** | | **Non-HDL-C (mmol/L)** | |
|---|---|---|---|---|---|---|---|---|---|---|---|---|---|---|---|---|---|---|
| | Regression Coefficient (95% CI) | P Value | Regression Coefficient (95% CI) | P Value | Regression Coefficient (95% CI) | P Value | Regression Coefficient (95% CI) | P Value | Regression Coefficient (95% CI) | P Value | Regression Coefficient (95% CI) | P Value | Regression Coefficient (95% CI) | P Value | Regression Coefficient (95% CI) | P Value | Regression Coefficient (95% CI) | P Value |
| ***Unadjusted*** | | | | | | | | | | | | | | | | | | |
| **Neighbourhood Disadvantage** | | | | | | | | | | | | | | | | | | |
| High SEIFA | Reference category | | | | | | | | | | | | | | | | | |
| Medium SEIFA | 0.00 (-0.01, 0.02) | 0.70 | 0.07 (-0.1, 0.24) | 0.39 | -2.83 (-6.41, 0.75) | 0.12 | -1.26 (-3.93, 1.42) | 0.36 | 0.06 (-0.09, 0.2) | 0.44 | 0.00 (-0.09, 0.1) | 0.96 | 0.04 (-0.11, 0.19) | 0.63 | 0.06 (-0.03, 0.14) | 0.18 | 0.11 (-0.08, 0.31) | 0.25 |
| Low SEIFA | 0.01 (-0.01, 0.03) | 0.47 | 0.17 (-0.01, 0.35) | 0.06 | -0.80 (-4.44, 2.84) | 0.67 | -0.71 (-3.46, 2.04) | 0.61 | 0.02 (-0.13, 0.17) | 0.78 | -0.03 (-0.13, 0.06) | 0.50 | 0.05 (-0.1, 0.21) | 0.51 | 0.03 (-0.06, 0.12) | 0.47 | 0.05 (-0.15, 0.26) | 0.60 |
| **Household Income** | | | | | | | | | | | | | | | | | | |
| Above $75,000; population median | Reference category | | | | | | | | | | | | | | | | | |
| Below $75,000; population median | 0.02 (0, 0.04) | 0.02 | 0.01 (-0.16, 0.17) | 0.94 | 1.27 (-2.01, 4.55) | 0.45 | 0.44 (-2.02, 2.91) | 0.72 | 0.11 (-0.02, 0.24) | 0.09 | 0.01 (-0.08, 0.1) | 0.76 | 0.00 (-0.14, 0.14) | 0.99 | 0.08 (0, 0.16) | 0.06 | 0.19 (0.01, 0.37) | 0.04 |
| **Maternal Education** | | | | | | | | | | | | | | | | | | |
| Postgraduate Degree | Reference category | | | | | | | | | | | | | | | | | |
| Bachelor's Degree | 0.00 (-0.02, 0.02) | 0.96 | 0.00 (-0.19, 0.2) | 0.98 | -0.02 (-3.97, 3.94) | 0.99 | -1.73 (-4.74, 1.27) | 0.26 | 0.11 (-0.06, 0.28) | 0.21 | -0.02 (-0.13, 0.1) | 0.78 | -0.01 (-0.19, 0.18) | 0.94 | 0.06 (-0.04, 0.16) | 0.20 | 0.18 (-0.05, 0.4) | 0.13 |
| Trade/Diploma/Certificate/Other | -0.01 (-0.03, 0.02) | 0.65 | 0.04 (-0.17, 0.26) | 0.70 | 1.51 (-2.77, 5.8) | 0.49 | -0.99 (-4.32, 2.34) | 0.56 | 0.05 (-0.13, 0.23) | 0.56 | -0.02 (-0.15, 0.1) | 0.74 | 0.10 (-0.1, 0.29) | 0.33 | 0.06 (-0.04, 0.16) | 0.26 | 0.11 (-0.13, 0.36) | 0.36 |
| Less than Year 12 of high school | 0.00 (-0.03, 0.02) | 0.76 | 0.08 (-0.16, 0.32) | 0.50 | -1.15 (-5.74, 3.44) | 0.62 | -1.33 (-4.95, 2.29) | 0.47 | 0.19 (-0.02, 0.4) | 0.07 | -0.04 (-0.17, 0.1) | 0.59 | 0.01 (-0.22, 0.23) | 0.93 | 0.10 (-0.02, 0.22) | 0.09 | 0.29 (0.02, 0.57) | 0.04 |
| ***Adjusted*** | | | | | | | | | | | | | | | | | | |
| **Neighbourhood Disadvantage** | | | | | | | | | | | | | | | | | | |
| High SEIFA | Reference category | | | | | | | | | | | | | | | | | |
| Medium SEIFA | 0.00 (-0.01, 0.02) | 0.69 | 0.07 (-0.1, 0.24) | 0.41 | -2.93 (-6.52, 0.66) | 0.11 | -1.24 (-3.94, 1.45) | 0.36 | 0.05 (-0.09, 0.2) | 0.46 | 0.01 (-0.09, 0.1) | 0.89 | 0.03 (-0.12, 0.18) | 0.66 | 0.06 (-0.03, 0.14) | 0.19 | 0.11 (-0.08, 0.3) | 0.26 |
| Low SEIFA | 0.01 (-0.01, 0.03) | 0.44 | 0.17 (-0.01, 0.36) | 0.06 | -0.91 (-4.55, 2.74) | 0.63 | -0.69 (-3.46, 2.08) | 0.63 | 0.02 (-0.13, 0.17) | 0.83 | -0.03 (-0.13, 0.07) | 0.56 | 0.05 (-0.11, 0.21) | 0.55 | 0.03 (-0.06, 0.12) | 0.50 | 0.05 (-0.16, 0.25) | 0.64 |
| **Household Income** | | | | | | | | | | | | | | | | | | |
| Above $75,000; population median | Reference category | | | | | | | | | | | | | | | | | |
| Below $75,000; population median | 0.02 (0, 0.04) | 0.02 | 0.01 (-0.16, 0.17) | 0.95 | 1.23 (-2.06, 4.51) | 0.47 | 0.46 (-2.02, 2.94) | 0.72 | 0.11 (-0.02, 0.24) | 0.10 | 0.02 (-0.07, 0.11) | 0.72 | 0.00 (-0.15, 0.14) | 0.97 | 0.08 (0, 0.16) | 0.06 | 0.19 (0.01, 0.37) | 0.04 |
| **Maternal Education** | | | | | | | | | | | | | | | | | | |
| Postgraduate Degree | Reference category | | | | | | | | | | | | | | | | | |
| Bachelor's Degree | 0.00 (-0.02, 0.02) | 0.99 | 0.01 (-0.18, 0.21) | 0.90 | 0.01 (-3.94, 3.97) | 1.00 | -1.73 (-4.74, 1.27) | 0.26 | 0.11 (-0.06, 0.28) | 0.21 | -0.02 (-0.13, 0.1) | 0.76 | -0.01 (-0.19, 0.18) | 0.95 | 0.06 (-0.04, 0.16) | 0.21 | 0.17 (-0.06, 0.4) | 0.14 |
| Trade/Diploma/Certificate/Other | -0.01 (-0.03, 0.02) | 0.61 | 0.03 (-0.18, 0.25) | 0.77 | 1.48 (-2.82, 5.78) | 0.50 | -0.99 (-4.33, 2.36) | 0.56 | 0.05 (-0.13, 0.23) | 0.56 | -0.02 (-0.14, 0.11) | 0.76 | 0.10 (-0.1, 0.29) | 0.34 | 0.06 (-0.04, 0.17) | 0.26 | 0.11 (-0.13, 0.36) | 0.36 |
| Less than Year 12 of high school | 0.00 (-0.03, 0.02) | 0.78 | 0.09 (-0.16, 0.33) | 0.49 | -1.22 (-5.81, 3.37) | 0.60 | -1.31 (-4.93, 2.32) | 0.48 | 0.19 (-0.02, 0.4) | 0.07 | -0.03 (-0.17, 0.1) | 0.62 | 0.01 (-0.22, 0.23) | 0.95 | 0.10 (-0.02, 0.22) | 0.10 | 0.29 (0.02, 0.56) | 0.04 |

### Table S16. Financial security sensitivity analysis, Born in Bradford

| | Systolic Blood Pressure | | Diastolic Blood Pressure | | LDL-C | | HDL-C | | Total Triglycerides | | Non-HDL-C | | Remnant C | |
|---|---|---|---|---|---|---|---|---|---|---|---|---|---|---|
| | Regression Coefficient (95% CI) | P Value | Regression Coefficient (95% CI) | P Value | Regression Coefficient (95% CI) | P Value | Regression Coefficient (95% CI) | P Value | Regression Coefficient (95% CI) | P Value | Regression Coefficient (95% CI) | P Value | Regression Coefficient (95% CI) | P Value |
| **Financial Security (sensitivity)** | | | | | | | | | | | | | | |
| ***Unadjusted*** | | | | | | | | | | | | | | |
| Living comfortably | Reference category | | | | | | | | | | | | | |
| Doing alright | 0.30 (-0.67, 1.27) | 0.55 | 0.41 (-0.51, 1.32) | 0.38 | 0.01 (-0.04, 0.05) | 0.76 | -0.01 (-0.04, 0.02) | 0.55 | 0.02 (-0.06, 0.09) | 0.66 | 0.01 (-0.04, 0.06) | 0.70 | 0.00 (-0.02, 0.03) | 0.86 |
| Just about getting by | -0.55 (-1.66, 0.55) | 0.33 | -1.00 (-2.02, 0.01) | 0.05 | 0.01 (-0.05, 0.06) | 0.73 | 0.00 (-0.04, 0.04) | 0.99 | 0.02 (-0.07, 0.11) | 0.65 | 0.02 (-0.04, 0.07) | 0.57 | 0.01 (-0.02, 0.04) | 0.66 |
| Financially insecure | 0.76 (-0.94, 2.46) | 0.38 | 1.03 (-0.52, 2.59) | 0.19 | 0.01 (-0.06, 0.09) | 0.71 | -0.04 (-0.09, 0.02) | 0.17 | 0.00 (-0.13, 0.13) | 0.98 | 0.02 (-0.07, 0.1) | 0.71 | 0.00 (-0.04, 0.04) | 0.96 |
| ***Adjusted*** | | | | | | | | | | | | | | |
| Living comfortably | Reference category | | | | | | | | | | | | | |
| Doing alright | 0.29 (-0.68, 1.26) | 0.56 | 0.38 (-0.53, 1.3) | 0.41 | 0.01 (-0.04, 0.05) | 0.77 | -0.01 (-0.04, 0.02) | 0.60 | 0.02 (-0.06, 0.09) | 0.64 | 0.01 (-0.04, 0.06) | 0.73 | 0.00 (-0.02, 0.03) | 0.89 |
| Just about getting by | -0.49 (-1.6, 0.61) | 0.38 | -1.03 (-2.04, -0.02) | 0.05 | 0.01 (-0.05, 0.06) | 0.74 | 0.00 (-0.04, 0.04) | 0.96 | 0.02 (-0.07, 0.1) | 0.66 | 0.02 (-0.04, 0.07) | 0.58 | 0.01 (-0.02, 0.03) | 0.68 |
| Financially insecure | 0.85 (-0.85, 2.55) | 0.33 | 0.87 (-0.68, 2.43) | 0.27 | 0.01 (-0.06, 0.09) | 0.74 | -0.04 (-0.09, 0.01) | 0.15 | 0.00 (-0.13, 0.14) | 0.97 | 0.01 (-0.07, 0.1) | 0.76 | 0.00 (-0.04, 0.04) | 0.99 |

### Table S17. Subgroup results in girls, Avon Longitudinal Study of Parents and Children

| | Max. cIMT (mm) | | PWV (m/s) | | Systolic Blood Pressure (mmHg) | | Diastolic Blood Pressure (mmHg) | | LDL-C (mmol/L) | | HDL-C (mmol/L) | | Total Triglycerides (mmol/L) | | Remnant Cholesterol (mmol/L) | | Non-HDL-C (mmol/L) | |
|---|---|---|---|---|---|---|---|---|---|---|---|---|---|---|---|---|---|---|
| | Regression Coefficient (95% CI) | P Value | Regression Coefficient (95% CI) | P Value | Regression Coefficient (95% CI) | P Value | Regression Coefficient (95% CI) | P Value | Regression Coefficient (95% CI) | P Value | Regression Coefficient (95% CI) | P Value | Regression Coefficient (95% CI) | P Value | Regression Coefficient (95% CI) | P Value | Regression Coefficient (95% CI) | P Value |
| ***Unadjusted*** | | | | | | | | | | | | | | | | | | |
| **Neighbourhood Disadvantage (IMD 2000)** | | | | | | | | | | | | | | | | | | |
| Low Deprivation | Reference category | | | | | | | | | | | | | | | | | |
| Medium Deprivation | 0.00 (-0.01, 0) | 0.46 | 0.03 (-0.04, 0.1) | 0.43 | 0.51 (-0.46, 1.49) | 0.30 | 0.33 (-0.36, 1.02) | 0.35 | -0.01 (-0.06, 0.05) | 0.85 | -0.02 (-0.06, 0.02) | 0.25 | 0.03 (-0.04, 0.1) | 0.38 | 0.00 (-0.03, 0.03) | 0.82 | 0.00 (-0.07, 0.06) | 0.94 |
| High Deprivation | -0.01 (-0.01, 0) | 0.02 | 0.06 (-0.01, 0.13) | 0.09 | 0.92 (-0.07, 1.9) | 0.07 | 0.89 (0.16, 1.61) | 0.02 | -0.02 (-0.08, 0.04) | 0.52 | -0.03 (-0.07, 0.01) | 0.12 | 0.02 (-0.05, 0.09) | 0.66 | 0.00 (-0.03, 0.04) | 0.88 | -0.02 (-0.09, 0.05) | 0.63 |
| **Financial Difficulties** | | | | | | | | | | | | | | | | | | |
| No Difficulties | Reference category | | | | | | | | | | | | | | | | | |
| Difficulties | 0.00 (-0.01, 0) | 0.22 | 0.05 (-0.01, 0.11) | 0.11 | 0.66 (-0.1, 1.43) | 0.09 | 0.23 (-0.32, 0.78) | 0.42 | -0.01 (-0.05, 0.04) | 0.80 | -0.02 (-0.05, 0.01) | 0.26 | 0.01 (-0.05, 0.06) | 0.81 | 0.00 (-0.03, 0.03) | 0.93 | 0.00 (-0.06, 0.05) | 0.86 |
| **Maternal Education** | | | | | | | | | | | | | | | | | | |
| University Degree | Reference category | | | | | | | | | | | | | | | | | |
| A Levels | 0.00 (-0.01, 0) | 0.24 | -0.01 (-0.09, 0.07) | 0.79 | 0.60 (-0.47, 1.67) | 0.28 | 0.54 (-0.25, 1.32) | 0.18 | 0.00 (-0.07, 0.07) | 0.93 | -0.01 (-0.06, 0.04) | 0.68 | 0.04 (-0.04, 0.12) | 0.33 | 0.00 (-0.04, 0.04) | 0.88 | 0.01 (-0.07, 0.09) | 0.88 |
| O Levels | 0.00 (-0.01, 0) | 0.15 | 0.06 (-0.01, 0.14) | 0.11 | 1.72 (0.7, 2.75) | 0.00 | 1.35 (0.59, 2.11) | 0.00 | -0.01 (-0.08, 0.06) | 0.81 | -0.02 (-0.07, 0.02) | 0.29 | 0.07 (-0.01, 0.14) | 0.08 | 0.00 (-0.03, 0.04) | 0.89 | -0.01 (-0.08, 0.07) | 0.88 |
| Vocational/CSE | -0.01 (-0.02, 0) | 0.00 | 0.13 (0.04, 0.22) | 0.00 | 2.94 (1.78, 4.1) | 0.00 | 2.45 (1.6, 3.31) | 0.00 | -0.01 (-0.09, 0.07) | 0.79 | -0.05 (-0.1, 0.01) | 0.08 | 0.07 (-0.02, 0.16) | 0.12 | 0.01 (-0.03, 0.05) | 0.64 | 0.00 (-0.09, 0.09) | 0.99 |
| ***Adjusted*** | | | | | | | | | | | | | | | | | | |
| **Neighbourhood Disadvantage (IMD 2000)** | | | | | | | | | | | | | | | | | | |
| Low Deprivation | Reference category | | | | | | | | | | | | | | | | | |
| Medium Deprivation | 0.00 (-0.01, 0) | 0.46 | 0.02 (-0.05, 0.1) | 0.49 | 0.54 (-0.43, 1.51) | 0.27 | 0.29 (-0.4, 0.98) | 0.41 | -0.01 (-0.06, 0.05) | 0.85 | -0.02 (-0.06, 0.02) | 0.25 | 0.03 (-0.04, 0.1) | 0.40 | 0.00 (-0.03, 0.04) | 0.81 | 0.00 (-0.07, 0.06) | 0.96 |
| High Deprivation | -0.01 (-0.01, 0) | 0.01 | 0.05 (-0.02, 0.12) | 0.16 | 1.03 (0.04, 2.01) | 0.04 | 0.79 (0.06, 1.52) | 0.03 | -0.02 (-0.08, 0.04) | 0.48 | -0.03 (-0.07, 0.01) | 0.12 | 0.02 (-0.06, 0.09) | 0.66 | 0.00 (-0.03, 0.04) | 0.90 | -0.02 (-0.09, 0.05) | 0.58 |
| **Financial Difficulties** | | | | | | | | | | | | | | | | | | |
| No Difficulties | Reference category | | | | | | | | | | | | | | | | | |
| Difficulties | 0.00 (-0.01, 0) | 0.17 | 0.04 (-0.02, 0.1) | 0.19 | 0.75 (-0.02, 1.52) | 0.06 | 0.14 (-0.41, 0.7) | 0.61 | -0.01 (-0.06, 0.04) | 0.74 | -0.02 (-0.05, 0.01) | 0.25 | 0.01 (-0.05, 0.06) | 0.78 | 0.00 (-0.03, 0.03) | 0.96 | -0.01 (-0.06, 0.05) | 0.79 |
| **Maternal Education** | | | | | | | | | | | | | | | | | | |
| University Degree | Reference category | | | | | | | | | | | | | | | | | |
| A Levels | 0.00 (-0.01, 0) | 0.23 | -0.02 (-0.1, 0.07) | 0.70 | 0.64 (-0.43, 1.71) | 0.24 | 0.49 (-0.29, 1.27) | 0.22 | 0.00 (-0.07, 0.07) | 0.92 | -0.01 (-0.06, 0.04) | 0.66 | 0.04 (-0.04, 0.12) | 0.35 | 0.00 (-0.04, 0.04) | 0.87 | 0.01 (-0.07, 0.09) | 0.87 |
| O Levels | 0.00 (-0.01, 0) | 0.16 | 0.06 (-0.02, 0.13) | 0.15 | 1.77 (0.75, 2.8) | 0.00 | 1.28 (0.53, 2.04) | 0.00 | -0.01 (-0.07, 0.06) | 0.86 | -0.03 (-0.07, 0.02) | 0.28 | 0.06 (-0.01, 0.13) | 0.10 | 0.00 (-0.03, 0.04) | 0.84 | 0.00 (-0.08, 0.07) | 0.94 |
| Vocational/CSE | -0.01 (-0.02, 0) | 0.00 | 0.12 (0.03, 0.21) | 0.01 | 3.07 (1.91, 4.23) | 0.00 | 2.32 (1.47, 3.17) | 0.00 | -0.01 (-0.09, 0.07) | 0.80 | -0.05 (-0.1, 0) | 0.07 | 0.06 (-0.02, 0.15) | 0.15 | 0.01 (-0.03, 0.05) | 0.61 | 0.00 (-0.09, 0.09) | 0.98 |

### Table S18. Subgroup results in boys, Avon Longitudinal Study of Parents and Children

| | Max. cIMT (mm) | | PWV (m/s) | | Systolic Blood Pressure (mmHg) | | Diastolic Blood Pressure (mmHg) | | LDL-C (mmol/L) | | HDL-C (mmol/L) | | Total Triglycerides (mmol/L) | | Remnant Cholesterol (mmol/L) | | Non-HDL-C (mmol/L) | |
|---|---|---|---|---|---|---|---|---|---|---|---|---|---|---|---|---|---|---|
| | Regression Coefficient (95% CI) | P Value | Regression Coefficient (95% CI) | P Value | Regression Coefficient (95% CI) | P Value | Regression Coefficient (95% CI) | P Value | Regression Coefficient (95% CI) | P Value | Regression Coefficient (95% CI) | P Value | Regression Coefficient (95% CI) | P Value | Regression Coefficient (95% CI) | P Value | Regression Coefficient (95% CI) | P Value |
| ***Unadjusted*** | | | | | | | | | | | | | | | | | | |
| **Neighbourhood Disadvantage (IMD 2000)** | | | | | | | | | | | | | | | | | | |
| Low Deprivation | Reference category | | | | | | | | | | | | | | | | | |
| Medium Deprivation | 0.00 (-0.01, 0.01) | 0.92 | 0.02 (-0.06, 0.11) | 0.62 | 0.96 (-0.2, 2.13) | 0.10 | 0.66 (-0.1, 1.43) | 0.09 | -0.01 (-0.06, 0.05) | 0.80 | 0.00 (-0.04, 0.04) | 1.00 | 0.02 (-0.05, 0.08) | 0.63 | -0.01 (-0.04, 0.03) | 0.76 | -0.01 (-0.08, 0.06) | 0.73 |
| High Deprivation | 0.00 (-0.01, 0.01) | 0.73 | 0.06 (-0.04, 0.16) | 0.25 | 0.70 (-0.65, 2.05) | 0.31 | 1.02 (0.13, 1.91) | 0.03 | -0.01 (-0.07, 0.05) | 0.76 | -0.01 (-0.05, 0.03) | 0.60 | 0.04 (-0.04, 0.11) | 0.33 | 0.01 (-0.03, 0.04) | 0.74 | 0.00 (-0.08, 0.07) | 0.93 |
| **Financial Difficulties** | | | | | | | | | | | | | | | | | | |
| No Difficulties | Reference category | | | | | | | | | | | | | | | | | |
| Difficulties | 0.00 (-0.01, 0) | 0.42 | 0.04 (-0.04, 0.11) | 0.33 | 0.62 (-0.38, 1.62) | 0.23 | 0.45 (-0.2, 1.1) | 0.17 | -0.02 (-0.07, 0.03) | 0.48 | -0.01 (-0.04, 0.02) | 0.50 | -0.01 (-0.07, 0.05) | 0.74 | -0.01 (-0.04, 0.01) | 0.31 | -0.03 (-0.09, 0.03) | 0.29 |
| **Maternal Education** | | | | | | | | | | | | | | | | | | |
| University Degree | Reference category | | | | | | | | | | | | | | | | | |
| A Levels | -0.01 (-0.01, 0) | 0.08 | 0.03 (-0.07, 0.13) | 0.58 | 1.04 (-0.33, 2.4) | 0.14 | 0.52 (-0.34, 1.38) | 0.23 | -0.02 (-0.09, 0.04) | 0.48 | 0.00 (-0.04, 0.04) | 0.91 | 0.01 (-0.07, 0.08) | 0.84 | -0.02 (-0.05, 0.02) | 0.41 | -0.04 (-0.12, 0.04) | 0.33 |
| O Levels | 0.00 (-0.01, 0) | 0.33 | 0.07 (-0.03, 0.16) | 0.16 | 0.55 (-0.8, 1.89) | 0.43 | 0.71 (-0.13, 1.55) | 0.10 | -0.03 (-0.09, 0.04) | 0.44 | 0.00 (-0.04, 0.04) | 0.93 | 0.03 (-0.05, 0.11) | 0.42 | -0.02 (-0.05, 0.02) | 0.41 | -0.04 (-0.12, 0.04) | 0.30 |
| Vocational/CSE | 0.00 (-0.01, 0.01) | 0.57 | 0.04 (-0.07, 0.16) | 0.43 | 1.33 (-0.17, 2.84) | 0.08 | 1.05 (0.1, 2.01) | 0.03 | 0.01 (-0.07, 0.09) | 0.80 | 0.00 (-0.05, 0.05) | 0.99 | 0.04 (-0.05, 0.13) | 0.37 | 0.01 (-0.03, 0.06) | 0.55 | 0.02 (-0.07, 0.11) | 0.62 |
| ***Adjusted*** | | | | | | | | | | | | | | | | | | |
| **Neighbourhood Disadvantage (IMD 2000)** | | | | | | | | | | | | | | | | | | |
| Low Deprivation | Reference category | | | | | | | | | | | | | | | | | |
| Medium Deprivation | 0.00 (-0.01, 0.01) | 0.91 | 0.01 (-0.07, 0.1) | 0.77 | 0.97 (-0.2, 2.13) | 0.10 | 0.55 (-0.22, 1.32) | 0.16 | -0.01 (-0.07, 0.05) | 0.75 | 0.00 (-0.04, 0.04) | 0.97 | 0.01 (-0.06, 0.08) | 0.72 | -0.01 (-0.04, 0.03) | 0.72 | -0.02 (-0.09, 0.05) | 0.66 |
| High Deprivation | 0.00 (-0.01, 0.01) | 0.73 | 0.04 (-0.06, 0.14) | 0.38 | 0.76 (-0.6, 2.11) | 0.27 | 0.84 (-0.05, 1.74) | 0.06 | -0.01 (-0.07, 0.05) | 0.68 | -0.01 (-0.06, 0.03) | 0.50 | 0.03 (-0.04, 0.11) | 0.37 | 0.01 (-0.03, 0.04) | 0.74 | -0.01 (-0.08, 0.07) | 0.86 |
| **Financial Difficulties** | | | | | | | | | | | | | | | | | | |
| No Difficulties | Reference category | | | | | | | | | | | | | | | | | |
| Difficulties | 0.00 (-0.01, 0) | 0.42 | 0.03 (-0.05, 0.1) | 0.48 | 0.68 (-0.33, 1.68) | 0.19 | 0.31 (-0.33, 0.96) | 0.34 | -0.02 (-0.07, 0.03) | 0.42 | -0.01 (-0.05, 0.02) | 0.40 | -0.01 (-0.07, 0.05) | 0.70 | -0.01 (-0.04, 0.01) | 0.33 | -0.03 (-0.09, 0.03) | 0.26 |
| **Maternal Education** | | | | | | | | | | | | | | | | | | |
| University Degree | Reference category | | | | | | | | | | | | | | | | | |
| A Levels | -0.01 (-0.01, 0) | 0.08 | 0.03 (-0.07, 0.13) | 0.60 | 1.06 (-0.31, 2.43) | 0.13 | 0.51 (-0.35, 1.36) | 0.24 | -0.02 (-0.09, 0.04) | 0.47 | 0.00 (-0.04, 0.04) | 0.94 | 0.01 (-0.07, 0.08) | 0.83 | -0.01 (-0.05, 0.02) | 0.43 | -0.04 (-0.12, 0.04) | 0.33 |
| O Levels | 0.00 (-0.01, 0) | 0.33 | 0.06 (-0.03, 0.16) | 0.20 | 0.55 (-0.8, 1.9) | 0.42 | 0.63 (-0.2, 1.47) | 0.14 | -0.03 (-0.09, 0.04) | 0.41 | 0.00 (-0.05, 0.04) | 0.90 | 0.03 (-0.05, 0.11) | 0.46 | -0.02 (-0.05, 0.02) | 0.40 | -0.04 (-0.12, 0.04) | 0.28 |
| Vocational/CSE | 0.00 (-0.01, 0.01) | 0.57 | 0.03 (-0.08, 0.14) | 0.58 | 1.36 (-0.15, 2.88) | 0.08 | 0.88 (-0.07, 1.83) | 0.07 | 0.01 (-0.07, 0.08) | 0.86 | 0.00 (-0.05, 0.05) | 0.93 | 0.04 (-0.05, 0.13) | 0.42 | 0.01 (-0.03, 0.06) | 0.56 | 0.02 (-0.07, 0.11) | 0.67 |

### Table S19. Effect modification (interaction test) by sex, Avon Longitudinal Study of Parents and Children

| | Max. cIMT (mm) | | PWV (m/s) | | Systolic Blood Pressure (mmHg) | | Diastolic Blood Pressure (mmHg) | | LDL-C (mmol/L) | | HDL-C (mmol/L) | | Total Triglycerides (mmol/L) | | Remnant Cholesterol (mmol/L) | | Non-HDL-C (mmol/L) | |
|---|---|---|---|---|---|---|---|---|---|---|---|---|---|---|---|---|---|---|
| | Regression Coefficient (95% CI) | P Value | Regression Coefficient (95% CI) | P Value | Regression Coefficient (95% CI) | P Value | Regression Coefficient (95% CI) | P Value | Regression Coefficient (95% CI) | P Value | Regression Coefficient (95% CI) | P Value | Regression Coefficient (95% CI) | P Value | Regression Coefficient (95% CI) | P Value | Regression Coefficient (95% CI) | P Value |
| ***Unadjusted*** | | | | | | | | | | | | | | | | | | |
| **Neighbourhood Disadvantage (IMD 2000)** | | | | | | | | | | | | | | | | | | |
| Low Deprivation | Reference category | | | | | | | | | | | | | | | | | |
| Medium Deprivation | 0.00 (-0.01, 0.01) | 0.67 | -0.01 (-0.12, 0.1) | 0.90 | 0.45 (-1.01, 1.92) | 0.55 | 0.33 (-0.65, 1.32) | 0.51 | 0.00 (-0.08, 0.08) | 0.97 | 0.02 (-0.03, 0.08) | 0.40 | -0.01 (-0.11, 0.08) | 0.76 | -0.01 (-0.05, 0.04) | 0.70 | -0.01 (-0.1, 0.08) | 0.83 |
| High Deprivation | 0.01 (0, 0.01) | 0.26 | 0.00 (-0.12, 0.11) | 0.94 | -0.22 (-1.8, 1.37) | 0.79 | 0.13 (-0.99, 1.26) | 0.81 | 0.01 (-0.07, 0.09) | 0.81 | 0.02 (-0.04, 0.08) | 0.50 | 0.02 (-0.08, 0.12) | 0.68 | 0.00 (-0.05, 0.05) | 0.89 | 0.01 (-0.09, 0.11) | 0.79 |
| **Financial Difficulties** | | | | | | | | | | | | | | | | | | |
| No Difficulties | Reference category | | | | | | | | | | | | | | | | | |
| Difficulties | 0.00 (-0.01, 0.01) | 0.88 | -0.01 (-0.1, 0.08) | 0.78 | -0.05 (-1.28, 1.19) | 0.94 | 0.22 (-0.62, 1.06) | 0.60 | -0.01 (-0.08, 0.05) | 0.71 | 0.01 (-0.04, 0.06) | 0.76 | -0.02 (-0.09, 0.06) | 0.67 | -0.01 (-0.05, 0.02) | 0.44 | -0.03 (-0.11, 0.05) | 0.50 |
| **Maternal Education** | | | | | | | | | | | | | | | | | | |
| University Degree | Reference category | | | | | | | | | | | | | | | | | |
| A Levels | 0.00 (-0.01, 0.01) | 0.56 | 0.04 (-0.09, 0.16) | 0.54 | 0.44 (-1.29, 2.17) | 0.62 | -0.01 (-1.17, 1.14) | 0.98 | -0.03 (-0.12, 0.07) | 0.59 | 0.01 (-0.05, 0.08) | 0.70 | -0.03 (-0.14, 0.07) | 0.56 | -0.02 (-0.07, 0.04) | 0.50 | -0.04 (-0.16, 0.07) | 0.44 |
| O Levels | 0.00 (-0.01, 0.01) | 0.85 | 0.00 (-0.12, 0.12) | 0.95 | -1.18 (-2.83, 0.47) | 0.16 | -0.64 (-1.77, 0.48) | 0.26 | -0.02 (-0.12, 0.08) | 0.73 | 0.02 (-0.04, 0.09) | 0.49 | -0.03 (-0.14, 0.07) | 0.54 | -0.02 (-0.07, 0.03) | 0.48 | -0.04 (-0.15, 0.08) | 0.55 |
| Vocational/CSE | 0.01 (0, 0.02) | 0.09 | -0.09 (-0.23, 0.05) | 0.23 | -1.61 (-3.47, 0.26) | 0.09 | -1.40 (-2.66, -0.14) | 0.03 | 0.02 (-0.09, 0.13) | 0.72 | 0.05 (-0.02, 0.12) | 0.19 | -0.03 (-0.15, 0.09) | 0.66 | 0.00 (-0.06, 0.06) | 0.91 | 0.02 (-0.1, 0.15) | 0.71 |
| ***Adjusted*** | | | | | | | | | | | | | | | | | | |
| **Neighbourhood Disadvantage (IMD 2000)** | | | | | | | | | | | | | | | | | | |
| Low Deprivation | Reference category | | | | | | | | | | | | | | | | | |
| Medium Deprivation | 0.00 (-0.01, 0.01) | 0.67 | -0.01 (-0.12, 0.1) | 0.83 | 0.46 (-1, 1.93) | 0.54 | 0.28 (-0.7, 1.27) | 0.57 | 0.00 (-0.08, 0.08) | 0.98 | 0.02 (-0.03, 0.08) | 0.41 | -0.02 (-0.11, 0.08) | 0.72 | -0.01 (-0.05, 0.04) | 0.70 | -0.01 (-0.1, 0.08) | 0.83 |
| High Deprivation | 0.01 (0, 0.01) | 0.25 | -0.01 (-0.13, 0.11) | 0.88 | -0.22 (-1.8, 1.36) | 0.79 | 0.08 (-1.04, 1.2) | 0.89 | 0.01 (-0.07, 0.09) | 0.81 | 0.02 (-0.04, 0.08) | 0.50 | 0.02 (-0.09, 0.12) | 0.74 | 0.00 (-0.05, 0.05) | 0.89 | 0.01 (-0.09, 0.11) | 0.78 |
| **Financial Difficulties** | | | | | | | | | | | | | | | | | | |
| No Difficulties | Reference category | | | | | | | | | | | | | | | | | |
| Difficulties | 0.00 (-0.01, 0.01) | 0.87 | -0.02 (-0.11, 0.07) | 0.72 | -0.05 (-1.28, 1.19) | 0.94 | 0.18 (-0.65, 1.02) | 0.67 | -0.01 (-0.08, 0.05) | 0.72 | 0.01 (-0.04, 0.06) | 0.76 | -0.02 (-0.1, 0.06) | 0.62 | -0.01 (-0.05, 0.02) | 0.44 | -0.03 (-0.11, 0.05) | 0.50 |
| **Maternal Education** | | | | | | | | | | | | | | | | | | |
| University Degree | Reference category | | | | | | | | | | | | | | | | | |
| A Levels | 0.00 (-0.01, 0.01) | 0.56 | 0.04 (-0.08, 0.17) | 0.51 | 0.44 (-1.29, 2.16) | 0.62 | 0.02 (-1.13, 1.17) | 0.97 | -0.03 (-0.12, 0.07) | 0.59 | 0.01 (-0.05, 0.08) | 0.69 | -0.03 (-0.14, 0.08) | 0.59 | -0.02 (-0.07, 0.04) | 0.50 | -0.05 (-0.16, 0.07) | 0.44 |
| O Levels | 0.00 (-0.01, 0.01) | 0.86 | 0.01 (-0.11, 0.12) | 0.93 | -1.17 (-2.82, 0.48) | 0.16 | -0.62 (-1.74, 0.5) | 0.28 | -0.02 (-0.12, 0.08) | 0.72 | 0.02 (-0.04, 0.09) | 0.49 | -0.03 (-0.14, 0.07) | 0.57 | -0.02 (-0.07, 0.03) | 0.48 | -0.04 (-0.15, 0.08) | 0.54 |
| Vocational/CSE | 0.01 (0, 0.02) | 0.09 | -0.08 (-0.22, 0.06) | 0.24 | -1.62 (-3.49, 0.24) | 0.09 | -1.38 (-2.64, -0.13) | 0.03 | 0.02 (-0.09, 0.13) | 0.71 | 0.05 (-0.02, 0.12) | 0.19 | -0.03 (-0.15, 0.09) | 0.66 | 0.00 (-0.06, 0.06) | 0.92 | 0.02 (-0.1, 0.15) | 0.71 |

### Table S20. Complete case results, Barwon Infant Study

| | Max. cIMT (mm) | | PWV (m/s) | | Systolic Blood Pressure (mmHg) | | Diastolic Blood Pressure (mmHg) | | LDL-C (mmol/L) | | HDL-C (mmol/L) | | Total Triglycerides (mmol/L) | | Remnant Cholesterol (mmol/L) | | Non-HDL-C (mmol/L) | |
|---|---|---|---|---|---|---|---|---|---|---|---|---|---|---|---|---|---|---|
| | Regression Coefficient (95% CI) | P Value | Regression Coefficient (95% CI) | P Value | Regression Coefficient (95% CI) | P Value | Regression Coefficient (95% CI) | P Value | Regression Coefficient (95% CI) | P Value | Regression Coefficient (95% CI) | P Value | Regression Coefficient (95% CI) | P Value | Regression Coefficient (95% CI) | P Value | Regression Coefficient (95% CI) | P Value |
| | | | | | | | | | | | *Unadjusted* | | | | | | | |
| **Neighbourhood Disadvantage** | | | | | | | | | | | | | | | | | | |
| High SEIFA | Reference category | | | | | | | | | | | | | | | | | |
| Medium SEIFA | 0.01 (0, 0.01) | 0.22 | 0.04 (-0.05, 0.12) | 0.42 | -1.95 (-3.7, -0.2) | 0.03 | -1.70 (-3.07, -0.34) | 0.01 | -0.07 (-0.13, 0) | 0.05 | 0.00 (-0.05, 0.05) | 0.89 | -0.01 (-0.07, 0.05) | 0.72 | -0.05 (-0.09, 0) | 0.06 | -0.11 (-0.22, -0.01) | 0.04 |
| Low SEIFA | 0.00 (-0.01, 0.01) | 0.46 | 0.09 (-0.01, 0.18) | 0.07 | -1.17 (-3, 0.65) | 0.21 | -0.87 (-2.29, 0.56) | 0.23 | -0.05 (-0.12, 0.01) | 0.12 | 0.00 (-0.06, 0.05) | 0.85 | -0.01 (-0.07, 0.05) | 0.80 | -0.04 (-0.09, 0.01) | 0.12 | -0.09 (-0.21, 0.02) | 0.10 |
| **Household Income** | | | | | | | | | | | | | | | | | | |
| Above $75,000; population median | Reference category | | | | | | | | | | | | | | | | | |
| Below $75,000; population median | 0.00 (0, 0.01) | 0.40 | 0.02 (-0.06, 0.1) | 0.62 | -0.46 (-2.12, 1.2) | 0.59 | 0.11 (-1.2, 1.41) | 0.87 | 0.01 (-0.06, 0.07) | 0.83 | -0.03 (-0.08, 0.02) | 0.23 | 0.05 (-0.01, 0.11) | 0.10 | 0.01 (-0.04, 0.05) | 0.73 | 0.02 (-0.09, 0.12) | 0.78 |
| **Maternal Education** | | | | | | | | | | | | | | | | | | |
| Postgraduate Degree | Reference category | | | | | | | | | | | | | | | | | |
| Bachelor's Degree | 0.01 (0, 0.02) | 0.14 | -0.03 (-0.13, 0.07) | 0.56 | -0.12 (-2.11, 1.87) | 0.91 | -0.15 (-1.71, 1.4) | 0.85 | -0.04 (-0.11, 0.04) | 0.36 | 0.02 (-0.04, 0.07) | 0.55 | -0.08 (-0.15, -0.01) | 0.03 | -0.04 (-0.1, 0.01) | 0.12 | -0.08 (-0.2, 0.05) | 0.22 |
| Trade/Diploma/Certificate/Other | 0.00 (-0.01, 0.01) | 0.75 | -0.02 (-0.13, 0.09) | 0.70 | 0.33 (-1.87, 2.53) | 0.77 | -0.08 (-1.81, 1.64) | 0.92 | -0.06 (-0.15, 0.02) | 0.17 | -0.01 (-0.07, 0.06) | 0.83 | -0.02 (-0.1, 0.05) | 0.57 | -0.04 (-0.1, 0.02) | 0.15 | -0.10 (-0.24, 0.03) | 0.14 |
| Less than Year 12 of high school | 0.00 (-0.01, 0.01) | 0.99 | 0.01 (-0.11, 0.13) | 0.86 | -0.42 (-2.84, 2) | 0.73 | -0.17 (-2.06, 1.72) | 0.86 | -0.02 (-0.11, 0.08) | 0.74 | 0.02 (-0.04, 0.09) | 0.48 | -0.06 (-0.14, 0.02) | 0.15 | -0.05 (-0.12, 0.01) | 0.09 | -0.07 (-0.22, 0.08) | 0.35 |
| | | | | | | | | | | | *Adjusted* | | | | | | | |
| **Neighbourhood Disadvantage** | | | | | | | | | | | | | | | | | | |
| High SEIFA | Reference category | | | | | | | | | | | | | | | | | |
| Medium SEIFA | 0.00 (0, 0.01) | 0.30 | 0.03 (-0.06, 0.12) | 0.51 | -2.16 (-3.98, -0.34) | 0.02 | -1.88 (-3.29, -0.46) | 0.01 | -0.06 (-0.13, 0.01) | 0.07 | 0.00 (-0.05, 0.05) | 0.92 | 0.01 (-0.05, 0.07) | 0.80 | -0.03 (-0.08, 0.01) | 0.16 | -0.10 (-0.21, 0.01) | 0.09 |
| Low SEIFA | -0.01 (-0.02, 0) | 0.27 | 0.09 (-0.01, 0.18) | 0.07 | -1.44 (-3.35, 0.47) | 0.14 | -1.03 (-2.51, 0.45) | 0.17 | -0.06 (-0.13, 0.02) | 0.12 | -0.01 (-0.06, 0.04) | 0.69 | 0.00 (-0.06, 0.07) | 0.92 | -0.03 (-0.08, 0.02) | 0.19 | -0.09 (-0.21, 0.03) | 0.13 |
| **Household Income** | | | | | | | | | | | | | | | | | | |
| Above $75,000; population median | Reference category | | | | | | | | | | | | | | | | | |
| Below $75,000; population median | 0.00 (-0.01, 0.01) | 0.59 | 0.02 (-0.06, 0.11) | 0.59 | -0.23 (-1.95, 1.49) | 0.79 | 0.27 (-1.08, 1.61) | 0.70 | -0.01 (-0.07, 0.06) | 0.86 | -0.03 (-0.08, 0.01) | 0.17 | 0.04 (-0.02, 0.1) | 0.15 | 0.00 (-0.05, 0.05) | 0.99 | -0.01 (-0.11, 0.1) | 0.92 |
| **Maternal Education** | | | | | | | | | | | | | | | | | | |
| Postgraduate Degree | Reference category | | | | | | | | | | | | | | | | | |
| Bachelor's Degree | 0.01 (0, 0.02) | 0.18 | 0.00 (-0.11, 0.1) | 0.94 | 0.12 (-1.96, 2.2) | 0.91 | 0.02 (-1.6, 1.64) | 0.98 | -0.03 (-0.11, 0.05) | 0.45 | 0.02 (-0.04, 0.07) | 0.58 | -0.08 (-0.15, -0.01) | 0.02 | -0.04 (-0.1, 0.01) | 0.15 | -0.07 (-0.2, 0.06) | 0.28 |
| Trade/Diploma/Certificate/Other | 0.00 (-0.01, 0.01) | 0.73 | 0.02 (-0.1, 0.13) | 0.77 | 0.38 (-1.91, 2.67) | 0.75 | 0.18 (-1.6, 1.96) | 0.84 | -0.07 (-0.16, 0.02) | 0.13 | -0.01 (-0.08, 0.05) | 0.73 | -0.03 (-0.11, 0.05) | 0.41 | -0.05 (-0.11, 0.01) | 0.10 | -0.12 (-0.26, 0.02) | 0.10 |
| Less than Year 12 of high school | 0.00 (-0.01, 0.01) | 0.91 | 0.05 (-0.08, 0.17) | 0.46 | -0.44 (-2.94, 2.05) | 0.73 | 0.02 (-1.92, 1.96) | 0.98 | -0.03 (-0.12, 0.07) | 0.58 | 0.02 (-0.05, 0.09) | 0.58 | -0.06 (-0.15, 0.02) | 0.14 | -0.06 (-0.13, 0) | 0.07 | -0.09 (-0.24, 0.07) | 0.26 |

### Table S21. Complete case results, Born in Bradford

| | Systolic Blood Pressure | | Diastolic Blood Pressure | | LDL-C | | HDL-C | | Total Triglycerides | | Non-HDL-C | | Remnant C | |
|---|---|---|---|---|---|---|---|---|---|---|---|---|---|---|
| | Regression Coefficient (95% CI) | P Value | Regression Coefficient (95% CI) | P Value | Regression Coefficient (95% CI) | P Value | Regression Coefficient (95% CI) | P Value | Regression Coefficient (95% CI) | P Value | Regression Coefficient (95% CI) | P Value | Regression Coefficient (95% CI) | P Value |
| ***Unadjusted*** | | | | | | | | | | | | | | |
| **Neighbourhood Deprivation (IMD)** | | | | | | | | | | | | | | |
| Low Deprivation | Reference category | | | | | | | | | | | | | |
| Medium Deprivation | -0.42 (-1.37, 0.54) | 0.39 | 0.10 (-0.79, 0.99) | 0.83 | 0.01 (-0.03, 0.04) | 0.78 | -0.03 (-0.06, 0) | 0.09 | 0.02 (-0.04, 0.08) | 0.51 | 0.01 (-0.06, 0.08) | 0.77 | 0.00 (-0.03, 0.04) | 0.78 |
| High Deprivation | 0.08 (-0.88, 1.04) | 0.88 | 0.98 (0.09, 1.88) | 0.03 | 0.03 (0, 0.07) | 0.08 | -0.04 (-0.07, 0) | 0.03 | 0.10 (0.04, 0.16) | 0.00 | 0.06 (-0.01, 0.13) | 0.07 | 0.03 (0, 0.06) | 0.09 |
| **Financial Security** | | | | | | | | | | | | | | |
| Financially Secure | Reference category | | | | | | | | | | | | | |
| Financially Struggling | -0.44 (-1.29, 0.4) | 0.31 | -0.81 (-1.6, -0.02) | 0.04 | 0.01 (-0.02, 0.05) | 0.43 | -0.01 (-0.03, 0.02) | 0.72 | 0.02 (-0.03, 0.08) | 0.45 | 0.02 (-0.04, 0.08) | 0.44 | 0.01 (-0.02, 0.04) | 0.48 |
| **Maternal Education** | | | | | | | | | | | | | | |
| Tertiary Degree (UK or Foreign) | Reference category | | | | | | | | | | | | | |
| A Level/A Level Equivalent/Other (e.g. City and Guilds, RSA/OCR, BTEC, Foreign Other) | -0.71 (-1.86, 0.45) | 0.23 | 0.17 (-0.91, 1.24) | 0.76 | 0.04 (-0.01, 0.08) | 0.11 | 0.01 (-0.03, 0.05) | 0.57 | -0.02 (-0.1, 0.05) | 0.51 | 0.04 (-0.04, 0.12) | 0.35 | 0.00 (-0.04, 0.04) | 0.90 |
| 5 GCSEs or Equivalent | 0.12 (-0.92, 1.16) | 0.83 | 0.00 (-0.97, 0.98) | 0.99 | 0.03 (-0.01, 0.07) | 0.11 | -0.01 (-0.04, 0.03) | 0.74 | 0.00 (-0.06, 0.07) | 0.91 | 0.04 (-0.03, 0.12) | 0.29 | 0.01 (-0.03, 0.04) | 0.69 |
| Less than 5 GCSEs or Equivalent | 0.28 (-0.84, 1.4) | 0.62 | 0.33 (-0.72, 1.38) | 0.54 | 0.01 (-0.03, 0.05) | 0.64 | -0.03 (-0.07, 0) | 0.09 | 0.03 (-0.04, 0.1) | 0.44 | -0.01 (-0.09, 0.07) | 0.81 | -0.02 (-0.06, 0.02) | 0.30 |
| ***Adjusted*** | | | | | | | | | | | | | | |
| **Neighbourhood Deprivation (IMD)** | | | | | | | | | | | | | | |
| Low Deprivation | Reference category | | | | | | | | | | | | | |
| Medium Deprivation | -0.26 (-1.37, 0.84) | 0.64 | 0.09 (-0.94, 1.12) | 0.86 | 0.00 (-0.04, 0.04) | 0.89 | -0.03 (-0.06, 0.01) | 0.11 | 0.02 (-0.04, 0.09) | 0.51 | -0.01 (-0.08, 0.07) | 0.88 | 0.00 (-0.04, 0.03) | 0.87 |
| High Deprivation | 0.07 (-1.05, 1.19) | 0.91 | 0.63 (-0.42, 1.67) | 0.24 | 0.03 (-0.01, 0.07) | 0.18 | -0.04 (-0.07, -0.01) | 0.02 | 0.10 (0.04, 0.17) | 0.00 | 0.05 (-0.02, 0.13) | 0.17 | 0.02 (-0.01, 0.06) | 0.19 |
| **Financial Security** | | | | | | | | | | | | | | |
| Financially Secure | Reference category | | | | | | | | | | | | | |
| Financially Struggling | -0.33 (-1.28, 0.62) | 0.49 | -0.63 (-1.52, 0.25) | 0.16 | 0.00 (-0.03, 0.04) | 0.79 | 0.00 (-0.03, 0.03) | 0.92 | 0.01 (-0.04, 0.07) | 0.64 | 0.01 (-0.06, 0.07) | 0.83 | 0.00 (-0.03, 0.03) | 0.88 |
| **Maternal Education** | | | | | | | | | | | | | | |
| Tertiary Degree (UK or Foreign) | Reference category | | | | | | | | | | | | | |
| A Level/A Level Equivalent/Other (e.g. City and Guilds, RSA/OCR, BTEC, Foreign Other) | -0.77 (-2.05, 0.52) | 0.24 | 0.23 (-0.97, 1.42) | 0.71 | 0.03 (-0.02, 0.08) | 0.22 | 0.01 (-0.02, 0.05) | 0.49 | -0.02 (-0.09, 0.06) | 0.63 | 0.03 (-0.06, 0.12) | 0.51 | 0.00 (-0.04, 0.04) | 0.97 |
| 5 GCSEs or Equivalent | 0.10 (-1.06, 1.26) | 0.86 | 0.07 (-1.02, 1.15) | 0.90 | 0.02 (-0.02, 0.06) | 0.37 | 0.00 (-0.03, 0.03) | 0.96 | -0.01 (-0.07, 0.06) | 0.84 | 0.01 (-0.06, 0.09) | 0.72 | -0.01 (-0.04, 0.03) | 0.78 |
| Less than 5 GCSEs or Equivalent | -0.01 (-1.27, 1.26) | 0.99 | 0.10 (-1.08, 1.28) | 0.86 | -0.01 (-0.05, 0.04) | 0.75 | -0.02 (-0.05, 0.02) | 0.38 | -0.01 (-0.08, 0.07) | 0.83 | -0.04 (-0.13, 0.04) | 0.30 | -0.04 (-0.08, 0) | 0.07 |

#### Table S22. Complete case results, Longitudinal Study of Parents and Children's Child Health CheckPoint

| | Max. cIMT (mm) | | PWV (m/s) | | Systolic Blood Pressure (mmHg) | | Diastolic Blood Pressure (mmHg) | | LDL-C (mmol/L) | | HDL-C (mmol/L) | | Total Triglycerides (mmol/L) | | Remnant Cholesterol (mmol/L) | | Non-HDL-C (mmol/L) | |
|---|---|---|---|---|---|---|---|---|---|---|---|---|---|---|---|---|---|---|
| | Regression Coefficient (95% CI) | P Value | Regression Coefficient (95% CI) | P Value | Regression Coefficient (95% CI) | P Value | Regression Coefficient (95% CI) | P Value | Regression Coefficient (95% CI) | P Value | Regression Coefficient (95% CI) | P Value | Regression Coefficient (95% CI) | P Value | Regression Coefficient (95% CI) | P Value | Regression Coefficient (95% CI) | P Value |
| ***Unadjusted*** | | | | | | | | | | | | | | | | | | |
| **Neighbourhood Disadvantage** | | | | | | | | | | | | | | | | | | |
| High SEIFA | Reference category | | | | | | | | | | | | | | | | | |
| Medium SEIFA | 0.00 (-0.01, 0.01) | 0.93 | 0.09 (0.02, 0.15) | 0.01 | 0.85 (-0.07, 1.77) | 0.07 | -0.24 (-0.9, 0.43) | 0.48 | 0.00 (-0.05, 0.04) | 0.87 | -0.05 (-0.09, -0.01) | 0.01 | 0.14 (0.06, 0.22) | 0.00 | 0.05 (0, 0.1) | 0.04 | 0.05 (-0.04, 0.13) | 0.28 |
| Low SEIFA | 0.00 (-0.01, 0.01) | 0.61 | 0.11 (0.04, 0.17) | 0.00 | 1.63 (0.7, 2.55) | 0.00 | 0.44 (-0.23, 1.11) | 0.20 | -0.02 (-0.07, 0.02) | 0.33 | -0.07 (-0.11, -0.03) | 0.00 | 0.14 (0.06, 0.22) | 0.00 | 0.03 (-0.02, 0.08) | 0.28 | 0.01 (-0.08, 0.09) | 0.89 |
| **Household Income** | | | | | | | | | | | | | | | | | | |
| Above $68,900; population median | Reference category | | | | | | | | | | | | | | | | | |
| Below $68,900; population median | 0.00 (-0.01, 0.01) | 0.96 | 0.03 (-0.03, 0.09) | 0.32 | 0.95 (0.15, 1.76) | 0.02 | 0.47 (-0.11, 1.05) | 0.11 | -0.02 (-0.06, 0.01) | 0.22 | -0.05 (-0.08, -0.01) | 0.00 | 0.07 (0.01, 0.14) | 0.03 | -0.01 (-0.06, 0.03) | 0.58 | -0.04 (-0.11, 0.04) | 0.36 |
| **Maternal Education** | | | | | | | | | | | | | | | | | | |
| Postgraduate Degree | Reference category | | | | | | | | | | | | | | | | | |
| Bachelor's Degree | 0.00 (-0.01, 0.01) | 0.65 | 0.05 (-0.05, 0.15) | 0.35 | 1.43 (0.04, 2.82) | 0.04 | 1.45 (0.45, 2.46) | 0.00 | 0.04 (-0.02, 0.11) | 0.20 | 0.01 (-0.05, 0.07) | 0.69 | 0.06 (-0.05, 0.18) | 0.29 | 0.08 (0, 0.15) | 0.04 | 0.12 (-0.01, 0.25) | 0.07 |
| Trade/Diploma/Certificate/Other | 0.00 (-0.01, 0.01) | 0.97 | 0.06 (-0.04, 0.15) | 0.25 | 2.42 (1.1, 3.74) | 0.00 | 1.59 (0.64, 2.55) | 0.00 | 0.02 (-0.04, 0.09) | 0.46 | -0.02 (-0.08, 0.03) | 0.44 | 0.13 (0.02, 0.25) | 0.02 | 0.07 (0, 0.15) | 0.04 | 0.10 (-0.03, 0.23) | 0.13 |
| Less than Year 12 of high school | 0.00 (-0.01, 0.01) | 0.76 | 0.04 (-0.06, 0.14) | 0.42 | 2.06 (0.65, 3.47) | 0.00 | 1.30 (0.28, 2.32) | 0.01 | 0.01 (-0.06, 0.07) | 0.88 | 0.00 (-0.06, 0.06) | 0.93 | 0.11 (-0.01, 0.23) | 0.08 | 0.04 (-0.04, 0.12) | 0.31 | 0.05 (-0.09, 0.18) | 0.52 |
| ***Adjusted*** | | | | | | | | | | | | | | | | | | |
| **Neighbourhood Disadvantage** | | | | | | | | | | | | | | | | | | |
| High SEIFA | Reference category | | | | | | | | | | | | | | | | | |
| Medium SEIFA | 0.00 (-0.01, 0.01) | 0.91 | 0.08 (0.01, 0.15) | 0.02 | 0.76 (-0.18, 1.7) | 0.11 | -0.27 (-0.95, 0.41) | 0.44 | 0.00 (-0.05, 0.04) | 0.84 | -0.05 (-0.09, -0.01) | 0.02 | 0.14 (0.07, 0.22) | 0.00 | 0.05 (0, 0.1) | 0.07 | 0.04 (-0.05, 0.13) | 0.35 |
| Low SEIFA | 0.00 (-0.01, 0.01) | 0.78 | 0.09 (0.02, 0.15) | 0.01 | 1.36 (0.39, 2.32) | 0.01 | 0.48 (-0.22, 1.17) | 0.18 | -0.01 (-0.06, 0.03) | 0.53 | -0.05 (-0.09, -0.01) | 0.01 | 0.15 (0.07, 0.23) | 0.00 | 0.04 (-0.01, 0.09) | 0.16 | 0.02 (-0.07, 0.11) | 0.63 |
| **Household Income** | | | | | | | | | | | | | | | | | | |
| Above $68,900; population median | Reference category | | | | | | | | | | | | | | | | | |
| Below $68,900; population median | 0.00 (-0.01, 0.01) | 0.75 | 0.02 (-0.03, 0.08) | 0.40 | 0.83 (0.01, 1.65) | 0.05 | 0.34 (-0.25, 0.93) | 0.26 | -0.02 (-0.06, 0.02) | 0.24 | -0.04 (-0.07, -0.01) | 0.02 | 0.07 (0, 0.14) | 0.04 | -0.01 (-0.05, 0.03) | 0.65 | -0.03 (-0.11, 0.04) | 0.40 |
| **Maternal Education** | | | | | | | | | | | | | | | | | | |
| Postgraduate Degree | Reference category | | | | | | | | | | | | | | | | | |
| Bachelor's Degree | 0.00 (-0.01, 0.01) | 0.66 | 0.05 (-0.05, 0.15) | 0.32 | 1.42 (-0.01, 2.85) | 0.05 | 1.49 (0.46, 2.52) | 0.00 | 0.06 (-0.01, 0.12) | 0.10 | 0.01 (-0.05, 0.07) | 0.75 | 0.11 (-0.01, 0.22) | 0.08 | 0.09 (0.02, 0.17) | 0.02 | 0.15 (0.01, 0.29) | 0.03 |
| Trade/Diploma/Certificate/Other | 0.00 (-0.01, 0.01) | 0.69 | 0.03 (-0.06, 0.13) | 0.48 | 2.21 (0.84, 3.58) | 0.00 | 1.63 (0.65, 2.61) | 0.00 | 0.05 (-0.02, 0.11) | 0.16 | -0.02 (-0.08, 0.03) | 0.43 | 0.20 (0.08, 0.31) | 0.00 | 0.11 (0.03, 0.18) | 0.01 | 0.15 (0.02, 0.29) | 0.02 |
| Less than Year 12 of high school | 0.00 (-0.02, 0.01) | 0.49 | 0.03 (-0.07, 0.14) | 0.56 | 1.89 (0.42, 3.36) | 0.01 | 1.40 (0.34, 2.45) | 0.01 | 0.02 (-0.05, 0.1) | 0.51 | 0.00 (-0.06, 0.06) | 1.00 | 0.16 (0.03, 0.28) | 0.01 | 0.06 (-0.02, 0.14) | 0.13 | 0.09 (-0.06, 0.23) | 0.24 |

### Table S23. Complete case results, Northern Finland Birth Cohort 1986

| | Systolic Blood Pressure | | Diastolic Blood Pressure | | LDL-C | | HDL-C | | Total Triglycerides | | Non-HDL-C | | Remnant C | |
|---|---|---|---|---|---|---|---|---|---|---|---|---|---|---|
| | Regression Coefficient (95% CI) | P Value | Regression Coefficient (95% CI) | P Value | Regression Coefficient (95% CI) | P Value | Regression Coefficient (95% CI) | P Value | Regression Coefficient (95% CI) | P Value | Regression Coefficient (95% CI) | P Value | Regression Coefficient (95% CI) | P Value |
| ***Unadjusted*** | | | | | | | | | | | | | | |
| **Neighbourhood Disadvantage** | | | | | | | | | | | | | | |
| Low Disadvantage | Reference category | | | | | | | | | | | | | |
| Medium Disadvantage | 0.08 (-0.68, 0.84) | 0.84 | 0.18 (-0.27, 0.64) | 0.43 | 0.05 (0.02, 0.08) | 0.00 | 0.01 (-0.01, 0.03) | 0.26 | 0.02 (-0.01, 0.05) | 0.24 | 0.08 (0.03, 0.13) | 0.00 | 0.03 (0, 0.05) | 0.03 |
| High Disadvantage | -0.63 (-1.39, 0.12) | 0.10 | 0.24 (-0.21, 0.7) | 0.30 | 0.05 (0.02, 0.08) | 0.00 | 0.01 (-0.01, 0.03) | 0.20 | 0.02 (0, 0.05) | 0.10 | 0.09 (0.04, 0.14) | 0.00 | 0.04 (0.01, 0.06) | 0.01 |
| **Parental Occupational Class** | | | | | | | | | | | | | | |
| High | Reference category | | | | | | | | | | | | | |
| Low | 1.93 (1.05, 2.82) | 0.00 | 0.76 (0.22, 1.29) | 0.01 | 0.06 (0.02, 0.09) | 0.00 | 0.00 (-0.02, 0.03) | 0.87 | 0.02 (-0.02, 0.05) | 0.35 | 0.09 (0.03, 0.15) | 0.00 | 0.04 (0.01, 0.07) | 0.01 |
| **Maternal Education** | | | | | | | | | | | | | | |
| University Degree | Reference category | | | | | | | | | | | | | |
| Matriculation | -1.06 (-2.45, 0.33) | 0.14 | -0.22 (-1.06, 0.62) | 0.60 | 0.03 (-0.02, 0.09) | 0.25 | -0.01 (-0.05, 0.03) | 0.70 | 0.02 (-0.03, 0.07) | 0.52 | 0.06 (-0.04, 0.16) | 0.22 | 0.03 (-0.02, 0.07) | 0.24 |
| Any Vocational School/College | 0.05 (-1.27, 1.36) | 0.94 | 0.33 (-0.46, 1.13) | 0.41 | 0.05 (-0.01, 0.1) | 0.11 | -0.02 (-0.06, 0.01) | 0.19 | 0.03 (-0.02, 0.08) | 0.27 | 0.08 (-0.02, 0.17) | 0.10 | 0.03 (-0.01, 0.08) | 0.13 |
| Less than 10 Years of Schooling | 0.45 (-0.94, 1.83) | 0.53 | 0.58 (-0.26, 1.42) | 0.17 | 0.04 (-0.01, 0.1) | 0.14 | -0.03 (-0.07, 0.01) | 0.09 | 0.08 (0.02, 0.13) | 0.00 | 0.09 (-0.01, 0.19) | 0.07 | 0.05 (0, 0.09) | 0.05 |
| ***Adjusted*** | | | | | | | | | | | | | | |
| **Neighbourhood Disadvantage** | | | | | | | | | | | | | | |
| Low Disadvantage | Reference category | | | | | | | | | | | | | |
| Medium Disadvantage | 0.10 (-0.58, 0.79) | 0.77 | 0.15 (-0.3, 0.61) | 0.51 | 0.05 (0.02, 0.08) | 0.00 | 0.01 (-0.01, 0.03) | 0.19 | 0.02 (-0.01, 0.04) | 0.26 | 0.08 (0.03, 0.13) | 0.00 | 0.03 (0, 0.05) | 0.03 |
| High Disadvantage | -0.66 (-1.35, 0.02) | 0.06 | 0.23 (-0.22, 0.68) | 0.32 | 0.06 (0.03, 0.09) | 0.00 | 0.02 (0, 0.04) | 0.12 | 0.03 (0, 0.05) | 0.06 | 0.10 (0.04, 0.15) | 0.00 | 0.04 (0.02, 0.06) | 0.00 |
| **Parental Occupational Class** | | | | | | | | | | | | | | |
| High | Reference category | | | | | | | | | | | | | |
| Low | 2.08 (1.27, 2.88) | 0.00 | 0.81 (0.28, 1.35) | 0.00 | 0.06 (0.02, 0.09) | 0.00 | 0.00 (-0.02, 0.02) | 0.99 | 0.02 (-0.02, 0.05) | 0.30 | 0.09 (0.03, 0.16) | 0.00 | 0.04 (0.01, 0.07) | 0.01 |
| **Maternal Education** | | | | | | | | | | | | | | |
| University Degree | Reference category | | | | | | | | | | | | | |
| Matriculation | -1.10 (-2.37, 0.16) | 0.09 | -0.27 (-1.1, 0.57) | 0.53 | 0.03 (-0.02, 0.09) | 0.25 | 0.00 (-0.04, 0.03) | 0.80 | 0.02 (-0.04, 0.07) | 0.56 | 0.06 (-0.04, 0.16) | 0.24 | 0.03 (-0.02, 0.07) | 0.28 |
| Any Vocational School/College | -0.15 (-1.34, 1.04) | 0.81 | 0.32 (-0.47, 1.11) | 0.43 | 0.05 (-0.01, 0.1) | 0.08 | -0.02 (-0.05, 0.02) | 0.28 | 0.03 (-0.02, 0.08) | 0.26 | 0.08 (-0.01, 0.18) | 0.08 | 0.03 (-0.01, 0.08) | 0.11 |
| Less than 10 Years of Schooling | 0.10 (-1.17, 1.36) | 0.88 | 0.47 (-0.36, 1.31) | 0.27 | 0.05 (-0.01, 0.1) | 0.12 | -0.03 (-0.07, 0.01) | 0.12 | 0.08 (0.03, 0.13) | 0.00 | 0.09 (0, 0.19) | 0.06 | 0.05 (0, 0.09) | 0.04 |

### Table 24. Complete case results, Avon Longitudinal Study of Parents and Children

| | Max. cIMT (mm) | | PWV (m/s) | | Systolic Blood Pressure (mmHg) | | Diastolic Blood Pressure (mmHg) | | LDL-C (mmol/L) | | HDL-C (mmol/L) | | Total Triglycerides (mmol/L) | | Remnant Cholesterol (mmol/L) | | Non-HDL-C (mmol/L) | |
|---|---|---|---|---|---|---|---|---|---|---|---|---|---|---|---|---|---|---|
| | Regression Coefficient (95% CI) | P Value | Regression Coefficient (95% CI) | P Value | Regression Coefficient (95% CI) | P Value | Regression Coefficient (95% CI) | P Value | Regression Coefficient (95% CI) | P Value | Regression Coefficient (95% CI) | P Value | Regression Coefficient (95% CI) | P Value | Regression Coefficient (95% CI) | P Value | Regression Coefficient (95% CI) | P Value |
| ***Unadjusted*** | | | | | | | | | | | | | | | | | | |
| **Neighbourhood Disadvantage (IMD 2000)** | | | | | | | | | | | | | | | | | | |
| Low Deprivation | Reference category | | | | | | | | | | | | | | | | | |
| Medium Deprivation | 0.00 (-0.01, 0) | 0.66 | 0.03 (-0.03, 0.09) | 0.39 | 1.12 (0.28, 1.96) | 0.01 | 0.41 (-0.09, 0.91) | 0.11 | -0.01 (-0.05, 0.02) | 0.41 | -0.02 (-0.04, 0) | 0.06 | 0.02 (-0.01, 0.05) | 0.22 | 0.00 (-0.03, 0.02) | 0.83 | -0.02 (-0.08, 0.04) | 0.56 |
| High Deprivation | -0.01 (-0.01, 0) | 0.03 | 0.02 (-0.05, 0.09) | 0.55 | 0.36 (-0.57, 1.28) | 0.45 | 0.88 (0.32, 1.43) | 0.00 | -0.01 (-0.05, 0.03) | 0.53 | -0.03 (-0.05, 0) | 0.04 | 0.03 (-0.01, 0.06) | 0.15 | 0.01 (-0.02, 0.04) | 0.48 | 0.00 (-0.07, 0.06) | 0.96 |
| **Financial Difficulties** | | | | | | | | | | | | | | | | | | |
| No Difficulties | Reference category | | | | | | | | | | | | | | | | | |
| Difficulties | 0.00 (-0.01, 0) | 0.11 | 0.02 (-0.02, 0.07) | 0.34 | 0.46 (-0.24, 1.16) | 0.19 | 0.31 (-0.11, 0.73) | 0.15 | -0.01 (-0.04, 0.02) | 0.51 | -0.02 (-0.03, 0) | 0.10 | -0.02 (-0.04, 0.01) | 0.22 | -0.01 (-0.03, 0.02) | 0.56 | -0.02 (-0.06, 0.03) | 0.51 |
| **Maternal Education** | | | | | | | | | | | | | | | | | | |
| University Degree | Reference category | | | | | | | | | | | | | | | | | |
| A Levels | 0.00 (-0.01, 0) | 0.04 | 0.00 (-0.06, 0.07) | 0.89 | 0.96 (0, 1.92) | 0.05 | 0.57 (-0.02, 1.15) | 0.06 | -0.02 (-0.06, 0.02) | 0.37 | -0.01 (-0.04, 0.01) | 0.32 | 0.03 (-0.01, 0.06) | 0.11 | 0.00 (-0.03, 0.02) | 0.76 | -0.02 (-0.09, 0.04) | 0.51 |
| O Levels | 0.00 (-0.01, 0) | 0.04 | 0.04 (-0.02, 0.11) | 0.18 | 0.91 (-0.01, 1.84) | 0.05 | 1.18 (0.62, 1.74) | 0.00 | -0.02 (-0.05, 0.02) | 0.42 | -0.01 (-0.04, 0.01) | 0.30 | 0.04 (0, 0.07) | 0.04 | 0.00 (-0.03, 0.03) | 0.86 | -0.02 (-0.08, 0.05) | 0.58 |
| Vocational/CSE | -0.01 (-0.01, 0) | 0.00 | 0.06 (-0.02, 0.13) | 0.14 | 1.88 (0.82, 2.93) | 0.00 | 1.91 (1.27, 2.55) | 0.00 | 0.00 (-0.04, 0.05) | 0.82 | -0.04 (-0.06, -0.01) | 0.01 | 0.05 (0.01, 0.09) | 0.02 | 0.03 (0, 0.06) | 0.09 | 0.03 (-0.04, 0.11) | 0.37 |
| ***Adjusted*** | | | | | | | | | | | | | | | | | | |
| **Neighbourhood Disadvantage (IMD 2000)** | | | | | | | | | | | | | | | | | | |
| Low Deprivation | Reference category | | | | | | | | | | | | | | | | | |
| Medium Deprivation | 0.00 (-0.01, 0) | 0.47 | 0.01 (-0.05, 0.07) | 0.73 | 0.83 (0.09, 1.58) | 0.03 | 0.43 (-0.07, 0.93) | 0.09 | -0.01 (-0.04, 0.03) | 0.76 | -0.02 (-0.04, 0.01) | 0.15 | 0.02 (-0.01, 0.05) | 0.21 | 0.00 (-0.03, 0.03) | 0.97 | 0.00 (-0.06, 0.05) | 0.87 |
| High Deprivation | 0.00 (-0.01, 0) | 0.05 | 0.05 (-0.01, 0.11) | 0.12 | 0.76 (-0.06, 1.59) | 0.07 | 0.78 (0.23, 1.34) | 0.01 | -0.02 (-0.06, 0.01) | 0.20 | -0.04 (-0.06, -0.01) | 0.00 | 0.02 (-0.01, 0.06) | 0.22 | 0.01 (-0.02, 0.03) | 0.72 | -0.02 (-0.08, 0.04) | 0.57 |
| **Financial Difficulties** | | | | | | | | | | | | | | | | | | |
| No Difficulties | Reference category | | | | | | | | | | | | | | | | | |
| Difficulties | 0.00 (-0.01, 0) | 0.12 | 0.03 (-0.01, 0.08) | 0.14 | 0.74 (0.11, 1.37) | 0.02 | 0.24 (-0.19, 0.66) | 0.27 | -0.01 (-0.04, 0.01) | 0.28 | -0.02 (-0.04, 0) | 0.03 | -0.01 (-0.04, 0.02) | 0.42 | -0.01 (-0.03, 0.01) | 0.50 | -0.02 (-0.07, 0.02) | 0.35 |
| **Maternal Education** | | | | | | | | | | | | | | | | | | |
| University Degree | Reference category | | | | | | | | | | | | | | | | | |
| A Levels | -0.01 (-0.01, 0) | 0.03 | 0.00 (-0.06, 0.07) | 0.90 | 0.95 (0.1, 1.81) | 0.03 | 0.52 (-0.06, 1.1) | 0.08 | -0.02 (-0.05, 0.02) | 0.39 | -0.01 (-0.03, 0.01) | 0.34 | 0.03 (-0.01, 0.06) | 0.10 | 0.00 (-0.03, 0.02) | 0.78 | -0.02 (-0.08, 0.04) | 0.54 |
| O Levels | 0.00 (-0.01, 0) | 0.09 | 0.07 (0.01, 0.13) | 0.03 | 1.33 (0.5, 2.15) | 0.00 | 1.08 (0.52, 1.64) | 0.00 | -0.02 (-0.06, 0.01) | 0.18 | -0.02 (-0.04, 0) | 0.05 | 0.03 (0, 0.07) | 0.06 | 0.00 (-0.03, 0.02) | 0.73 | -0.03 (-0.09, 0.03) | 0.36 |
| Vocational/CSE | -0.01 (-0.01, 0) | 0.00 | 0.08 (0.01, 0.15) | 0.03 | 2.47 (1.52, 3.41) | 0.00 | 1.81 (1.18, 2.45) | 0.00 | 0.00 (-0.04, 0.04) | 0.98 | -0.04 (-0.07, -0.02) | 0.00 | 0.05 (0.01, 0.09) | 0.01 | 0.03 (0, 0.06) | 0.09 | 0.03 (-0.04, 0.1) | 0.44 |

### Table S25. Subgroup results in White-British children, Born in Bradford

| | Systolic Blood Pressure | | Diastolic Blood Pressure | | LDL-C | | HDL-C | | Total Triglycerides | | Non-HDL-C | | Remnant C | |
|---|---|---|---|---|---|---|---|---|---|---|---|---|---|---|
| | Regression Coefficient (95% CI) | P Value | Regression Coefficient (95% CI) | P Value | Regression Coefficient (95% CI) | P Value | Regression Coefficient (95% CI) | P Value | Regression Coefficient (95% CI) | P Value | Regression Coefficient (95% CI) | P Value | Regression Coefficient (95% CI) | P Value |
| ***Unadjusted*** | | | | | | | | | | | | | | |
| **Neighbourhood Deprivation (IMD)** | | | | | | | | | | | | | | |
| Low Deprivation | Reference category | | | | | | | | | | | | | |
| Medium Deprivation | 0.05 (-1.69, 1.79) | 0.96 | 0.10 (-1.51, 1.72) | 0.90 | 0.02 (-0.06, 0.09) | 0.70 | -0.02 (-0.07, 0.03) | 0.38 | 0.03 (-0.11, 0.17) | 0.65 | 0.02 (-0.06, 0.1) | 0.68 | 0.00 (-0.04, 0.05) | 0.93 |
| High Deprivation | 0.16 (-1.61, 1.93) | 0.86 | 0.58 (-1.1, 2.25) | 0.50 | 0.02 (-0.07, 0.1) | 0.72 | -0.03 (-0.09, 0.02) | 0.24 | 0.11 (-0.04, 0.27) | 0.15 | 0.03 (-0.06, 0.12) | 0.51 | 0.01 (-0.03, 0.06) | 0.56 |
| **Financial Security** | | | | | | | | | | | | | | |
| Financially Secure | Reference category | | | | | | | | | | | | | |
| Financially Struggling | -0.45 (-1.98, 1.09) | 0.57 | -0.80 (-2.21, 0.61) | 0.26 | 0.02 (-0.05, 0.09) | 0.59 | 0.01 (-0.04, 0.05) | 0.80 | 0.02 (-0.1, 0.15) | 0.71 | 0.03 (-0.05, 0.1) | 0.52 | 0.01 (-0.04, 0.05) | 0.80 |
| **Maternal Education** | | | | | | | | | | | | | | |
| Tertiary Degree (UK or Foreign) | Reference category | | | | | | | | | | | | | |
| A Level/A Level Equivalent/Other (e.g. City and Guilds, RSA/OCR, BTEC, Foreign Other) | 0.27 (-1.66, 2.2) | 0.79 | 0.66 (-1.13, 2.45) | 0.47 | 0.02 (-0.07, 0.12) | 0.64 | 0.00 (-0.05, 0.05) | 0.98 | 0.00 (-0.15, 0.15) | 0.97 | 0.01 (-0.09, 0.11) | 0.82 | -0.01 (-0.06, 0.04) | 0.68 |
| 5 GCSEs or Equivalent | 0.09 (-1.75, 1.94) | 0.92 | -0.06 (-1.81, 1.68) | 0.94 | 0.01 (-0.07, 0.1) | 0.76 | -0.02 (-0.07, 0.03) | 0.43 | 0.03 (-0.12, 0.18) | 0.66 | 0.00 (-0.09, 0.09) | 0.94 | -0.02 (-0.06, 0.03) | 0.50 |
| Less than 5 GCSEs or Equivalent | -0.54 (-2.73, 1.65) | 0.63 | -1.28 (-3.33, 0.77) | 0.22 | 0.02 (-0.08, 0.13) | 0.67 | -0.04 (-0.1, 0.03) | 0.26 | 0.08 (-0.09, 0.26) | 0.36 | 0.01 (-0.1, 0.12) | 0.84 | -0.01 (-0.07, 0.05) | 0.70 |
| ***Adjusted*** | | | | | | | | | | | | | | |
| **Neighbourhood Deprivation (IMD)** | | | | | | | | | | | | | | |
| Low Deprivation | Reference category | | | | | | | | | | | | | |
| Medium Deprivation | 0.08 (-1.66, 1.81) | 0.93 | 0.11 (-1.5, 1.72) | 0.90 | 0.02 (-0.06, 0.09) | 0.70 | -0.02 (-0.07, 0.03) | 0.37 | 0.03 (-0.11, 0.17) | 0.65 | 0.02 (-0.06, 0.1) | 0.68 | 0.00 (-0.04, 0.05) | 0.94 |
| High Deprivation | 0.26 (-1.51, 2.02) | 0.77 | 0.58 (-1.1, 2.25) | 0.50 | 0.01 (-0.07, 0.1) | 0.73 | -0.03 (-0.09, 0.02) | 0.24 | 0.11 (-0.04, 0.27) | 0.15 | 0.03 (-0.06, 0.12) | 0.52 | 0.01 (-0.03, 0.06) | 0.58 |
| **Financial Security** | | | | | | | | | | | | | | |
| Financially Secure | Reference category | | | | | | | | | | | | | |
| Financially Struggling | -0.36 (-1.89, 1.17) | 0.65 | -0.78 (-2.19, 0.63) | 0.28 | 0.02 (-0.05, 0.09) | 0.59 | 0.00 (-0.04, 0.05) | 0.86 | 0.02 (-0.1, 0.15) | 0.70 | 0.03 (-0.05, 0.1) | 0.51 | 0.01 (-0.04, 0.05) | 0.78 |
| **Maternal Education** | | | | | | | | | | | | | | |
| Tertiary Degree (UK or Foreign) | Reference category | | | | | | | | | | | | | |
| A Level/A Level Equivalent/Other (e.g. City and Guilds, RSA/OCR, BTEC, Foreign Other) | 0.20 (-1.73, 2.13) | 0.84 | 0.69 (-1.11, 2.48) | 0.45 | 0.02 (-0.07, 0.12) | 0.62 | 0.00 (-0.05, 0.05) | 0.97 | 0.01 (-0.14, 0.16) | 0.95 | 0.01 (-0.08, 0.11) | 0.77 | -0.01 (-0.06, 0.04) | 0.72 |
| 5 GCSEs or Equivalent | -0.02 (-1.86, 1.83) | 0.99 | -0.12 (-1.87, 1.63) | 0.90 | 0.01 (-0.07, 0.1) | 0.78 | -0.02 (-0.07, 0.04) | 0.51 | 0.03 (-0.12, 0.18) | 0.69 | -0.01 (-0.1, 0.08) | 0.90 | -0.02 (-0.07, 0.03) | 0.46 |
| Less than 5 GCSEs or Equivalent | -0.76 (-2.95, 1.43) | 0.50 | -1.33 (-3.38, 0.73) | 0.21 | 0.02 (-0.08, 0.13) | 0.66 | -0.03 (-0.1, 0.03) | 0.30 | 0.08 (-0.1, 0.26) | 0.37 | 0.01 (-0.1, 0.12) | 0.85 | -0.01 (-0.07, 0.05) | 0.67 |

### Table S26. Subgroup results in Pakistani children, Born in Bradford

| | Systolic Blood Pressure | | Diastolic Blood Pressure | | LDL-C | | HDL-C | | Total Triglycerides | | Non-HDL-C | | Remnant C | |
|---|---|---|---|---|---|---|---|---|---|---|---|---|---|---|
| | Regression Coefficient (95% CI) | P Value | Regression Coefficient (95% CI) | P Value | Regression Coefficient (95% CI) | P Value | Regression Coefficient (95% CI) | P Value | Regression Coefficient (95% CI) | P Value | Regression Coefficient (95% CI) | P Value | Regression Coefficient (95% CI) | P Value |
| ***Unadjusted*** | | | | | | | | | | | | | | |
| **Neighbourhood Deprivation (IMD)** | | | | | | | | | | | | | | |
| Low Deprivation | Reference category | | | | | | | | | | | | | |
| Medium Deprivation | -0.25 (-1.67, 1.17) | 0.73 | -0.43 (-1.74, 0.88) | 0.52 | -0.02 (-0.08, 0.05) | 0.60 | -0.03 (-0.07, 0.01) | 0.12 | -0.03 (-0.15, 0.09) | 0.64 | -0.03 (-0.09, 0.04) | 0.43 | -0.01 (-0.05, 0.03) | 0.64 |
| High Deprivation | 0.11 (-1.32, 1.54) | 0.88 | 0.24 (-1.09, 1.57) | 0.72 | 0.00 (-0.06, 0.06) | 0.99 | -0.04 (-0.08, 0) | 0.06 | 0.04 (-0.07, 0.16) | 0.48 | 0.00 (-0.07, 0.07) | 0.95 | 0.00 (-0.04, 0.04) | 0.89 |
| **Financial Security** | | | | | | | | | | | | | | |
| Financially Secure | Reference category | | | | | | | | | | | | | |
| Financially Struggling | -0.40 (-1.56, 0.76) | 0.50 | -1.04 (-2.1, 0.02) | 0.05 | 0.00 (-0.06, 0.05) | 0.89 | -0.01 (-0.04, 0.03) | 0.72 | -0.01 (-0.09, 0.08) | 0.91 | 0.00 (-0.06, 0.06) | 0.94 | 0.00 (-0.03, 0.03) | 0.91 |
| **Maternal Education** | | | | | | | | | | | | | | |
| Tertiary Degree (UK or Foreign) | Reference category | | | | | | | | | | | | | |
| A Level/A Level Equivalent/Other (e.g. City and Guilds, RSA/OCR, BTEC, Foreign Other) | -0.95 (-2.68, 0.77) | 0.28 | 0.45 (-1.14, 2.04) | 0.58 | 0.00 (-0.07, 0.08) | 0.91 | 0.00 (-0.04, 0.05) | 0.84 | 0.00 (-0.12, 0.13) | 0.96 | 0.01 (-0.08, 0.1) | 0.84 | 0.00 (-0.04, 0.05) | 0.85 |
| 5 GCSEs or Equivalent | 0.31 (-1.12, 1.75) | 0.67 | 0.27 (-1.07, 1.61) | 0.69 | 0.01 (-0.06, 0.07) | 0.86 | -0.01 (-0.05, 0.04) | 0.70 | 0.00 (-0.12, 0.11) | 0.94 | 0.01 (-0.06, 0.08) | 0.79 | 0.00 (-0.03, 0.04) | 0.84 |
| Less than 5 GCSEs or Equivalent | 1.10 (-0.39, 2.58) | 0.15 | 1.16 (-0.2, 2.52) | 0.09 | -0.01 (-0.08, 0.06) | 0.78 | -0.03 (-0.07, 0.01) | 0.14 | 0.03 (-0.09, 0.14) | 0.61 | -0.02 (-0.1, 0.05) | 0.54 | -0.01 (-0.05, 0.03) | 0.50 |
| ***Adjusted*** | | | | | | | | | | | | | | |
| **Neighbourhood Deprivation (IMD)** | | | | | | | | | | | | | | |
| Low Deprivation | Reference category | | | | | | | | | | | | | |
| Medium Deprivation | -0.32 (-1.73, 1.1) | 0.66 | -0.42 (-1.73, 0.9) | 0.53 | -0.02 (-0.08, 0.05) | 0.60 | -0.03 (-0.07, 0.01) | 0.10 | -0.03 (-0.15, 0.09) | 0.64 | -0.03 (-0.09, 0.04) | 0.44 | -0.01 (-0.05, 0.03) | 0.65 |
| High Deprivation | 0.13 (-1.29, 1.55) | 0.86 | 0.22 (-1.11, 1.55) | 0.75 | 0.00 (-0.06, 0.06) | 0.99 | -0.04 (-0.09, 0) | 0.04 | 0.04 (-0.07, 0.16) | 0.46 | 0.00 (-0.06, 0.07) | 0.92 | 0.00 (-0.04, 0.04) | 0.87 |
| **Financial Security** | | | | | | | | | | | | | | |
| Financially Secure | Reference category | | | | | | | | | | | | | |
| Financially Struggling | -0.33 (-1.49, 0.82) | 0.57 | -1.07 (-2.13, -0.01) | 0.05 | 0.00 (-0.06, 0.05) | 0.90 | -0.01 (-0.04, 0.03) | 0.68 | 0.00 (-0.09, 0.08) | 0.93 | 0.00 (-0.06, 0.06) | 0.95 | 0.00 (-0.03, 0.03) | 0.91 |
| **Maternal Education** | | | | | | | | | | | | | | |
| Tertiary Degree (UK or Foreign) | Reference category | | | | | | | | | | | | | |
| A Level/A Level Equivalent/Other (e.g. City and Guilds, RSA/OCR, BTEC, Foreign Other) | -1.01 (-2.73, 0.71) | 0.25 | 0.48 (-1.11, 2.07) | 0.56 | 0.00 (-0.07, 0.08) | 0.92 | 0.01 (-0.04, 0.06) | 0.82 | 0.00 (-0.12, 0.13) | 0.97 | 0.01 (-0.08, 0.1) | 0.85 | 0.00 (-0.04, 0.05) | 0.86 |
| 5 GCSEs or Equivalent | 0.29 (-1.14, 1.72) | 0.69 | 0.30 (-1.04, 1.64) | 0.66 | 0.01 (-0.06, 0.07) | 0.88 | -0.01 (-0.05, 0.04) | 0.80 | -0.01 (-0.12, 0.11) | 0.92 | 0.01 (-0.06, 0.08) | 0.81 | 0.00 (-0.03, 0.04) | 0.86 |
| Less than 5 GCSEs or Equivalent | 0.94 (-0.55, 2.44) | 0.22 | 1.24 (-0.13, 2.61) | 0.07 | -0.01 (-0.08, 0.06) | 0.76 | -0.03 (-0.07, 0.01) | 0.18 | 0.03 (-0.09, 0.14) | 0.64 | -0.02 (-0.1, 0.05) | 0.51 | -0.01 (-0.05, 0.02) | 0.48 |

### Table S27. Effect modification (interaction test) by ethnicity, Born in Bradford

| | Systolic Blood Pressure | | Diastolic Blood Pressure | | LDL-C | | HDL-C | | Total Triglycerides | | Non-HDL-C | | Remnant C | |
|---|---|---|---|---|---|---|---|---|---|---|---|---|---|---|
| | Regression Coefficient (95% CI) | P Value | Regression Coefficient (95% CI) | P Value | Regression Coefficient (95% CI) | P Value | Regression Coefficient (95% CI) | P Value | Regression Coefficient (95% CI) | P Value | Regression Coefficient (95% CI) | P Value | Regression Coefficient (95% CI) | P Value |
| ***Unadjusted*** | | | | | | | | | | | | | | |
| **Neighbourhood Deprivation (IMD)** | | | | | | | | | | | | | | |
| Low Deprivation | Reference category | | | | | | | | | | | | | |
| Medium Deprivation | -0.36 (-2.64, 1.92) | 0.76 | -0.74 (-2.85, 1.38) | 0.50 | -0.08 (-0.18, 0.01) | 0.07 | -0.04 (-0.11, 0.04) | 0.33 | -0.07 (-0.21, 0.08) | 0.38 | -0.11 (-0.27, 0.06) | 0.21 | -0.02 (-0.1, 0.06) | 0.60 |
| High Deprivation | -0.17 (-2.48, 2.13) | 0.88 | -0.56 (-2.7, 1.58) | 0.61 | -0.04 (-0.14, 0.05) | 0.38 | -0.03 (-0.11, 0.05) | 0.43 | -0.15 (-0.3, 0) | 0.05 | -0.07 (-0.24, 0.1) | 0.41 | -0.03 (-0.11, 0.05) | 0.49 |
| **Financial Security** | | | | | | | | | | | | | | |
| Financially Secure | Reference category | | | | | | | | | | | | | |
| Financially Struggling | 0.00 (-1.95, 1.95) | 1.00 | -0.27 (-2.08, 1.55) | 0.77 | -0.05 (-0.13, 0.02) | 0.18 | -0.04 (-0.1, 0.02) | 0.23 | -0.05 (-0.18, 0.07) | 0.42 | -0.08 (-0.22, 0.07) | 0.29 | -0.02 (-0.09, 0.04) | 0.50 |
| **Maternal Education** | | | | | | | | | | | | | | |
| Tertiary Degree (UK or Foreign) | Reference category | | | | | | | | | | | | | |
| A Level/A Level Equivalent/Other (e.g. City and Guilds, RSA/OCR, BTEC, Foreign Other) | -1.43 (-4.02, 1.17) | 0.28 | -0.11 (-2.53, 2.31) | 0.93 | -0.02 (-0.12, 0.08) | 0.70 | 0.01 (-0.07, 0.1) | 0.77 | 0.05 (-0.12, 0.21) | 0.59 | 0.01 (-0.18, 0.2) | 0.94 | 0.03 (-0.06, 0.12) | 0.55 |
| 5 GCSEs or Equivalent | 0.22 (-2.16, 2.61) | 0.86 | 0.43 (-1.8, 2.65) | 0.71 | 0.02 (-0.08, 0.12) | 0.69 | 0.03 (-0.05, 0.1) | 0.50 | -0.05 (-0.2, 0.1) | 0.52 | 0.07 (-0.1, 0.24) | 0.43 | 0.05 (-0.03, 0.13) | 0.23 |
| Less than 5 GCSEs or Equivalent | 1.71 (-0.98, 4.4) | 0.21 | 2.77 (0.26, 5.28) | 0.03 | -0.06 (-0.17, 0.05) | 0.27 | 0.02 (-0.06, 0.11) | 0.59 | -0.07 (-0.25, 0.11) | 0.44 | -0.06 (-0.26, 0.14) | 0.53 | 0.00 (-0.1, 0.09) | 0.95 |
| ***Adjusted*** | | | | | | | | | | | | | | |
| **Neighbourhood Deprivation (IMD)** | | | | | | | | | | | | | | |
| Low Deprivation | Reference category | | | | | | | | | | | | | |
| Medium Deprivation | -0.76 (-3.29, 1.76) | 0.55 | -0.73 (-3.09, 1.62) | 0.54 | -0.08 (-0.17, 0.02) | 0.10 | -0.04 (-0.11, 0.04) | 0.34 | -0.04 (-0.19, 0.11) | 0.56 | -0.09 (-0.26, 0.08) | 0.31 | -0.01 (-0.09, 0.07) | 0.81 |
| High Deprivation | -0.09 (-2.68, 2.5) | 0.95 | -0.45 (-2.86, 1.96) | 0.71 | -0.03 (-0.13, 0.06) | 0.49 | -0.02 (-0.1, 0.06) | 0.64 | -0.14 (-0.29, 0.01) | 0.08 | -0.05 (-0.23, 0.12) | 0.57 | -0.02 (-0.1, 0.07) | 0.69 |
| **Financial Security** | | | | | | | | | | | | | | |
| Financially Secure | Reference category | | | | | | | | | | | | | |
| Financially Struggling | 0.21 (-1.96, 2.38) | 0.85 | -0.27 (-2.29, 1.76) | 0.80 | -0.06 (-0.14, 0.02) | 0.16 | -0.05 (-0.11, 0.02) | 0.15 | -0.04 (-0.17, 0.09) | 0.59 | -0.08 (-0.23, 0.06) | 0.26 | -0.03 (-0.1, 0.04) | 0.48 |
| **Maternal Education** | | | | | | | | | | | | | | |
| Tertiary Degree (UK or Foreign) | Reference category | | | | | | | | | | | | | |
| A Level/A Level Equivalent/Other (e.g. City and Guilds, RSA/OCR, BTEC, Foreign Other) | -1.17 (-4.02, 1.69) | 0.42 | 0.51 (-2.16, 3.17) | 0.71 | -0.02 (-0.12, 0.09) | 0.75 | 0.00 (-0.08, 0.09) | 0.91 | 0.07 (-0.1, 0.24) | 0.41 | 0.01 (-0.18, 0.21) | 0.90 | 0.03 (-0.06, 0.12) | 0.53 |
| 5 GCSEs or Equivalent | -0.15 (-2.79, 2.48) | 0.91 | 0.59 (-1.87, 3.05) | 0.64 | 0.02 (-0.08, 0.12) | 0.68 | 0.01 (-0.06, 0.09) | 0.75 | -0.03 (-0.19, 0.12) | 0.66 | 0.08 (-0.1, 0.25) | 0.40 | 0.06 (-0.03, 0.14) | 0.19 |
| Less than 5 GCSEs or Equivalent | 1.92 (-1.07, 4.91) | 0.21 | 2.24 (-0.55, 5.02) | 0.12 | -0.05 (-0.17, 0.06) | 0.36 | 0.02 (-0.07, 0.1) | 0.73 | -0.02 (-0.2, 0.16) | 0.84 | -0.05 (-0.25, 0.16) | 0.64 | 0.00 (-0.09, 0.1) | 0.93 |

### Table S28. Subgroup results in girls, Born in Bradford

| | Systolic Blood Pressure | | Diastolic Blood Pressure | | LDL-C | | HDL-C | | Total Triglycerides | | Non-HDL-C | | Remnant C | |
|---|---|---|---|---|---|---|---|---|---|---|---|---|---|---|
| | Regression Coefficient (95% CI) | P Value | Regression Coefficient (95% CI) | P Value | Regression Coefficient (95% CI) | P Value | Regression Coefficient (95% CI) | P Value | Regression Coefficient (95% CI) | P Value | Regression Coefficient (95% CI) | P Value | Regression Coefficient (95% CI) | P Value |
| ***Unadjusted*** | | | | | | | | | | | | | | |
| **Neighbourhood Deprivation (IMD)** | | | | | | | | | | | | | | |
| Low Deprivation | Reference category | | | | | | | | | | | | | |
| Medium Deprivation | -0.57 (-1.98, 0.85) | 0.43 | 0.39 (-0.88, 1.67) | 0.55 | 0.01 (-0.05, 0.08) | 0.66 | -0.02 (-0.06, 0.02) | 0.27 | 0.01 (-0.11, 0.12) | 0.91 | 0.02 (-0.04, 0.09) | 0.51 | 0.01 (-0.03, 0.05) | 0.70 |
| High Deprivation | -0.16 (-1.52, 1.2) | 0.82 | 0.53 (-0.73, 1.78) | 0.41 | 0.02 (-0.04, 0.09) | 0.54 | -0.03 (-0.07, 0.01) | 0.12 | 0.06 (-0.06, 0.17) | 0.33 | 0.03 (-0.04, 0.1) | 0.37 | 0.01 (-0.03, 0.05) | 0.60 |
| **Financial Security** | | | | | | | | | | | | | | |
| Financially Secure | Reference category | | | | | | | | | | | | | |
| Financially Struggling | -1.03 (-2.22, 0.17) | 0.09 | -1.38 (-2.49, -0.27) | 0.01 | 0.01 (-0.05, 0.07) | 0.72 | 0.00 (-0.03, 0.04) | 0.95 | -0.02 (-0.11, 0.08) | 0.75 | 0.01 (-0.05, 0.08) | 0.69 | 0.00 (-0.03, 0.04) | 0.87 |
| **Maternal Education** | | | | | | | | | | | | | | |
| Tertiary Degree (UK or Foreign) | Reference category | | | | | | | | | | | | | |
| A Level/A Level Equivalent/Other (e.g. City and Guilds, RSA/OCR, BTEC, Foreign Other) | -0.27 (-2.02, 1.49) | 0.77 | 0.42 (-1.18, 2.02) | 0.61 | 0.02 (-0.06, 0.1) | 0.56 | 0.01 (-0.04, 0.06) | 0.68 | 0.00 (-0.13, 0.14) | 0.97 | 0.04 (-0.05, 0.12) | 0.40 | 0.01 (-0.03, 0.06) | 0.55 |
| 5 GCSEs or Equivalent | -0.28 (-1.77, 1.2) | 0.71 | -0.70 (-2.08, 0.68) | 0.32 | 0.02 (-0.05, 0.09) | 0.62 | -0.02 (-0.06, 0.03) | 0.43 | 0.01 (-0.11, 0.13) | 0.89 | 0.02 (-0.05, 0.1) | 0.57 | 0.00 (-0.04, 0.04) | 0.86 |
| Less than 5 GCSEs or Equivalent | -0.55 (-2.2, 1.09) | 0.51 | -0.48 (-1.98, 1.02) | 0.53 | 0.01 (-0.07, 0.09) | 0.77 | -0.03 (-0.07, 0.01) | 0.14 | 0.07 (-0.06, 0.2) | 0.30 | 0.02 (-0.06, 0.1) | 0.70 | 0.00 (-0.04, 0.05) | 0.86 |
| ***Adjusted*** | | | | | | | | | | | | | | |
| **Neighbourhood Deprivation (IMD)** | | | | | | | | | | | | | | |
| Low Deprivation | Reference category | | | | | | | | | | | | | |
| Medium Deprivation | -0.46 (-1.94, 1.01) | 0.54 | 0.06 (-1.27, 1.39) | 0.93 | 0.01 (-0.05, 0.08) | 0.68 | -0.02 (-0.06, 0.02) | 0.34 | 0.01 (-0.1, 0.12) | 0.88 | 0.02 (-0.04, 0.09) | 0.55 | 0.01 (-0.03, 0.05) | 0.72 |
| High Deprivation | 0.10 (-1.31, 1.51) | 0.89 | 0.17 (-1.13, 1.47) | 0.80 | 0.02 (-0.05, 0.09) | 0.55 | -0.03 (-0.08, 0.01) | 0.13 | 0.06 (-0.05, 0.18) | 0.29 | 0.03 (-0.04, 0.1) | 0.38 | 0.01 (-0.03, 0.05) | 0.59 |
| **Financial Security** | | | | | | | | | | | | | | |
| Financially Secure | Reference category | | | | | | | | | | | | | |
| Financially Struggling | -0.95 (-2.15, 0.24) | 0.12 | -1.43 (-2.54, -0.32) | 0.01 | 0.01 (-0.05, 0.07) | 0.73 | 0.00 (-0.03, 0.03) | 0.98 | -0.01 (-0.11, 0.08) | 0.77 | 0.01 (-0.05, 0.07) | 0.69 | 0.00 (-0.03, 0.04) | 0.87 |
| **Maternal Education** | | | | | | | | | | | | | | |
| Tertiary Degree (UK or Foreign) | Reference category | | | | | | | | | | | | | |
| A Level/A Level Equivalent/Other (e.g. City and Guilds, RSA/OCR, BTEC, Foreign Other) | -0.36 (-2.11, 1.38) | 0.68 | 0.50 (-1.1, 2.1) | 0.54 | 0.02 (-0.06, 0.1) | 0.56 | 0.01 (-0.04, 0.06) | 0.66 | 0.00 (-0.13, 0.13) | 0.98 | 0.04 (-0.05, 0.12) | 0.41 | 0.01 (-0.03, 0.06) | 0.56 |
| 5 GCSEs or Equivalent | -0.42 (-1.91, 1.07) | 0.58 | -0.73 (-2.11, 0.65) | 0.30 | 0.02 (-0.06, 0.09) | 0.64 | -0.01 (-0.06, 0.03) | 0.52 | 0.00 (-0.12, 0.12) | 0.97 | 0.02 (-0.06, 0.09) | 0.61 | 0.00 (-0.04, 0.04) | 0.92 |
| Less than 5 GCSEs or Equivalent | -0.72 (-2.38, 0.94) | 0.39 | -0.61 (-2.13, 0.9) | 0.43 | 0.01 (-0.07, 0.09) | 0.80 | -0.03 (-0.07, 0.01) | 0.22 | 0.07 (-0.07, 0.2) | 0.34 | 0.01 (-0.07, 0.09) | 0.76 | 0.00 (-0.04, 0.05) | 0.92 |

### Table S29. Subgroup results in boys, Born in Bradford

| | Systolic Blood Pressure | | Diastolic Blood Pressure | | LDL-C | | HDL-C | | Total Triglycerides | | Non-HDL-C | | Remnant C | |
|---|---|---|---|---|---|---|---|---|---|---|---|---|---|---|
| | Regression Coefficient (95% CI) | P Value | Regression Coefficient (95% CI) | P Value | Regression Coefficient (95% CI) | P Value | Regression Coefficient (95% CI) | P Value | Regression Coefficient (95% CI) | P Value | Regression Coefficient (95% CI) | P Value | Regression Coefficient (95% CI) | P Value |
| ***Unadjusted*** | | | | | | | | | | | | | | |
| **Neighbourhood Deprivation (IMD)** | | | | | | | | | | | | | | |
| Low Deprivation | Reference category | | | | | | | | | | | | | |
| Medium Deprivation | -0.30 (-1.66, 1.06) | 0.67 | -0.22 (-1.46, 1.03) | 0.74 | -0.01 (-0.07, 0.05) | 0.73 | -0.03 (-0.07, 0.01) | 0.16 | 0.01 (-0.1, 0.11) | 0.87 | -0.01 (-0.08, 0.05) | 0.64 | 0.00 (-0.04, 0.03) | 0.81 |
| High Deprivation | 0.35 (-0.98, 1.69) | 0.60 | 1.48 (0.2, 2.76) | 0.02 | 0.01 (-0.05, 0.07) | 0.78 | -0.03 (-0.07, 0.01) | 0.16 | 0.07 (-0.03, 0.18) | 0.17 | 0.02 (-0.04, 0.08) | 0.53 | 0.01 (-0.02, 0.05) | 0.52 |
| **Financial Security** | | | | | | | | | | | | | | |
| Financially Secure | Reference category | | | | | | | | | | | | | |
| Financially Struggling | 0.18 (-1.02, 1.38) | 0.77 | -0.25 (-1.37, 0.87) | 0.66 | 0.00 (-0.06, 0.06) | 0.98 | 0.00 (-0.04, 0.03) | 0.87 | 0.03 (-0.06, 0.12) | 0.55 | 0.01 (-0.05, 0.07) | 0.83 | 0.01 (-0.02, 0.04) | 0.70 |
| **Maternal Education** | | | | | | | | | | | | | | |
| Tertiary Degree (UK or Foreign) | Reference category | | | | | | | | | | | | | |
| A Level/A Level Equivalent/Other (e.g. City and Guilds, RSA/OCR, BTEC, Foreign Other) | -1.18 (-2.76, 0.4) | 0.14 | -0.06 (-1.58, 1.46) | 0.94 | 0.01 (-0.06, 0.08) | 0.85 | 0.00 (-0.05, 0.05) | 0.93 | 0.00 (-0.12, 0.12) | 0.99 | 0.00 (-0.08, 0.07) | 0.91 | -0.01 (-0.05, 0.03) | 0.60 |
| 5 GCSEs or Equivalent | 0.53 (-0.97, 2.02) | 0.49 | 0.67 (-0.73, 2.06) | 0.35 | 0.01 (-0.06, 0.07) | 0.85 | 0.00 (-0.05, 0.05) | 0.92 | 0.03 (-0.09, 0.15) | 0.64 | 0.01 (-0.06, 0.07) | 0.83 | 0.00 (-0.04, 0.04) | 0.96 |
| Less than 5 GCSEs or Equivalent | 0.98 (-0.62, 2.58) | 0.23 | 1.06 (-0.42, 2.54) | 0.16 | 0.00 (-0.07, 0.06) | 0.91 | -0.04 (-0.08, 0.01) | 0.09 | 0.05 (-0.07, 0.16) | 0.45 | -0.02 (-0.09, 0.05) | 0.59 | -0.02 (-0.06, 0.02) | 0.43 |
| ***Adjusted*** | | | | | | | | | | | | | | |
| **Neighbourhood Deprivation (IMD)** | | | | | | | | | | | | | | |
| Low Deprivation | Reference category | | | | | | | | | | | | | |
| Medium Deprivation | -0.38 (-1.79, 1.03) | 0.60 | -0.75 (-2.04, 0.55) | 0.26 | -0.02 (-0.08, 0.04) | 0.56 | -0.03 (-0.07, 0.01) | 0.14 | 0.00 (-0.11, 0.11) | 0.95 | -0.03 (-0.1, 0.04) | 0.37 | -0.01 (-0.05, 0.02) | 0.51 |
| High Deprivation | 0.29 (-1.09, 1.68) | 0.68 | 0.92 (-0.4, 2.23) | 0.17 | 0.00 (-0.06, 0.06) | 0.98 | -0.03 (-0.08, 0.01) | 0.13 | 0.06 (-0.05, 0.18) | 0.26 | 0.00 (-0.06, 0.07) | 0.89 | 0.00 (-0.03, 0.04) | 0.82 |
| **Financial Security** | | | | | | | | | | | | | | |
| Financially Secure | Reference category | | | | | | | | | | | | | |
| Financially Struggling | 0.24 (-0.96, 1.44) | 0.70 | -0.26 (-1.38, 0.85) | 0.64 | 0.00 (-0.06, 0.06) | 0.97 | 0.00 (-0.04, 0.03) | 0.83 | 0.03 (-0.06, 0.12) | 0.55 | 0.01 (-0.05, 0.07) | 0.82 | 0.01 (-0.02, 0.04) | 0.69 |
| **Maternal Education** | | | | | | | | | | | | | | |
| Tertiary Degree (UK or Foreign) | Reference category | | | | | | | | | | | | | |
| A Level/A Level Equivalent/Other (e.g. City and Guilds, RSA/OCR, BTEC, Foreign Other) | -1.17 (-2.75, 0.41) | 0.15 | 0.15 (-1.38, 1.68) | 0.85 | 0.01 (-0.06, 0.08) | 0.80 | 0.00 (-0.05, 0.05) | 0.99 | 0.00 (-0.12, 0.12) | 0.98 | 0.00 (-0.07, 0.07) | 1.00 | -0.01 (-0.05, 0.03) | 0.68 |
| 5 GCSEs or Equivalent | 0.53 (-0.97, 2.04) | 0.49 | 0.66 (-0.74, 2.06) | 0.35 | 0.01 (-0.06, 0.07) | 0.88 | 0.00 (-0.05, 0.05) | 0.98 | 0.02 (-0.1, 0.15) | 0.70 | 0.01 (-0.06, 0.07) | 0.88 | 0.00 (-0.04, 0.04) | 1.00 |
| Less than 5 GCSEs or Equivalent | 0.94 (-0.67, 2.56) | 0.25 | 0.93 (-0.56, 2.42) | 0.22 | -0.01 (-0.08, 0.06) | 0.82 | -0.03 (-0.07, 0.01) | 0.17 | 0.03 (-0.09, 0.15) | 0.60 | -0.03 (-0.1, 0.04) | 0.44 | -0.02 (-0.06, 0.02) | 0.32 |

### Table S30. Effect modification (interaction test) by sex, Born in Bradford

| | Systolic Blood Pressure | | Diastolic Blood Pressure | | LDL-C | | HDL-C | | Total Triglycerides | | Non-HDL-C | | Remnant C | |
|---|---|---|---|---|---|---|---|---|---|---|---|---|---|---|
| | Regression Coefficient (95% CI) | P Value | Regression Coefficient (95% CI) | P Value | Regression Coefficient (95% CI) | P Value | Regression Coefficient (95% CI) | P Value | Regression Coefficient (95% CI) | P Value | Regression Coefficient (95% CI) | P Value | Regression Coefficient (95% CI) | P Value |
| ***Unadjusted*** | | | | | | | | | | | | | | |
| **Neighbourhood Deprivation (IMD)** | | | | | | | | | | | | | | |
| Low Deprivation | Reference category | | | | | | | | | | | | | |
| Medium Deprivation | 0.27 (-1.7, 2.24) | 0.79 | -0.61 (-2.41, 1.2) | 0.51 | -0.02 (-0.11, 0.06) | 0.58 | -0.01 (-0.06, 0.04) | 0.73 | 0.00 (-0.14, 0.15) | 0.97 | -0.04 (-0.13, 0.06) | 0.44 | -0.01 (-0.06, 0.04) | 0.64 |
| High Deprivation | 0.51 (-1.39, 2.42) | 0.60 | 0.95 (-0.82, 2.73) | 0.29 | -0.01 (-0.1, 0.08) | 0.80 | 0.00 (-0.05, 0.06) | 0.95 | 0.02 (-0.14, 0.18) | 0.83 | -0.01 (-0.1, 0.08) | 0.82 | 0.00 (-0.05, 0.05) | 0.96 |
| **Financial Security** | | | | | | | | | | | | | | |
| Financially Secure | Reference category | | | | | | | | | | | | | |
| Financially Struggling | 1.21 (-0.48, 2.9) | 0.16 | 1.13 (-0.46, 2.72) | 0.16 | -0.01 (-0.09, 0.07) | 0.82 | 0.00 (-0.05, 0.04) | 0.86 | 0.04 (-0.09, 0.18) | 0.52 | -0.01 (-0.09, 0.08) | 0.89 | 0.00 (-0.04, 0.05) | 0.90 |
| **Maternal Education** | | | | | | | | | | | | | | |
| Tertiary Degree (UK or Foreign) | Reference category | | | | | | | | | | | | | |
| A Level/A Level Equivalent/Other (e.g. City and Guilds, RSA/OCR, BTEC, Foreign Other) | -0.92 (-3.29, 1.45) | 0.45 | -0.48 (-2.7, 1.75) | 0.67 | -0.02 (-0.12, 0.09) | 0.75 | -0.01 (-0.07, 0.05) | 0.68 | 0.00 (-0.19, 0.18) | 0.98 | -0.04 (-0.15, 0.07) | 0.47 | -0.02 (-0.08, 0.04) | 0.42 |
| 5 GCSEs or Equivalent | 0.81 (-1.31, 2.92) | 0.45 | 1.37 (-0.6, 3.33) | 0.17 | -0.01 (-0.11, 0.09) | 0.81 | 0.01 (-0.04, 0.07) | 0.61 | 0.02 (-0.16, 0.2) | 0.82 | -0.01 (-0.11, 0.09) | 0.78 | 0.00 (-0.06, 0.05) | 0.94 |
| Less than 5 GCSEs or Equivalent | 1.54 (-0.77, 3.84) | 0.19 | 1.54 (-0.57, 3.64) | 0.15 | -0.02 (-0.12, 0.09) | 0.76 | -0.01 (-0.06, 0.05) | 0.81 | -0.02 (-0.2, 0.15) | 0.78 | -0.04 (-0.14, 0.07) | 0.52 | -0.02 (-0.08, 0.04) | 0.52 |
| ***Adjusted*** | | | | | | | | | | | | | | |
| **Neighbourhood Deprivation (IMD)** | | | | | | | | | | | | | | |
| Low Deprivation | Reference category | | | | | | | | | | | | | |
| Medium Deprivation | 0.25 (-1.71, 2.22) | 0.80 | -0.65 (-2.45, 1.15) | 0.48 | -0.02 (-0.11, 0.06) | 0.58 | -0.01 (-0.06, 0.04) | 0.74 | 0.00 (-0.14, 0.15) | 0.98 | -0.04 (-0.13, 0.06) | 0.43 | -0.01 (-0.06, 0.04) | 0.63 |
| High Deprivation | 0.45 (-1.46, 2.35) | 0.64 | 0.92 (-0.85, 2.7) | 0.31 | -0.01 (-0.11, 0.08) | 0.79 | 0.00 (-0.05, 0.06) | 0.88 | 0.01 (-0.15, 0.18) | 0.86 | -0.01 (-0.11, 0.08) | 0.80 | 0.00 (-0.05, 0.05) | 0.98 |
| **Financial Security** | | | | | | | | | | | | | | |
| Financially Secure | Reference category | | | | | | | | | | | | | |
| Financially Struggling | 1.22 (-0.46, 2.9) | 0.16 | 1.16 (-0.43, 2.75) | 0.15 | -0.01 (-0.09, 0.07) | 0.82 | 0.00 (-0.05, 0.04) | 0.88 | 0.04 (-0.09, 0.17) | 0.53 | -0.01 (-0.09, 0.08) | 0.89 | 0.00 (-0.04, 0.05) | 0.89 |
| **Maternal Education** | | | | | | | | | | | | | | |
| Tertiary Degree (UK or Foreign) | Reference category | | | | | | | | | | | | | |
| A Level/A Level Equivalent/Other (e.g. City and Guilds, RSA/OCR, BTEC, Foreign Other) | -0.94 (-3.3, 1.41) | 0.43 | -0.43 (-2.66, 1.79) | 0.70 | -0.02 (-0.12, 0.09) | 0.75 | -0.01 (-0.07, 0.05) | 0.68 | 0.00 (-0.18, 0.18) | 1.00 | -0.04 (-0.15, 0.07) | 0.48 | -0.02 (-0.08, 0.04) | 0.43 |
| 5 GCSEs or Equivalent | 0.79 (-1.32, 2.9) | 0.46 | 1.31 (-0.65, 3.28) | 0.19 | -0.01 (-0.11, 0.09) | 0.80 | 0.01 (-0.04, 0.07) | 0.64 | 0.02 (-0.15, 0.2) | 0.80 | -0.01 (-0.12, 0.09) | 0.77 | 0.00 (-0.06, 0.05) | 0.94 |
| Less than 5 GCSEs or Equivalent | 1.48 (-0.81, 3.78) | 0.20 | 1.51 (-0.59, 3.61) | 0.16 | -0.02 (-0.12, 0.09) | 0.76 | -0.01 (-0.06, 0.05) | 0.85 | -0.03 (-0.2, 0.15) | 0.77 | -0.04 (-0.15, 0.07) | 0.50 | -0.02 (-0.09, 0.04) | 0.51 |

### Table S31. Subgroup results in girls, Longitudinal Study of Parents and Children's Child Health CheckPoint

| | **Max. cIMT (mm)** | | **PWV (m/s)** | | **Systolic Blood Pressure (mmHg)** | | **Diastolic Blood Pressure (mmHg)** | | **LDL-C (mmol/L)** | | **HDL-C (mmol/L)** | | **Total Triglycerides (mmol/L)** | | **Remnant Cholesterol (mmol/L)** | | **Non-HDL-C (mmol/L)** | |
|---|---|---|---|---|---|---|---|---|---|---|---|---|---|---|---|---|---|---|
| | Regression Coefficient (95% CI) | P Value | Regression Coefficient (95% CI) | P Value | Regression Coefficient (95% CI) | P Value | Regression Coefficient (95% CI) | P Value | Regression Coefficient (95% CI) | P Value | Regression Coefficient (95% CI) | P Value | Regression Coefficient (95% CI) | P Value | Regression Coefficient (95% CI) | P Value | Regression Coefficient (95% CI) | P Value |
| ***Unadjusted*** | | | | | | | | | | | | | | | | | | |
| **Neighbourhood Disadvantage** | | | | | | | | | | | | | | | | | | |
| High SEIFA | Reference category | | | | | | | | | | | | | | | | | |
| Medium SEIFA | 0.00 (-0.01, 0.01) | 0.99 | 0.08 (-0.01, 0.16) | 0.07 | 0.56 (-0.71, 1.82) | 0.39 | -0.24 (-1.17, 0.68) | 0.61 | 0.01 (-0.07, 0.09) | 0.84 | -0.05 (-0.1, 0.01) | 0.08 | 0.12 (-0.02, 0.26) | 0.10 | 0.02 (-0.05, 0.08) | 0.62 | 0.02 (-0.09, 0.14) | 0.67 |
| Low SEIFA | 0.01 (0, 0.01) | 0.21 | 0.13 (0.04, 0.22) | 0.00 | 1.55 (0.25, 2.86) | 0.02 | 0.28 (-0.67, 1.22) | 0.57 | -0.01 (-0.09, 0.08) | 0.89 | -0.05 (-0.1, 0) | 0.05 | 0.15 (0, 0.29) | 0.05 | 0.01 (-0.05, 0.07) | 0.75 | 0.00 (-0.11, 0.12) | 0.94 |
| **Household Income** | | | | | | | | | | | | | | | | | | |
| Above $68,900; population median | Reference category | | | | | | | | | | | | | | | | | |
| Below $68,900; population median | 0.00 (-0.01, 0.01) | 0.72 | 0.03 (-0.04, 0.1) | 0.44 | 1.45 (0.35, 2.54) | 0.01 | 0.72 (-0.07, 1.52) | 0.07 | 0.00 (-0.07, 0.07) | 0.94 | -0.03 (-0.08, 0.01) | 0.17 | 0.07 (-0.04, 0.19) | 0.22 | -0.01 (-0.07, 0.05) | 0.74 | -0.01 (-0.11, 0.09) | 0.81 |
| **Maternal Education** | | | | | | | | | | | | | | | | | | |
| Postgraduate Degree | Reference category | | | | | | | | | | | | | | | | | |
| Bachelor's Degree | 0.00 (-0.01, 0.01) | 0.65 | 0.08 (-0.06, 0.21) | 0.26 | 1.00 (-0.97, 2.96) | 0.32 | 1.51 (0.07, 2.95) | 0.04 | 0.03 (-0.1, 0.16) | 0.68 | 0.04 (-0.04, 0.12) | 0.31 | 0.01 (-0.21, 0.24) | 0.90 | 0.04 (-0.07, 0.14) | 0.50 | 0.06 (-0.12, 0.24) | 0.49 |
| Trade/Diploma/Certificate/Other | 0.01 (-0.01, 0.02) | 0.33 | 0.09 (-0.04, 0.22) | 0.16 | 2.16 (0.26, 4.05) | 0.03 | 1.31 (-0.07, 2.69) | 0.06 | 0.02 (-0.11, 0.14) | 0.79 | 0.01 (-0.07, 0.09) | 0.78 | 0.10 (-0.12, 0.32) | 0.39 | 0.03 (-0.07, 0.13) | 0.51 | 0.05 (-0.12, 0.22) | 0.57 |
| Less than Year 12 of high school | 0.00 (-0.01, 0.02) | 0.60 | 0.06 (-0.08, 0.19) | 0.43 | 1.17 (-0.84, 3.18) | 0.26 | 0.77 (-0.69, 2.24) | 0.30 | -0.01 (-0.14, 0.12) | 0.91 | 0.01 (-0.07, 0.1) | 0.75 | 0.11 (-0.13, 0.35) | 0.35 | -0.01 (-0.12, 0.1) | 0.91 | -0.01 (-0.2, 0.17) | 0.89 |
| ***Adjusted*** | | | | | | | | | | | | | | | | | | |
| **Neighbourhood Disadvantage** | | | | | | | | | | | | | | | | | | |
| High SEIFA | Reference category | | | | | | | | | | | | | | | | | |
| Medium SEIFA | 0.00 (-0.01, 0.01) | 0.94 | 0.07 (-0.01, 0.16) | 0.11 | 0.53 (-0.74, 1.8) | 0.41 | -0.16 (-1.08, 0.76) | 0.73 | 0.01 (-0.07, 0.09) | 0.86 | -0.04 (-0.1, 0.01) | 0.11 | 0.12 (-0.02, 0.26) | 0.10 | 0.02 (-0.04, 0.08) | 0.57 | 0.03 (-0.09, 0.14) | 0.66 |
| Low SEIFA | 0.01 (0, 0.01) | 0.19 | 0.12 (0.03, 0.2) | 0.01 | 1.51 (0.2, 2.82) | 0.02 | 0.39 (-0.55, 1.34) | 0.42 | -0.01 (-0.09, 0.08) | 0.89 | -0.05 (-0.1, 0) | 0.08 | 0.15 (0, 0.29) | 0.05 | 0.01 (-0.05, 0.08) | 0.67 | 0.01 (-0.11, 0.12) | 0.89 |
| **Household Income** | | | | | | | | | | | | | | | | | | |
| Above $68,900; population median | Reference category | | | | | | | | | | | | | | | | | |
| Below $68,900; population median | 0.00 (-0.01, 0.01) | 0.71 | 0.03 (-0.05, 0.1) | 0.50 | 1.43 (0.34, 2.53) | 0.01 | 0.76 (-0.04, 1.55) | 0.06 | 0.00 (-0.07, 0.07) | 0.94 | -0.03 (-0.07, 0.01) | 0.19 | 0.07 (-0.05, 0.19) | 0.22 | -0.01 (-0.06, 0.05) | 0.77 | -0.01 (-0.11, 0.09) | 0.83 |
| **Maternal Education** | | | | | | | | | | | | | | | | | | |
| Postgraduate Degree | Reference category | | | | | | | | | | | | | | | | | |
| Bachelor's Degree | 0.00 (-0.01, 0.01) | 0.65 | 0.06 (-0.07, 0.19) | 0.40 | 0.94 (-1.03, 2.91) | 0.35 | 1.66 (0.23, 3.1) | 0.02 | 0.03 (-0.1, 0.16) | 0.67 | 0.05 (-0.03, 0.13) | 0.23 | 0.01 (-0.21, 0.24) | 0.91 | 0.04 (-0.06, 0.14) | 0.43 | 0.07 (-0.11, 0.25) | 0.45 |
| Trade/Diploma/Certificate/Other | 0.01 (-0.01, 0.02) | 0.36 | 0.08 (-0.05, 0.2) | 0.25 | 2.10 (0.2, 4.01) | 0.03 | 1.46 (0.08, 2.83) | 0.04 | 0.02 (-0.11, 0.15) | 0.75 | 0.02 (-0.06, 0.1) | 0.68 | 0.09 (-0.13, 0.32) | 0.40 | 0.04 (-0.06, 0.14) | 0.43 | 0.06 (-0.11, 0.23) | 0.50 |
| Less than Year 12 of high school | 0.00 (-0.01, 0.02) | 0.60 | 0.03 (-0.11, 0.16) | 0.71 | 1.08 (-0.94, 3.1) | 0.30 | 1.01 (-0.45, 2.48) | 0.17 | -0.01 (-0.14, 0.12) | 0.93 | 0.02 (-0.06, 0.11) | 0.56 | 0.11 (-0.13, 0.35) | 0.37 | 0.00 (-0.11, 0.11) | 0.97 | 0.00 (-0.19, 0.18) | 0.97 |

#### Table S32. Subgroup results in boys, Longitudinal Study of Parents and Children's Child Health CheckPoint

| | Max. cIMT (mm) | | PWV (m/s) | | Systolic Blood Pressure (mmHg) | | Diastolic Blood Pressure (mmHg) | | LDL-C (mmol/L) | | HDL-C (mmol/L) | | Total Triglycerides (mmol/L) | | Remnant Cholesterol (mmol/L) | | Non-HDL-C (mmol/L) | |
|---|---|---|---|---|---|---|---|---|---|---|---|---|---|---|---|---|---|---|
| | Regression Coefficient (95% CI) | P Value | Regression Coefficient (95% CI) | P Value | Regression Coefficient (95% CI) | P Value | Regression Coefficient (95% CI) | P Value | Regression Coefficient (95% CI) | P Value | Regression Coefficient (95% CI) | P Value | Regression Coefficient (95% CI) | P Value | Regression Coefficient (95% CI) | P Value | Regression Coefficient (95% CI) | P Value |
| ***Unadjusted*** | | | | | | | | | | | | | | | | | | |
| **Neighbourhood Disadvantage** | | | | | | | | | | | | | | | | | | |
| High SEIFA | Reference category | | | | | | | | | | | | | | | | | |
| Medium SEIFA | 0.00 (-0.01, 0.01) | 0.77 | 0.09 (0, 0.19) | 0.06 | 1.02 (-0.28, 2.33) | 0.12 | -0.29 (-1.23, 0.66) | 0.55 | -0.01 (-0.09, 0.07) | 0.85 | -0.05 (-0.1, 0.01) | 0.11 | 0.13 (-0.02, 0.28) | 0.08 | 0.05 (-0.02, 0.11) | 0.15 | 0.04 (-0.07, 0.15) | 0.50 |
| Low SEIFA | 0.01 (0, 0.02) | 0.05 | 0.09 (0, 0.19) | 0.06 | 1.64 (0.34, 2.93) | 0.01 | 0.61 (-0.34, 1.55) | 0.21 | -0.02 (-0.1, 0.06) | 0.63 | -0.08 (-0.13, -0.03) | 0.00 | 0.15 (0.01, 0.29) | 0.04 | 0.03 (-0.03, 0.1) | 0.33 | 0.01 (-0.1, 0.13) | 0.84 |
| **Household Income** | | | | | | | | | | | | | | | | | | |
| Above $68,900; population median | Reference category | | | | | | | | | | | | | | | | | |
| Below $68,900; population median | 0.01 (0, 0.01) | 0.16 | 0.03 (-0.05, 0.12) | 0.45 | 0.44 (-0.69, 1.57) | 0.45 | 0.22 (-0.58, 1.02) | 0.59 | -0.02 (-0.09, 0.05) | 0.60 | -0.06 (-0.1, -0.01) | 0.02 | 0.11 (-0.01, 0.23) | 0.09 | 0.00 (-0.06, 0.05) | 0.95 | -0.02 (-0.12, 0.08) | 0.67 |
| **Maternal Education** | | | | | | | | | | | | | | | | | | |
| Postgraduate Degree | Reference category | | | | | | | | | | | | | | | | | |
| Bachelor's Degree | 0.00 (-0.01, 0.02) | 0.52 | 0.03 (-0.11, 0.18) | 0.68 | 1.73 (-0.22, 3.68) | 0.08 | 1.26 (-0.13, 2.65) | 0.07 | 0.04 (-0.09, 0.16) | 0.59 | 0.02 (-0.06, 0.1) | 0.60 | 0.00 (-0.23, 0.23) | 1.00 | 0.05 (-0.04, 0.14) | 0.28 | 0.09 (-0.08, 0.25) | 0.30 |
| Trade/Diploma/Certificate/Other | 0.00 (-0.01, 0.01) | 0.57 | 0.03 (-0.11, 0.16) | 0.70 | 2.59 (0.77, 4.41) | 0.00 | 1.79 (0.48, 3.1) | 0.01 | 0.02 (-0.1, 0.14) | 0.78 | -0.02 (-0.1, 0.05) | 0.57 | 0.09 (-0.12, 0.3) | 0.39 | 0.05 (-0.04, 0.14) | 0.26 | 0.07 (-0.09, 0.22) | 0.40 |
| Less than Year 12 of high school | 0.01 (-0.01, 0.02) | 0.41 | 0.04 (-0.11, 0.18) | 0.63 | 2.77 (0.79, 4.75) | 0.01 | 1.73 (0.29, 3.17) | 0.02 | 0.02 (-0.11, 0.15) | 0.76 | 0.01 (-0.08, 0.09) | 0.87 | 0.09 (-0.14, 0.32) | 0.44 | 0.05 (-0.04, 0.14) | 0.30 | 0.07 (-0.1, 0.24) | 0.43 |
| ***Adjusted*** | | | | | | | | | | | | | | | | | | |
| **Neighbourhood Disadvantage** | | | | | | | | | | | | | | | | | | |
| High SEIFA | Reference category | | | | | | | | | | | | | | | | | |
| Medium SEIFA | 0.00 (-0.01, 0.01) | 0.92 | 0.08 (-0.02, 0.17) | 0.12 | 0.90 (-0.41, 2.21) | 0.18 | -0.23 (-1.18, 0.72) | 0.64 | -0.01 (-0.09, 0.08) | 0.86 | -0.04 (-0.09, 0.02) | 0.18 | 0.13 (-0.02, 0.28) | 0.09 | 0.05 (-0.02, 0.11) | 0.14 | 0.04 (-0.07, 0.15) | 0.47 |
| Low SEIFA | 0.01 (0, 0.02) | 0.03 | 0.08 (-0.02, 0.17) | 0.12 | 1.52 (0.22, 2.82) | 0.02 | 0.66 (-0.29, 1.61) | 0.17 | -0.02 (-0.1, 0.06) | 0.63 | -0.07 (-0.12, -0.02) | 0.01 | 0.14 (0.01, 0.28) | 0.04 | 0.03 (-0.03, 0.1) | 0.33 | 0.01 (-0.1, 0.13) | 0.84 |
| **Household Income** | | | | | | | | | | | | | | | | | | |
| Above $68,900; population median | Reference category | | | | | | | | | | | | | | | | | |
| Below $68,900; population median | 0.01 (0, 0.01) | 0.18 | 0.04 (-0.04, 0.12) | 0.36 | 0.47 (-0.66, 1.6) | 0.42 | 0.20 (-0.6, 1) | 0.63 | -0.02 (-0.09, 0.05) | 0.54 | -0.06 (-0.11, -0.01) | 0.01 | 0.11 (-0.02, 0.23) | 0.09 | -0.01 (-0.06, 0.05) | 0.83 | -0.03 (-0.12, 0.07) | 0.57 |
| **Maternal Education** | | | | | | | | | | | | | | | | | | |
| Postgraduate Degree | Reference category | | | | | | | | | | | | | | | | | |
| Bachelor's Degree | 0.00 (-0.01, 0.02) | 0.57 | 0.04 (-0.1, 0.18) | 0.56 | 1.80 (-0.14, 3.75) | 0.07 | 1.23 (-0.16, 2.62) | 0.08 | 0.03 (-0.09, 0.16) | 0.60 | 0.02 (-0.06, 0.1) | 0.69 | 0.00 (-0.22, 0.23) | 0.98 | 0.05 (-0.04, 0.14) | 0.29 | 0.08 (-0.08, 0.25) | 0.31 |
| Trade/Diploma/Certificate/Other | 0.00 (-0.01, 0.01) | 0.59 | 0.03 (-0.11, 0.16) | 0.69 | 2.64 (0.82, 4.45) | 0.00 | 1.81 (0.5, 3.12) | 0.01 | 0.02 (-0.1, 0.15) | 0.71 | -0.03 (-0.1, 0.05) | 0.52 | 0.10 (-0.11, 0.3) | 0.36 | 0.06 (-0.03, 0.14) | 0.19 | 0.08 (-0.08, 0.24) | 0.31 |
| Less than Year 12 of high school | 0.01 (-0.01, 0.02) | 0.43 | 0.04 (-0.11, 0.18) | 0.60 | 2.83 (0.85, 4.81) | 0.00 | 1.74 (0.3, 3.18) | 0.02 | 0.03 (-0.11, 0.16) | 0.70 | 0.00 (-0.08, 0.09) | 0.93 | 0.10 (-0.13, 0.32) | 0.41 | 0.06 (-0.03, 0.15) | 0.22 | 0.08 (-0.09, 0.25) | 0.35 |

### Table S33. Effect modification (interaction test) by sex, Longitudinal Study of Parents and Children's Child Health CheckPoint

| | Max. cIMT (mm) | | PWV (m/s) | | Systolic Blood Pressure (mmHg) | | Diastolic Blood Pressure (mmHg) | | LDL-C (mmol/L) | | HDL-C (mmol/L) | | Total Triglycerides (mmol/L) | | Remnant Cholesterol (mmol/L) | | Non-HDL-C (mmol/L) | |
|---|---|---|---|---|---|---|---|---|---|---|---|---|---|---|---|---|---|---|
| | Regression Coefficient (95% CI) | P Value | Regression Coefficient (95% CI) | P Value | Regression Coefficient (95% CI) | P Value | Regression Coefficient (95% CI) | P Value | Regression Coefficient (95% CI) | P Value | Regression Coefficient (95% CI) | P Value | Regression Coefficient (95% CI) | P Value | Regression Coefficient (95% CI) | P Value | Regression Coefficient (95% CI) | P Value |
| ***Unadjusted*** | | | | | | | | | | | | | | | | | | |
| **Neighbourhood Disadvantage** | | | | | | | | | | | | | | | | | | |
| High SEIFA | Reference category | | | | | | | | | | | | | | | | | |
| Medium SEIFA | 0.00 (-0.01, 0.01) | 0.82 | 0.02 (-0.11, 0.14) | 0.81 | 0.47 (-1.34, 2.28) | 0.61 | -0.04 (-1.36, 1.28) | 0.95 | -0.02 (-0.13, 0.1) | 0.78 | 0.00 (-0.07, 0.07) | 0.94 | 0.01 (-0.19, 0.21) | 0.90 | 0.03 (-0.06, 0.12) | 0.50 | 0.01 (-0.15, 0.18) | 0.86 |
| Low SEIFA | 0.00 (-0.01, 0.01) | 0.60 | -0.04 (-0.17, 0.1) | 0.59 | 0.09 (-1.75, 1.93) | 0.93 | 0.33 (-1.01, 1.67) | 0.63 | -0.01 (-0.13, 0.1) | 0.81 | -0.03 (-0.1, 0.04) | 0.46 | 0.00 (-0.2, 0.2) | 1.00 | 0.02 (-0.07, 0.11) | 0.64 | 0.01 (-0.16, 0.17) | 0.93 |
| **Household Income** | | | | | | | | | | | | | | | | | | |
| Above $68,900; population median | Reference category | | | | | | | | | | | | | | | | | |
| Below $68,900; population median | 0.00 (-0.01, 0.01) | 0.43 | 0.00 (-0.11, 0.12) | 0.97 | -1.01 (-2.59, 0.57) | 0.21 | -0.50 (-1.63, 0.63) | 0.38 | -0.02 (-0.12, 0.08) | 0.75 | -0.03 (-0.09, 0.04) | 0.42 | 0.03 (-0.14, 0.21) | 0.72 | 0.01 (-0.07, 0.09) | 0.85 | -0.01 (-0.15, 0.13) | 0.90 |
| **Maternal Education** | | | | | | | | | | | | | | | | | | |
| Postgraduate Degree | Reference category | | | | | | | | | | | | | | | | | |
| Bachelor's Degree | 0.00 (-0.02, 0.02) | 0.90 | -0.05 (-0.24, 0.15) | 0.66 | 0.73 (-2.04, 3.5) | 0.60 | -0.25 (-2.25, 1.76) | 0.81 | 0.01 (-0.18, 0.2) | 0.93 | -0.02 (-0.13, 0.09) | 0.72 | -0.01 (-0.34, 0.32) | 0.93 | 0.01 (-0.12, 0.15) | 0.83 | 0.02 (-0.22, 0.27) | 0.85 |
| Trade/Diploma/Certificate/Other | 0.00 (-0.02, 0.01) | 0.76 | -0.07 (-0.26, 0.12) | 0.49 | 0.43 (-2.21, 3.08) | 0.75 | 0.48 (-1.43, 2.39) | 0.62 | 0.00 (-0.18, 0.18) | 0.99 | -0.03 (-0.14, 0.08) | 0.55 | 0.00 (-0.31, 0.3) | 0.98 | 0.02 (-0.12, 0.15) | 0.81 | 0.02 (-0.22, 0.25) | 0.88 |
| Less than Year 12 of high school | 0.00 (-0.02, 0.02) | 0.83 | -0.02 (-0.22, 0.18) | 0.85 | 1.60 (-1.23, 4.43) | 0.27 | 0.96 (-1.1, 3.01) | 0.36 | 0.03 (-0.17, 0.22) | 0.78 | -0.01 (-0.12, 0.11) | 0.91 | -0.02 (-0.37, 0.32) | 0.89 | 0.05 (-0.09, 0.2) | 0.46 | 0.08 (-0.17, 0.34) | 0.53 |
| ***Adjusted*** | | | | | | | | | | | | | | | | | | |
| **Neighbourhood Disadvantage** | | | | | | | | | | | | | | | | | | |
| High SEIFA | Reference category | | | | | | | | | | | | | | | | | |
| Medium SEIFA | 0.00 (-0.01, 0.01) | 0.86 | 0.01 (-0.12, 0.14) | 0.87 | 0.44 (-1.37, 2.25) | 0.63 | -0.01 (-1.33, 1.3) | 0.98 | -0.02 (-0.13, 0.1) | 0.78 | 0.01 (-0.07, 0.08) | 0.88 | 0.01 (-0.19, 0.21) | 0.90 | 0.03 (-0.06, 0.12) | 0.49 | 0.01 (-0.15, 0.18) | 0.86 |
| Low SEIFA | 0.00 (-0.01, 0.01) | 0.58 | -0.04 (-0.17, 0.09) | 0.59 | 0.08 (-1.76, 1.93) | 0.93 | 0.33 (-1.01, 1.66) | 0.63 | -0.02 (-0.13, 0.1) | 0.79 | -0.03 (-0.1, 0.04) | 0.46 | 0.00 (-0.21, 0.2) | 0.99 | 0.02 (-0.07, 0.11) | 0.69 | 0.00 (-0.16, 0.17) | 0.97 |
| **Household Income** | | | | | | | | | | | | | | | | | | |
| Above $68,900; population median | Reference category | | | | | | | | | | | | | | | | | |
| Below $68,900; population median | 0.00 (-0.01, 0.01) | 0.44 | 0.01 (-0.1, 0.12) | 0.82 | -0.96 (-2.54, 0.62) | 0.23 | -0.56 (-1.69, 0.57) | 0.33 | -0.02 (-0.12, 0.08) | 0.70 | -0.03 (-0.09, 0.04) | 0.36 | 0.03 (-0.14, 0.21) | 0.71 | 0.00 (-0.08, 0.08) | 0.94 | -0.02 (-0.16, 0.12) | 0.82 |
| **Maternal Education** | | | | | | | | | | | | | | | | | | |
| Postgraduate Degree | Reference category | | | | | | | | | | | | | | | | | |
| Bachelor's Degree | 0.00 (-0.02, 0.02) | 0.97 | -0.02 (-0.21, 0.18) | 0.88 | 0.87 (-1.9, 3.64) | 0.54 | -0.41 (-2.41, 1.59) | 0.69 | 0.01 (-0.19, 0.2) | 0.96 | -0.03 (-0.14, 0.08) | 0.57 | -0.01 (-0.34, 0.32) | 0.94 | 0.01 (-0.13, 0.15) | 0.91 | 0.01 (-0.23, 0.26) | 0.92 |
| Trade/Diploma/Certificate/Other | 0.00 (-0.02, 0.01) | 0.71 | -0.05 (-0.24, 0.14) | 0.60 | 0.52 (-2.12, 3.16) | 0.70 | 0.39 (-1.51, 2.3) | 0.69 | 0.00 (-0.18, 0.18) | 1.00 | -0.04 (-0.15, 0.07) | 0.47 | 0.00 (-0.31, 0.3) | 0.99 | 0.01 (-0.12, 0.15) | 0.83 | 0.01 (-0.22, 0.25) | 0.90 |
| Less than Year 12 of high school | 0.00 (-0.02, 0.02) | 0.93 | 0.01 (-0.19, 0.21) | 0.90 | 1.77 (-1.06, 4.6) | 0.22 | 0.79 (-1.26, 2.84) | 0.45 | 0.03 (-0.17, 0.23) | 0.78 | -0.02 (-0.14, 0.1) | 0.73 | -0.02 (-0.36, 0.33) | 0.92 | 0.05 (-0.09, 0.19) | 0.49 | 0.08 (-0.18, 0.34) | 0.55 |

#### Table S34. Mediation analysis, vascular measures, Avon Longitudinal Study of Parents and Children

| | Max. cIMT (mm) | | | | PWV (m/s) | | | | SBP (mmHg) | | | | DBP (mmHg) | | | |
|---|---|---|---|---|---|---|---|---|---|---|---|---|---|---|---|---|
| | TCE (95% CI) | P Val | IIE (95% CI) | P Val | TCE (95% CI) | P Val | IIE (95% CI) | P Val | TCE (95% CI) | P Val | IIE (95% CI) | P Val | TCE (95% CI) | P Val | IIE (95% CI) | P Val |
| **Neighbourhood Disadvantage** | | | | | | | | | | | | | | | | |
| Low Deprivation | Reference group | | | | | | | | | | | | | | | |
| Mediation Deprivation | -0.001 (-0.005, 0.003) | 0.60 | -0.0004 (-0.0009, 0.0001) | 0.13 | 0.02 (-0.04, 0.07) | 0.52 | 0.002 (-0.004, 0.009) | 0.45 | 0.8 (-0.02, 1.5) | 0.06 | 0.5 (0.2, 0.8) | 0.00 | 0.4 (-0.1, 0.9) | 0.13 | 0.3 (0.1, 0.5) | 0.00 |
| High Deprivation | -0.004 (-0.009, 0.00004) | 0.05 | -0.0003 (-0.001, 0005) | 0.46 | 0.05 (-0.01, 0.1) | 0.12 | 0.007 (-0.005, 0.02) | 0.27 | 0.9 (0.09, 1.8) | 0.03 | 1.0 (0.6, 1.3) | 0.00 | 0.8 (0.3, 1.4) | 0.005 | 0.6 (0.4, 0.8) | 0.00 |
| **Financial Difficulties** | | | | | | | | | | | | | | | | |
| No Difficulties | Reference group | | | | | | | | | | | | | | | |
| Difficulties | -0.003 (-0.006, 0.0007) | 0.13 | -0.0002 (-0.0006, 0.0003) | 0.47 | 0.03 (-0.01, 0.08) | 0.17 | 0.003 (-0.004, 0.009) | 0.40 | 0.7 (0.1, 1.3) | 0.02 | 0.4 (0.2, 0.7) | 0.00 | 0.2 (-0.2, 0.6) | 0.32 | 0.3 (0.1, 0.4) | 0.00 |
| **Maternal Education** | | | | | | | | | | | | | | | | |
| University Degree | Reference group | | | | | | | | | | | | | | | |
| A Levels | -0.005 (-0.01, -0.0002) | 0.04 | -0.000006 (-0.0005, 0.0004) | 0.79 | 0.002 (-0.06, 0.07) | 0.96 | 0.003 (-0.004, 0.01) | 0.45 | 0.8 (-0.09, 1.6) | 0.08 | 0.5 (0.2, 0.8) | 0.002 | 0.5 (-0.09, 1.1) | 0.10 | 0.3 (0.1, 0.5) | 0.002 |
| O Levels | -0.004 (-0.01, 0.0007) | 0.10 | -0.0006 (-0.001, 0.00007) | 0.08 | 0.06 (-0.003, 0.1) | 0.06 | 0.005 (-0.005, 0.01) | 0.32 | 0.8 (0.3, 2.0) | 0.006 | 0.8 (0.5, 1.1) | 0.00 | 1.0 (0.4, 1.6) | 0.001 | 0.5 (0.3, 0.7) | 0.00 |
| Vocational/CSE | -0.007 (-0.01, -0.002) | 0.005 | -0.0001 (-0.001, 0.001) | 0.81 | 0.08 (0.009, 0.2) | 0.03 | 0.003 (-0.02, 0.02) | 0.78 | 2.3 (1.4, 3.3) | 0.00 | 1.1 (0.8, 1.5) | 0.00 | 1.7 (1.1, 2.4) | 0.00 | 0.7 (0.5, 1.0) | 0.00 |

#### Table S35. Mediation analysis, metabolic measures, Avon Longitudinal Study of Parents and Children

| | LDL-C (mmol/L) | | | | HDL-C (mmol/L) | | | | Total Triglycerides (mmol/L) | | | | Remnant-C (mmol/L) | | | | Non-HDL-C (mmol/L) | | | |
|---|---|---|---|---|---|---|---|---|---|---|---|---|---|---|---|---|---|---|---|---|
| | **TCE (95% CI)** | **P Val** | **IIE (95% CI)** | **P Val** | **TCE (95% CI)** | **P Val** | **IIE (95% CI)** | **P Val** | **TCE (95% CI)** | **P Val** | **IIE (95% CI)** | **P Val** | **TCE (95% CI)** | **P Val** | **IIE (95% CI)** | **P Val** | **TCE (95% CI)** | **P Val** | **IIE (95% CI)** | **P Val** |
| **Neighbourhood Disadvantage** | | | | | | | | | | | | | | | | | | | | |
| Low Deprivation | Reference group | | | | | | | | | | | | | | | | | | | |
| Mediation Deprivation | -0.006 (-0.05, 0.04) | 0.77 | 0.0003 (-0.002, 0.008) | 0.28 | -0.01 (-0.04, 0.01) | 0.36 | -0.004 (-0.008, 0.0004) | 0.08 | 0.02 (-0.03, 0.07) | 0.40 | 0.009 (0.002, 0.02) | 0.02 | -0.0007 (-0.02, 0.02) | 0.95 | 0.005 (0.001, 0.009) | 0.01 | -0.003 (-0.05, 0.04) | 0.89 | 0.008 (0.0005, 0.01) | 0.04 |
| High Deprivation | -0.02 (-0.06, 0.03) | 0.48 | 0.003 (-0.006, 0.01) | 0.53 | -0.02 (-0.05, 0.006) | 0.11 | -0.007 (-0.01, -0.0003) | 0.36 | 0.02 (-0.03, 0.08) | 0.36 | 0.02 (0.005, 0.03) | 0.004 | 0.003 (-0.02, 0.03) | 0.78 | 0.008 (0.003, 0.01) | 0.002 | -0.01 (-0.07, 0.04) | 0.67 | 0.01 (0.002, 0.02) | 0.02 |
| **Financial Difficulties** | | | | | | | | | | | | | | | | | | | | |
| No Difficulties | Reference group | | | | | | | | | | | | | | | | | | | |
| Difficulties | -0.01 (-0.05, 0.02) | 0.46 | 0.002 (-0.003, 0.007) | 0.49 | -0.02 (0.04, 0.006) | 0.15 | -0.003 (-0.006, 0.001) | 0.19 | -0.0003 (-0.04, 0.04) | 0.99 | 0.007 (0.0002, 0.01) | 0.05 | -0.005 (-0.02, 0.01) | 0.59 | 0.004 (0.0001, 0.007) | 0.04 | -0.02 (-0.06, 0.02) | 0.37 | 0.005 (-0.001, 0.01) | 0.11 |
| **Maternal Education** | | | | | | | | | | | | | | | | | | | | |
| University Degree | Reference group | | | | | | | | | | | | | | | | | | | |
| A Levels | -0.008 (-00.06, 0.04) | 0.75 | 0.003 (-0.004, 0.007) | 0.52 | -0.006 (-0.04, 0.03) | 0.73 | -0.004 (-0.008, 0.0007) | 0.10 | 0.03 (-0.03, 0.08) | 0.34 | 0.008 (0.0003, 0.02) | 0.04 | -0.004 (-0.03, 0.02) | 0.78 | 0.004 (0.000006, 0.008 | 0.05 | -0.01 (-0.07, 0.04) | 0.68 | 0.006 (-0.001, 0.01) | 0.12 |
| O Levels | -0.02 (-0.06, 0.03) | 0.51 | 0.004 (-0.003, 0.01) | 0.25 | -0.02 (-0.05, 0.03) | 0.36 | -0.006 (-0.01, -0.0003) | 0.04 | 0.05 (-0.01, 0.1) | 0.13 | 0.01 (0.004, 0.02) | 0.004 | -0.007 (-0.03, 0.02) | 0.60 | 0.008 (0.003, 0.01) | 0.001 | -0.02 (-0.07, 0.03) | 0.46 | 0.01 (0.003, 0.02) | 0.01 |
| Vocational/CSE | -0.003 (-0.06, 0.05) | 0.91 | 0.005 (-0.01, 0.02) | 0.52 | -0.03 (-0.07, 0.007) | 0.11 | -0.008 (-0.02, 0.002) | 0.12 | 0.05 (-0.01, 0.1) | 0.11 | 0.02 (0.003, 0.04) | 0.02 | 0.01 (-0.02, 0.04) | 0.49 | 0.01 (0.003, 0.02) | 0.009 | 0.009 (-0.05, 0.07) | 0.78 | 0.02 (-0.0008, 0.03) | 0.06 |
